# Impact of the COVID-19 pandemic and vaccination on global, regional, and national burden of Guillain-Barré syndrome: observational study

**DOI:** 10.1101/2024.07.09.24309613

**Authors:** Xīn Gào, Chen Zhao, Junting Yang, Ziming Yang, Jingnan Feng, Siyan Zhan, Dongsheng Fan, Zhike Liu

## Abstract

**Objectives:** To estimate the global, regional, and national burden and trends of Guillain-Barré Syndrome (GBS), and to explore the impact of the COVID-19 pandemic and related public health measures on the GBS burden.

**Design:** Observational study.

**Setting:** 204 countries and territories.

**Data sources:** Global Burden of Diseases, Injuries, and Risk Factors Study (GBD) 2021 and Our World in Data.

**Main outcome measures:** All-cause and COVID-19-specific age-standardized years lived with disability (YLD) rates and estimated annual percentage change (EAPC).

**Results:** Globally, the burden of GBS significantly increased during the COVID-19 pandemic, age-standardized YLD rates were 0.86 (95% uncertainty interval (UI): 0.56 to 1.24) per 100,000 people in 2020, rising to 1.75 (95% UI: 1.12 to 2.54) per 100,000 people in 2021, reflecting an EAPC of 70.35% (95% UI: 38.73 to 109.06) from 2019 to 2021. Regionally, the highest age-standardized YLD rates in 2020 and 2021 were observed in the high-income Asia Pacific and central Latin America, respectively, while the lowest rates were found in East Asia for both years. Age-standardized YLD rates rose 3.7 times faster in countries with a low socio-demographic index (SDI) versus countries with a high SDI (103.21% per year (95% UI: 71.10 to 141.35%) vs 28.02% per year (11.46 to 47.03)). Males generally had higher YLD rates across all age groups, with the highest disparity in those aged 80 and above. During the first two years of the pandemic, the increase in YLD rates was more pronounced in females and younger populations aged 15-29 years. A statistically negative correlation was observed between COVID-19 vaccination coverage and the GBS burden, with increases in full vaccination coverage per 100 people leading to decreases in all-cause YLD rates by 0.0070 and COVID-19-specific YLD rates by 0.0128 per 100,000 people.

**Conclusion:** Predominantly driven by the surge of COVID-19 cases, the burden of GBS significantly increased during the first two years of the pandemic, especially in those countries with lower SDI levels. Greater increases in the GBS burden were observed in females and younger populations (15-29 years). Importantly, the benefits of vaccination outweighed the risks in controlling the post-COVID-19 GBS burden, although significant disparities in vaccination coverage existed between countries. These findings offer robust evidence facilitating public education and transparent communication on COVID-19 vaccination and highlight the urgency for targeted control measures to manage the burden of post-infection complications, such as GBS, during future COVID-19 waves or other potential pandemics.

**WHAT IS ALREADY KNOWN ON THIS TOPIC:** Guillain-Barré syndrome (GBS) is an autoimmune disease, and respiratory and gastrointestinal infections are the major triggering factors.

From 1990 to 2019, the burden of GBS was relatively low, with negligible changes observed in most countries and territories.

The impact of the COVID-19 pandemic and vaccination on the burden of GBS remains not fully understood.

**WHAT THIS STUDY ADDS:** During the COVID-19 pandemic, the global age-standardized years lived with disability (YLD) related to GBS increased by 70.30% annually, and low socio-demographic index (SDI) regions experienced the most rapid GBS burden increment (103.21%).

The severe acute respiratory syndrome coronavirus 2 (SARS-CoV-2) infection was the primary factor contributing to the increased burden of GBS during the COVID-19 pandemic.

Improving vaccination coverage was the most effective measure to reduce the post-COVID-19 burden of GBS, and each additional fully vaccinated person (per hundred people) decreased the COVID-19-specific age-standardized YLD of GBS by 0.0128 per 100,000 population.

## Introduction

Guillain-Barré syndrome (GBS) is an acute-onset autoimmune polyradiculoneuropathy, which is the most common cause of acute flaccid paralysis worldwide.^1^ This condition can lead to life-threatening progressive motor weakness and up to 30% of patients with GBS require admission to the intensive care unit and mechanical ventilation due to respiratory failure.^2^ ^3^ Approximately two-thirds of GBS cases present antecedent infections, mostly respiratory and gastrointestinal infections.^4^ The prevalence of GBS can show a transient rise in outbreaks of certain infectious diseases. An example is the surge of GBS following the 2015–2016 Zika virus outbreak in Latin America and the Caribbean, with a 2.6-fold increase in incidence above the background rate.^5^ More recently, the severe acute respiratory syndrome coronavirus 2 (SARS-CoV-2) caused a global pandemic resulting in 776 million infected cases and 7 million deaths to date.^6^ Cumulating evidence suggests a potential link between SARS-CoV-2 infection and GBS.^7–10^ However, comprehensive estimates of the global GBS burden during the pandemic remain lacking. Existing studies are largely limited to smaller populations, leaving substantial gaps in data on GBS related to COVID-19 that need to be addressed urgently.

As one of the most cost-effective and efficient measures for COVID-19 prevention and control, vaccination has raised public concerns about the safety, particularly regarding the risk of GBS.^11^(ref) Two nationwide database studies have consistently shown a higher incidence of GBS following adenovirus-based vaccines, while mRNA-based vaccines have not demonstrated this association.^12^ ^13^ On the other hand, vaccination has the potential to reduce the burden of GBS by preventing SARS-CoV-2 transmission and infection. Therefore, it is urged to investigate the benefits and risks at the population level to provide evidence-based support for immunization policy decisions. To the best of our knowledge, the overall impact of global vaccination efforts on the GBS burden has not been investigated.

Although the Global Burden of Disease, Injuries, and Risk Factors Study (GBD) 2021 has recently updated the prevalence and years lived with disability (YLDs) of GBS, the results did not account for GBS cases attributable to COVID-19. Instead, the study grouped GBS and persistent cognitive symptoms following COVID-19 under the category of neurological complications related to COVID-19 for pooled analysis.^14^ In our study, we aimed to conduct a comprehensive analysis of the global, regional, and national prevalence and YLDs of GBS across all age groups, sexes, and causes, between 1990 and 2021 with a particular focus on the first two years of the COVID-19 pandemic. Furthermore, we explored public health factors, including vaccination and non-pharmacological interventions, that might influence the GBS burden and tested the hypothesis that widespread COVID-19 vaccination does not increase the GBS burden during the pandemic.

## Methods

### Study design and data sources

The GBD is a comprehensive epidemiological database coordinated by the Institute for Health Metrics and Evaluation (IHME).^15^ In GBD 2021, the burden of 371 diseases and injuries was estimated by calculating the incidence, point prevalence (hereafter referred to as prevalence), YLDs, years of life lost, and disability-adjusted life years for 204 countries and territories using data from 100,983 sources.^16^ GBD 2021 classified GBS as an impairment that is considered a consequence of a range of causes in the disease and injury hierarchy, and for which there tends to be a higher total occurrence of the impairment rather than the cause-specific estimates. GBS was identified using the International Classification of Diseases (ICD-10) code G61.0 and ICD-9 code 357.0 (acute infective polyneuritis). We obtained data on the global, regional, and national prevalence and YLDs of GBS between 1990 and 2021 (https://vizhub.healthdata.org/gbd-results/).^17^ The trends in the burden of GBS over the past three decades, as well as during the first two years of the COVID-19 pandemic, were analyzed in this investigation. The socio-demographic index (SDI) was employed for the correlation analysis between the social and economic development of each region or country and the GBS burden.^18^

Our World in Data (OWID) is an online scientific publication covering topics on poverty, public health, education, climate change, and other critical issues.^19^ This study extracted data on COVID-19 vaccination coverage in 2021^20^ and the government stringency index (GSI) in both 2020 and 2021^21^ from OWID to investigate the impact of public health measures on the burden of GBS during the first two years of the pandemic (https://ourworldindata.org/). The GSI is a composite score of nine metrics, including school closures, workplace closures, cancellation of public events, restrictions on public gatherings, closures of public transport, stay-at-home requirements, public information campaigns, restrictions on internal movements, and international travel controls, which tracks the strictness of government responses to the COVID-19 pandemic.^21^

The GBD study adhered to the Guidelines for Accurate and Transparent Health Estimates Reporting (GATHER) statement ^22^ and obtained ethical clearance from the Institutional Review Board of the University of Washington (study 9060). OWID used publicly available data from reliable sources, including the World Health Organization (WHO), the National Ministry of Health, and the Centers for Disease Control and Prevention (CDC). Secondary analysis of the de-identified data from GBD and OWID did not require additional ethical approval.

### YLDs

This study considered YLDs as the primary measure of disease burden, which represents health loss due to nonfatal diseases. YLDs were calculated by multiplying the prevalence of GBS by its corresponding disability weight, and it reflected the severity of the disability on a scale from 0 (perfect health) to 1 (equivalent to death).^23^ In GBD 2021, the prevalence of GBS was estimated using DisMod-MR 2.1 (disease model–Bayesian meta-regression), which synthesized data from multiple sources, accounting for variations in study design, case definition, and diagnostic criteria.^16^ Moreover, we extracted YLDs of GBS attributed to any reported etiological cause, including lower respiratory infections, upper respiratory infections, COVID-19, diarrheal diseases, Zika virus, other unspecified infectious diseases, and other neurological disorders, i.e., cases with unknown specific causes.

### Statistical analysis

Prevalence and YLDs of GBS and their corresponding 95% uncertainty intervals (UIs) were reported to summarize the GBS burden for the globe, 21 GBD regions, and 204 countries and territories. Male-to-female YLD ratios were calculated across calendar years, all age groups, and all locations.

To measure the age-standardized YLD rate temporal trends over 1990-2019 and 2019-2021, estimated annual percentage changes (EAPCs) were computed by fitting a linear regression model to the log-transformed age-standardized YLD rate, as described by Hankey and colleagues.^24^ We additionally performed joinpoint regression models to identify "points" where a significant change in the trend of a data series occurred.^25^ These points represented shifts in the underlying trend, which could be due to various factors or interventions. Each segment between two adjacent points represented a different linear trend. Weighted Bayesian Information Criterion (WBIC) method was used to select the model with the lowest WBIC, which balanced goodness-of-fit with model complexity.^26^

A three-step approach was applied to investigate the link between the GBS burden and potential influencing factors during the COVID-19 pandemic. Firstly, we explored the strength and direction of the relationships between age-standardized YLD rates of GBS, SDI, COVID-19 incidence, COVID-19 vaccination coverage rates, and GSI using scatter plots and Pearson’s correlation coefficients. Secondly, we used generalized linear models (GLIMs) with a Gaussian distribution and log-link function to assess the association between vaccination coverage rates (as of December 31, 2021) and age-standardized YLD rates in 2021. We developed two models: a comprehensive model adjusted for SDI, median GSI in 2021, and YLD rates in 2020, and a minimal sufficient model adjusted only for SDI. The minimal sufficient model was selected using a directed acyclic graph (**supplementary figure 1**). Furthermore, the direct effect of vaccination on the GBS burden was evaluated with a model adjusted for SDI and age-standardized incidence rate of COVID-19. Thirdly, we performed a generalized additive model (GAM) to investigate the potential non-linear shape of the association between age-standardized YLD rates of GBS and vaccination coverage rates.

R (version 4.3.3 and 4.4.0) and Joinpoint Software (version 5.2.0) were used for statistical analysis and data visualization. Two-sided hypothesis tests were conducted with a statistical significance level of 0.05. The Methods section in the supplementary file provides more details on the statistical analyses.

### Patient and public involvement

The GBD is a global observational epidemiological study designed to report mortality and disability for major diseases. OWID is an open-access online database that provides comprehensive information on a wide range of global issues. Our study used secondary data from these databases without direct access to the participants. No patients were involved in setting the research question, collecting and analyzing the data, interpreting the results, or writing the manuscript.

## Results

### Spatial distribution and temporal trends

Globally, 230.1 thousand (95% UI: 194.3 to 269.7) and 471.9 thousand (95% UI: 389.2 to 554.1) individuals were affected by GBS in 2020 and 2021, respectively (**supplementary table S1**). GBS accounted for 68.1 thousand (95% UI: 44.8 to 98.6) YLDs in 2020, with an age-standardized YLD rates of 0.86 (95% UI: 0.56 to 1.24) per 100,000 people, and 139.6 thousand (95% UI: 90.4 to 202.4) YLDs in 2021, with an age-standardized YLD rate of 1.75 (95% UI: 1.12 to 2.54) per 100,000 people (**table 1**). The EAPCs in age-standardized YLD rate were 70.35% (95% UI: 38.73‒109.06) for the period from 2019 to 2021, and were 0.16% (95% UI: 0.14 to 0.18) over the longer period from 1990 to 2019 (**table 1**).

**Table 1.**
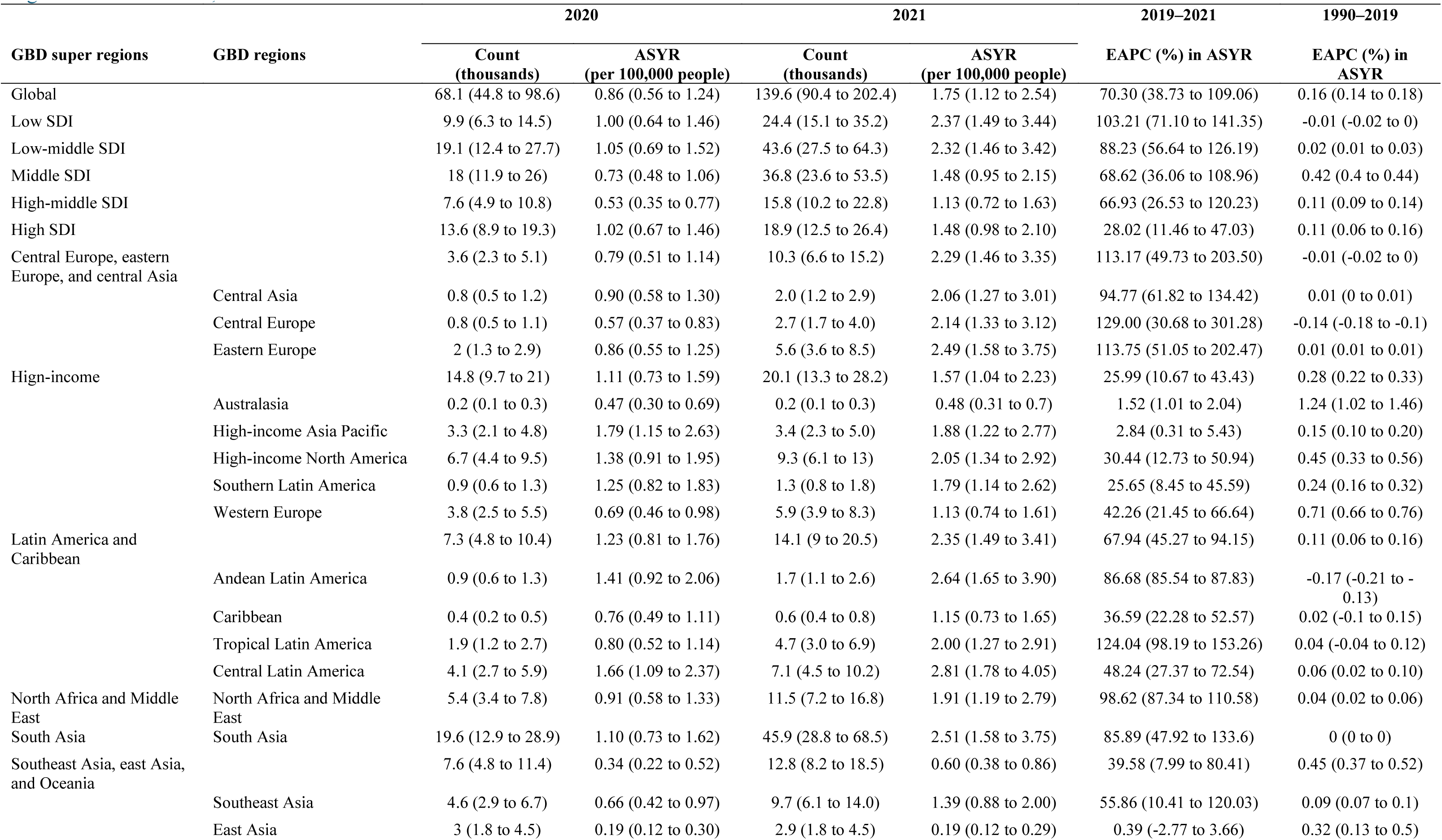
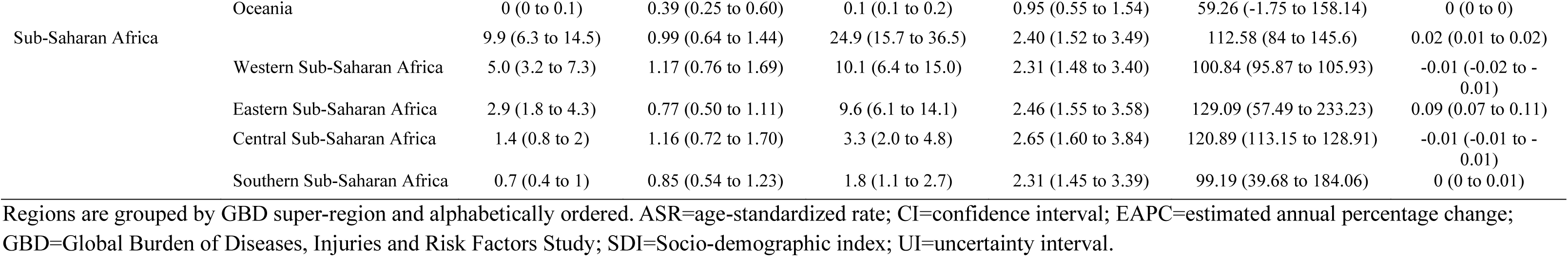
Years lived with disability of Guillain-Barrésyndrome in 2020 and 2021 and the estimated annual percentage change in the age-standardised rates, at global, regional and local levels, from 1990 to 2021.

At the regional level, age-standardized YLD rates were highest in the high-income Asia Pacific region (1.79 [95% UI: 1.15 to 2.63] per 100,000 people) and central Latin America (2.81 [1.78 to 4.05] per 100,000 people) in 2020 and 2021, respectively (**table 1**). The lowest age-standardized YLD rates were observed in East Asia for both 2020 (0.19 [95% UI: 0.12 to 0.30] per 100,000 people) and 2021 (0.19 [95% UI: 0.12 to 0.29] per 100,000 people). During the first two years of the COVID-19 pandemic, Central Europe and Eastern Sub-Saharan Africa experienced the most rapid increase in the GBS burden (**supplementary figure S2**). EAPCs in age-standardized YLD rates were 129.00% (30.68 to 301.28) and 129.09% 57.49 to 233.23), respectively (**table 1**). East Asia presented the lowest EAPC in age-standardized YLD rates, which was 0.39% (-2.77 to 3.66). Between 1990 and 2019, a relatively faster increase in age-standardized YLD rates was observed in high-income regions with EAPCs ranging from 0.15% to 1.24%. Age-standardized YLD rates slightly but statistically significantly decreased in Central Europe, Andean Latin America, Western Sub-Saharan Africa, and Central Sub-Saharan Africa with EAPCs ranging from -0.17% to -0.01%. Notably, Joinpoint regression analysis showed that the age-standardized YLD rates of GBS significantly elevated in the Caribbean in 2016 (**supplementary figure S3**).

Among 204 countries and territories, Singapore, Japan, and Mexico presented the most severe burden of GBS in 2020, while Iraq, the Plurinational State of Bolivia, and North Macedonia had the most severe burden of GBS in 2021 (**figure 1**, **figure 2** and **supplementary table S2**). The lowest burden of GBS was observed in East Asian countries and territories, including China, Taiwan (Province of China), and the Democratic People’s Republic of Korea, during the first two years of the COVID-19 pandemic. Fifty-nine out of 204 countries and territories experienced a rapid increase in GBS burden between 2019 and 2021, with EAPCs in age-standardized YLD rates greater than 100% (**figure 3** and **supplementary table S2**). The long-term EAPC remained relatively stable or showed minimal changes for most countries during 1990-2019.

**Figure 1.**
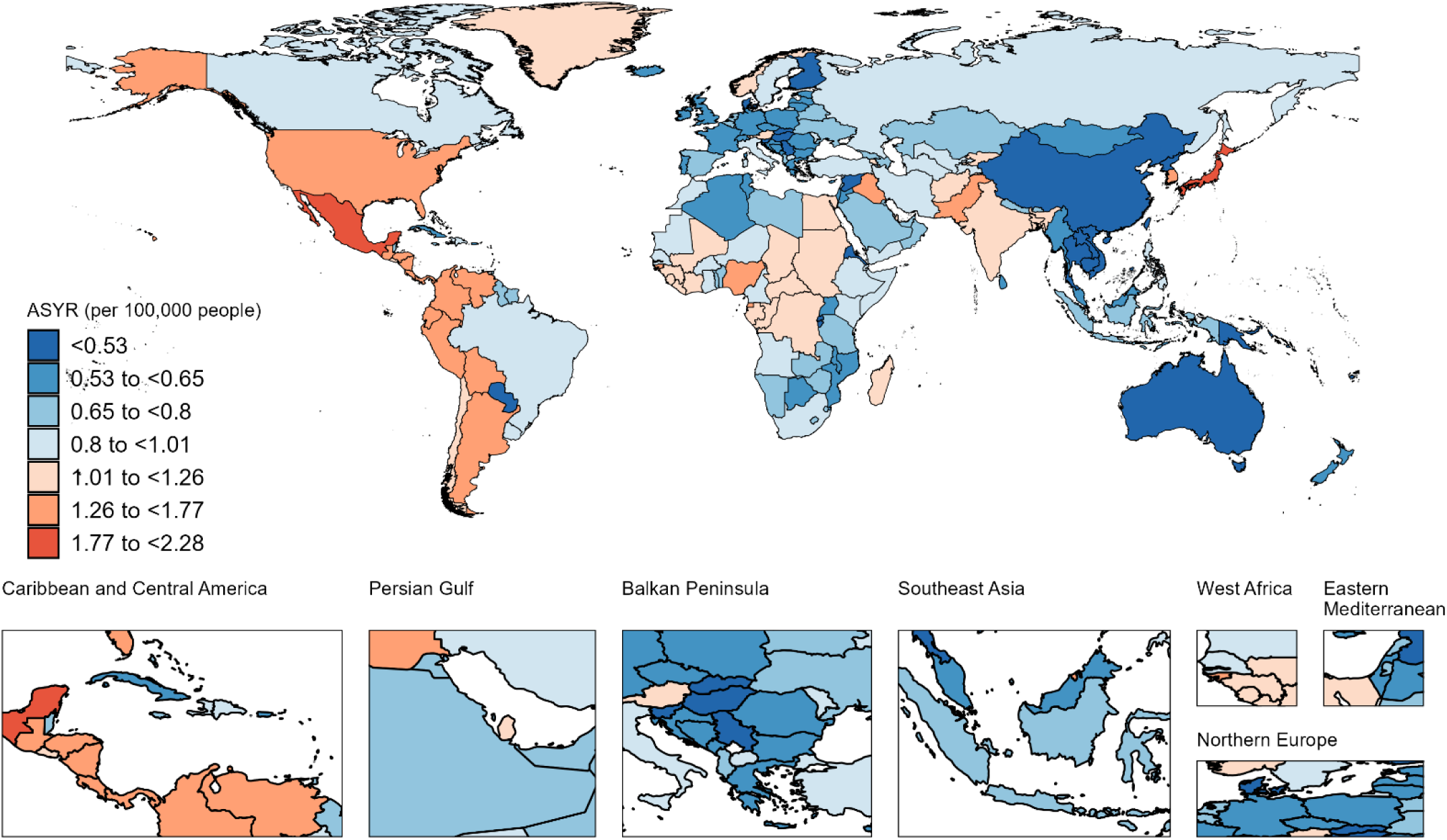
Age-standardised years lived with disability rate (ASYR) of Guillain-Barré syndrome per 100,000 people in 2020.

**Figure 2.**
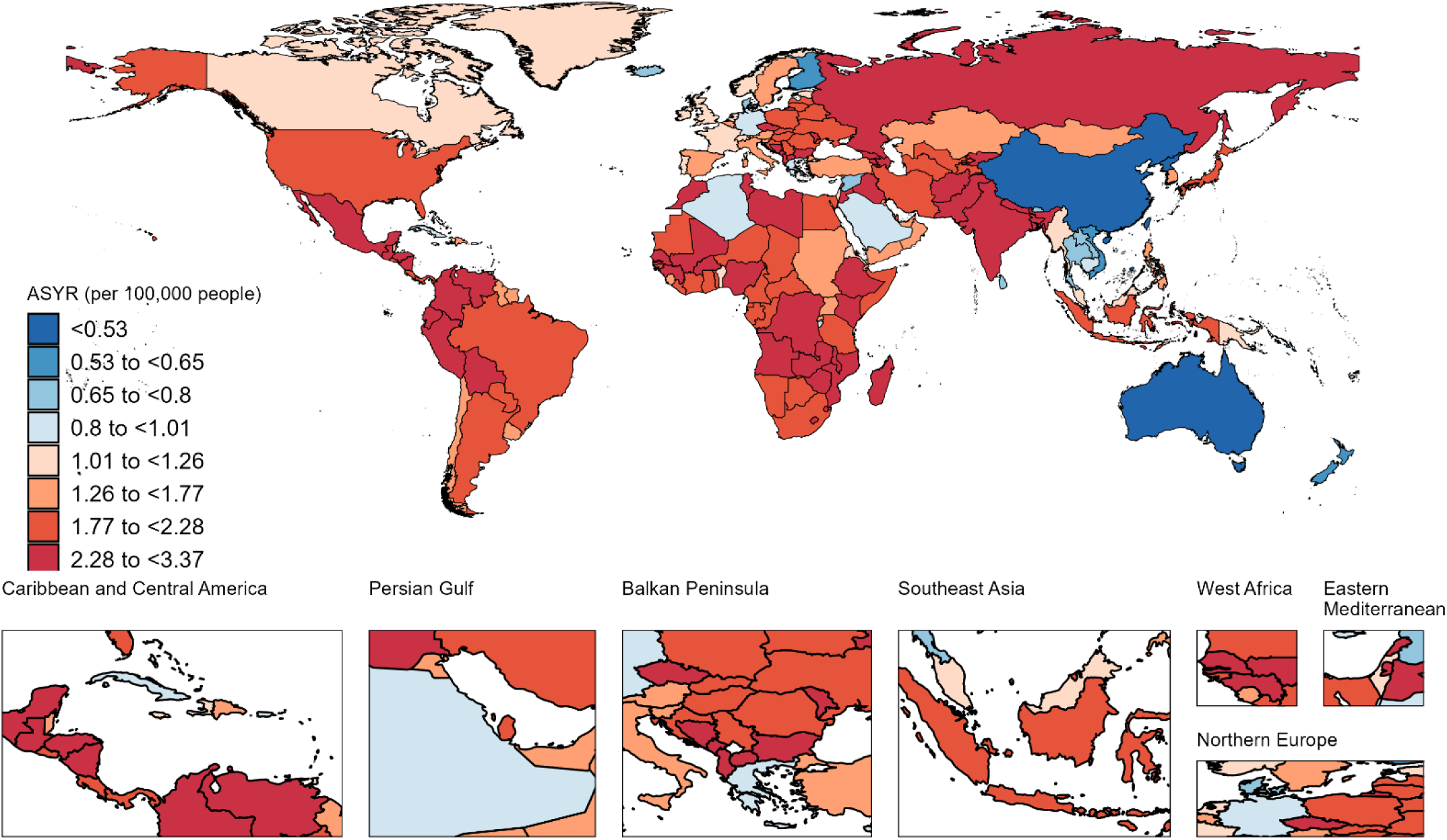
Age-standardised years lived with disability rate (ASYR) of Guillain-Barré syndrome per 100,000 people in 2021.

**Figure 3.**
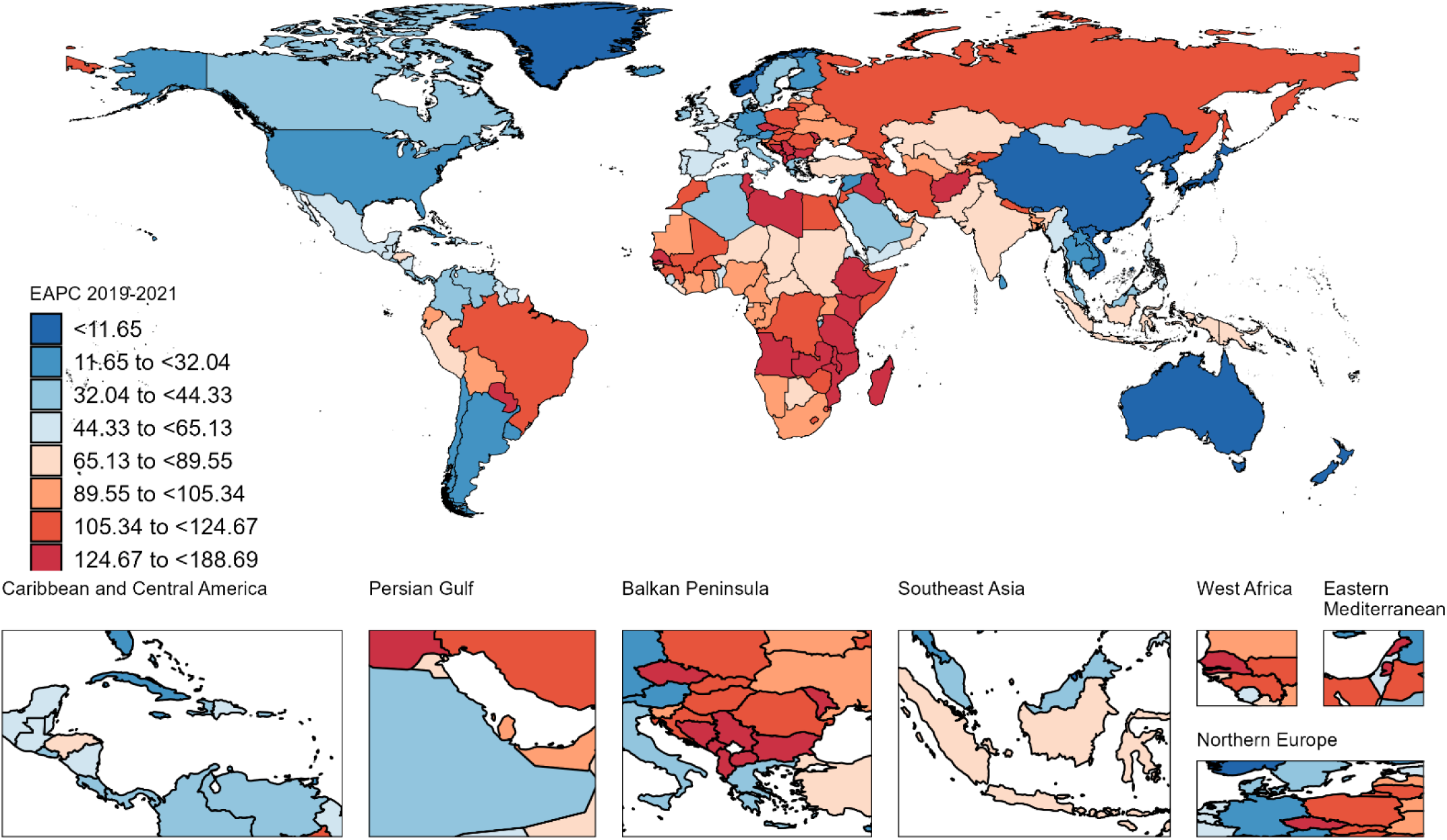
Estimated annual percentage changes (EAPC) in age-standardised years lived with disability rate of Guillain-Barré syndrome per 100,000 people between 2019 and 2021.

### Patterns by sex and age

All-cause YLD rates of GBS differed between sexes, and the extent of this difference varied across age groups and by geographic locations. In general, male YLD rates were higher than female YLD rates in all age groups, with considerable heterogeneity across countries and territories (**figure 4A**). The greatest variability in the YLD rate male-to-female ratio was observed in the age group of 80 years and older. In 2021, the YLD rate male-to-female ratios were broadly lower than those in 1990, 2010, and 2019, indicating a decreased disparity in the GBS burden between males and females. **Figure 4B** shows that all-cause YLD rates of GBS increased more rapidly in females than in males across all age groups during the COVID-19 pandemic. Moreover, we observed that COVID-19-specific age-standardized YLD rates were similar among males and females, and the age-standardized YLD rates male-to-female ratios were close to 1, worldwide and across all regions (**supplementary table S3**).

**Figure 4.**
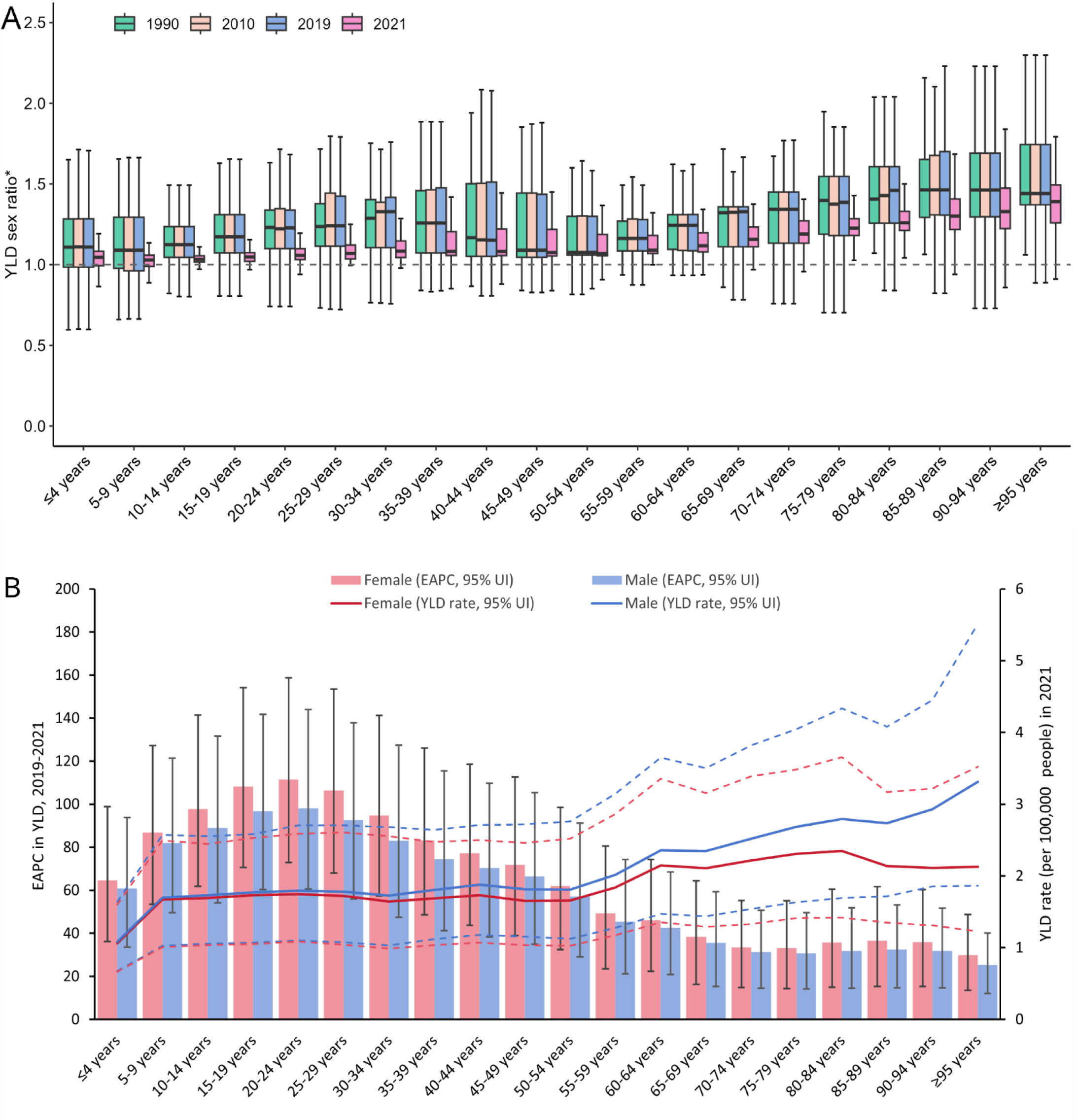
Age-sex distribution of Guillain-Barré syndrome burden: (A) all-cause years lived with disability (YLDs) rate male-to-female ratio by age in 1990, 2010, 2019, and 2021; (B) YLDs rate in 2021 and estimated annual percentage change (EAPC) in YLDs between 2019 and 2021, by age and sex.

In 2021, the global burden of GBS was the lowest in children under the age of five and increased with advancing age in both sexes (**figure 4B** and **supplementary figure S5**). The highest YLD rates were observed in individuals over the age of 94 (2.45 [95% UI: 1.39 to 4.07] per 100,000 people). However, during 2019-2021, YLD rates increased more rapidly in younger age groups, particularly among those aged 15–29. The EAPC ranged from 106.31% to 111.45% among females and from 92.53% to 98.03% among males (**figure 4B**).

### Variation by SDI

Before the COVID-19 pandemic, age-standardized YLD rates were positively associated with SDI levels in high-income regions (**supplementary figure S6**). Over the period from 2019 to 2021, the EAPC in age-standardized YLD rates showed significant increases across regions, reflecting the global rise in GBS burden (**table 1**). However, this increase was more pronounced in regions with lower SDI levels (**table 1**). **Figure 5** shows a statistically significant association between EAPC in age-standardized YLD rates and SDI levels across 204 countries and territories (Pearson’s R = -0.26, *p* < 0.001). In 2020, age-standardized YLD rates of regions with low SDI (1.00 [0.64 to 1.46] per 100,000 people), low-middle SDI (1.05 [0.69 to 1.52] per 100,000 people), and high SDI (1.02 [0.67 to 1.46] per 100,000 people) were above the global level (**table 1**). In 2021, regions with low SDI and low-middle SDI experienced the highest age-standardized YLD rates, which were 2.37 (1.49 to 3.44) per 100,000 people and 2.32 (1.46 to 3.42) per 100,000 people, respectively.

**Figure 5.**
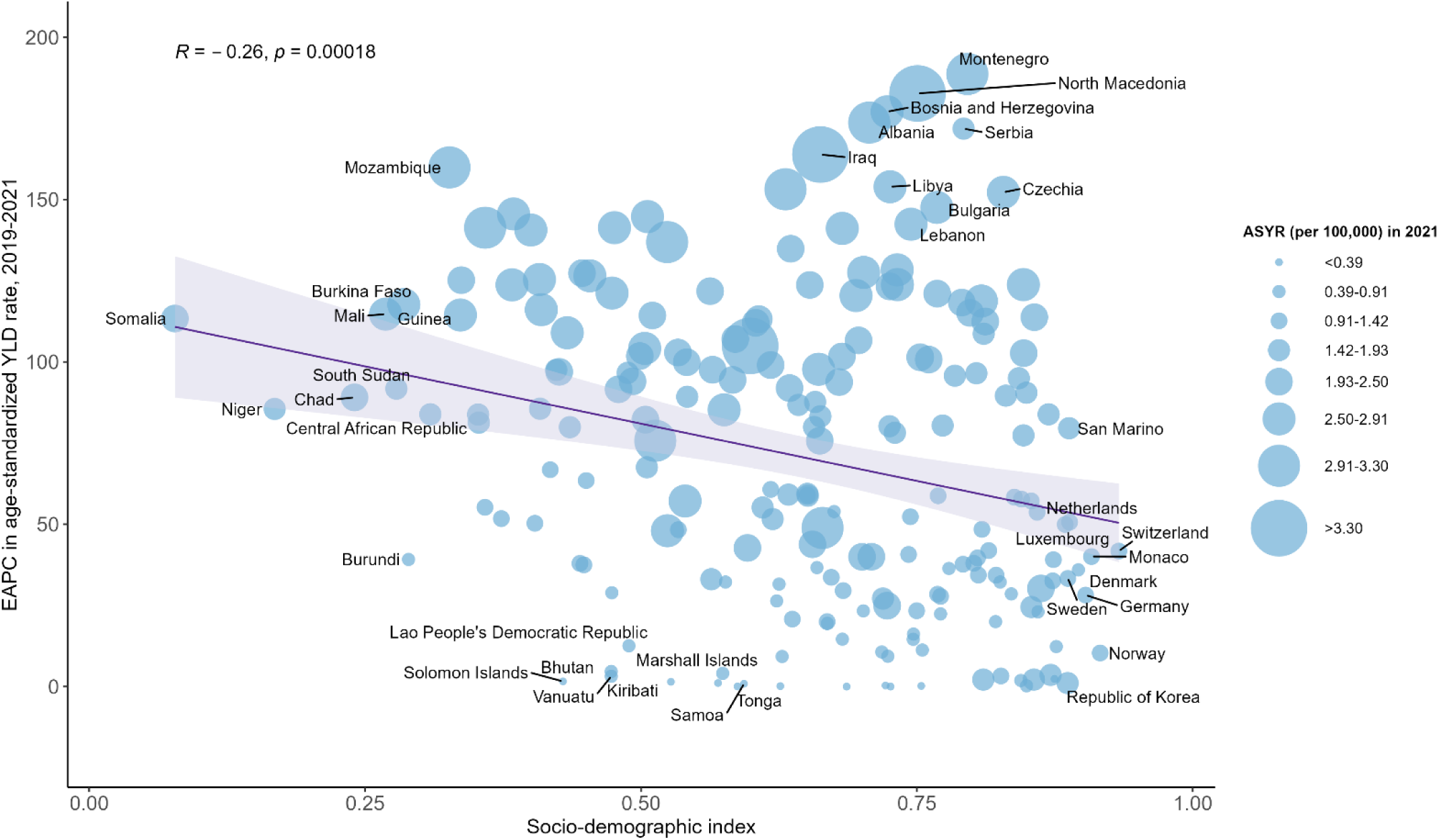
Association between esimated annual percentage change in age-standardised years lived with disability rate during the COVID-19 pandemic, across 204 countries and territories. ASYR=age-standardized yeas lived with disability rate; EAPC=estimated annual percentage change; YLD=years lived with disability. Notes: Pearson correlation coefficient (R) and *p* value were presented to show the strength and direction of the association. Akaike Information Criterion (AIC) was used to compare the linear model and the smooth models, and the AIC score indicated both models had similar performance.

### Increased GBS burden attributed to COVID-19

At the global level, the YLDs attributable to specific causes of GBS remained stable between 1990 and 2019, with respiratory infections and other neurological disorders accounting for over half of the GBS burden **(supplementary figure S8** and **supplementary table S4**). However, during the COVID-19 pandemic, the coronavirus became the primary driver of the increase in GBS YLDs across all ages, sexes, and GBD regions (**supplementary figure S9-S12**). In 2020, the global leading causes of GBS YLDs were COVID-19 (cause-specific YLD rate: 0.25 [95% UI 0.16 to 0.38] per 100,000 people), upper respiratory infections (0.19 [0.12 to 0.3] per 100,000 people), and other neurological disorders (0.23 [0.13 to 0.36] per 100,000 people) (**figure 6** and **supplementary table S4**). In 2021, the global cause-specific YLD rates were 1.14 (0.71 to 1.72) per 100,000 people, 0.23 (0.13 to 0.36) per 100,000 people, and 0.19 (0.12 to 0.30) per 100,000 people for COVID-19, upper respiratory infections, and other neurological disorders. Most regions experienced a higher burden of GBS due to COVID-19 compared to the global average, except for high-income regions, Southeast Asia, East Asia, and Oceania, which had lower burdens in both 2020 and 2021 (**figure 6 and supplementary table S4**).

**Figure 6.**
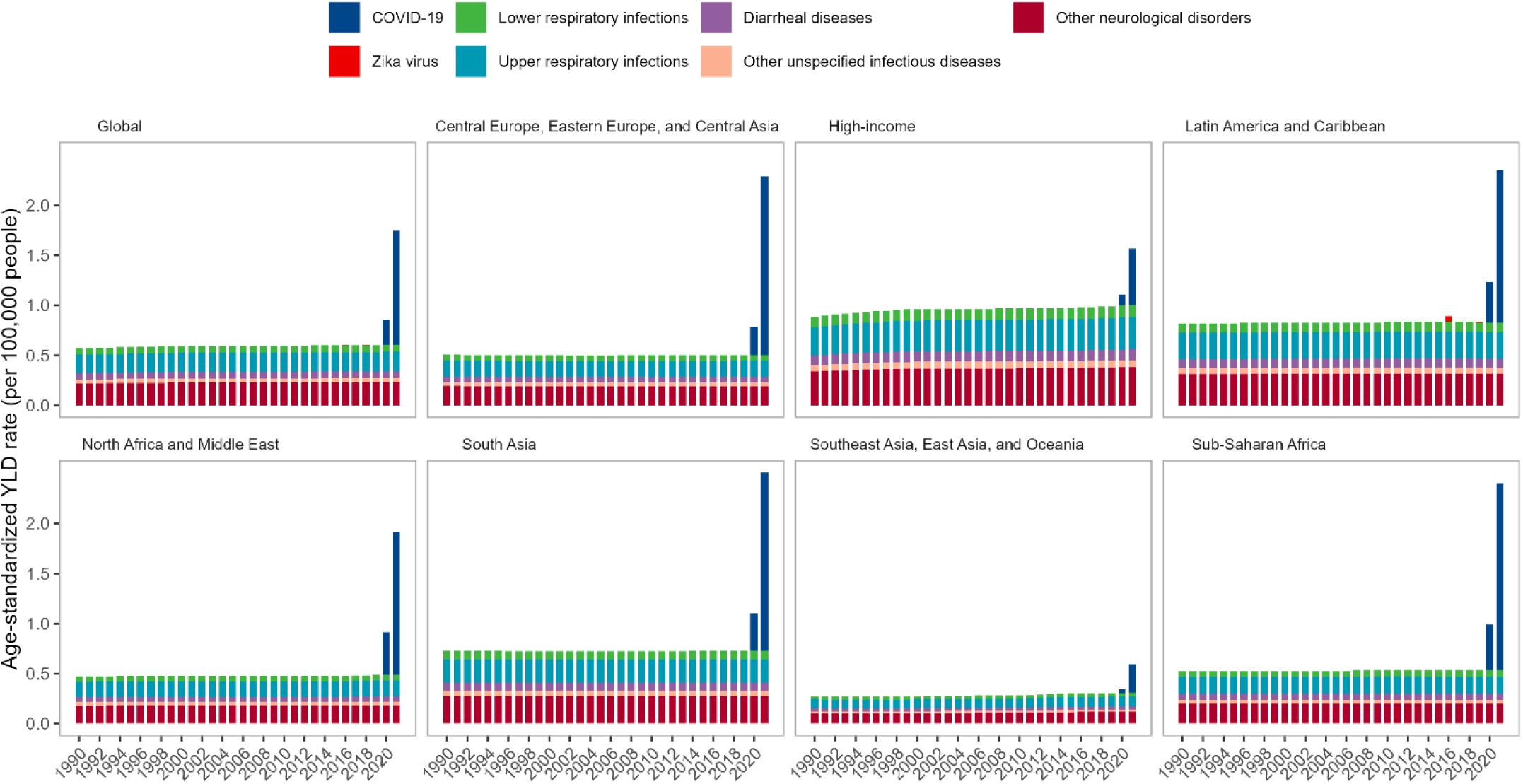
Age-standardized years lived with disability rates of Guillain-Barré syndrome attributed to underlying causes by sex, super region and years from 1990-2021.

### Impact of vaccination against COVID-19 on the GBS burden

We observed significantly negative correlations between COVID-19 vaccination coverage rates and both overall and COVID-19-specific age-standardized YLD rates of GBS in 2021 (**table 2 and supplementary figure S16-S17**). In the minimal sufficient GLIM model, each additional fully vaccinated person (per 100) significantly reduced all-cause age-standardized YLD rates by 0.0070 (per 100,000 people) and COVID-19-specific age-standardized YLD rates by 0.0128 (per 100,000 people). In the direct effect GLIM model, total vaccination also significantly lowered all-cause age-standardized YLD rate (β = -0.0039, p = 0.01) and COVID-19-specific age-standardized YLD rate (*β* = -0.0087, p < 0.001), suggesting that COVID-19 vaccines might exert a direct protective effect on the GBS burden in addition to preventing COVID-19 (**table 2**). **Figure 7** demonstrates smooth curves and estimated degrees of freedom (EDFs) derived from adjusted GAMs, suggesting non-linear relationships between vaccination coverage rates and age-standardized YLD rates of GBS. Stronger associations were observed for the countries with moderate vaccination coverage, while weaker associations were observed for the countries with lower or higher vaccination coverage.

**Table 2.** Association between COVID-19 vaccine coverage and age-standardized YLD rate at national level in 2021, using generalized linear model with Gaussian distribution and log-link function.

| Independent variable | Model | n | All-cause age-standardized YLD rate |  |  |  | Age-standardized YLD rate due to COVID-19 |  |  |  |
| --- | --- | --- | --- | --- | --- | --- | --- | --- | --- | --- |
| | | | $\beta$ | SE | t-value | p | $\beta$ | SE | t-value | p |
| No. of total vaccinations (per hundred)# | Comprehensive model* | 148 | -0.0033 | 0.0005 | -5.81 | <.001 | -0.0040 | 0.0009 | -4.56 | <.001 |
|  | Minimal sufficient model† | 161 | -0.0028 | 0.0007 | -4.24 | <.001 | -0.0052 | 0.0010 | -5.15 | <.001 |
|  | Direct effect model‡ | 161 | -0.0014 | 0.0005 | -2.6 | 0.01 | -0.0033 | 0.0007 | -4.42 | <.001 |
| No. of people vaccinated (per hundred)# | Comprehensive model* | 137 | -0.0066 | 0.0014 | -4.73 | <.001 | -0.0070 | 0.0022 | -3.13 | 0.002 |
|  | Minimal sufficient model† | 150 | -0.0064 | 0.0016 | -3.90 | <.001 | -0.0118 | 0.0024 | -4.87 | <.001 |
|  | Direct effect model‡ | 150 | -0.0032 | 0.0014 | -2.38 | 0.019 | -0.0078 | 0.0018 | -4.32 | <.001 |
| No. of people fully vaccinated (per hundred)# | Comprehensive model* | 143 | -0.0081 | 0.0015 | -5.60 | <.001 | -0.0090 | 0.0023 | -3.91 | <.001 |
|  | Minimal sufficient model† | 156 | -0.0070 | 0.0017 | -4.20 | <.001 | -0.0128 | 0.0025 | -5.08 | <.001 |
|  | Direct effect model‡ | 156 | -0.0039 | 0.0014 | -2.84 | 0.005 | -0.0087 | 0.0019 | -4.67 | <.001 |

$\beta$ =regression coefficient; COVID-19=coronavirus disease 2019; SE=standard error; YLD=years lived with disability.

**Figure 7.**
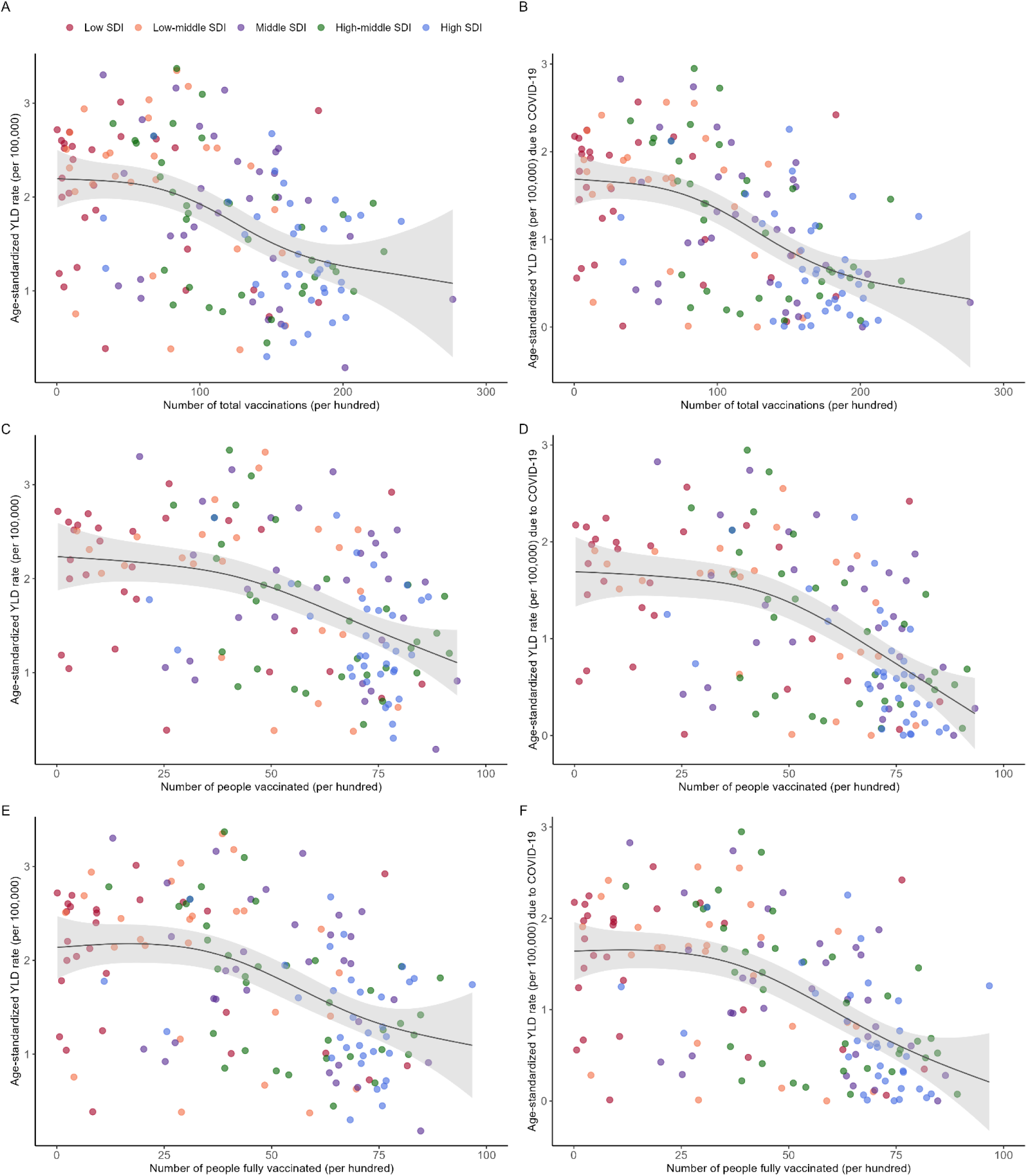
Number of people vaccinated per hundred by 31.12.2021 versus (A) all-cause age-standardized year lived with disability rate in 2021, and (B) age-standardized year lived with disability rate due to COVID-19 in 2021.

We also explored associations between the GBS burden and other potential influence factors, including GSI, and the GBS burden in the previous year (**supplementary figure S15-S20**). Results derived from a comprehensive GLIM showed that GSI, as well as SDI, was not statistically significantly associated with all-cause and COVID-19-specific GBS burden in 2021, adjusting for vaccination coverage rate by the end of 2021 and the GBS burden in 2020 (**supplementary table S5**).

## Discussion

This is the first study that investigated the association between the COVID-19 pandemic and vaccination coverage and the GBS burden globally. Our findings revealed a substantial increase in the global age-standardized YLDs related to GBS during the first two years of the COVID-19 pandemic, with SARS-CoV-2 infection being the primary contributor to this sharp rise in the GBS burden. We identified significant regional variations, noting that low SDI regions experienced the steepest increase in the GBS burden. Importantly, improving vaccination coverage was the most effective measure to mitigate the post-COVID-19 burden of GBS, despite the existing inequalities in vaccination rates associated with SDI. This study provides a crucial basis for understanding the global epidemiology of GBS during the COVID-19 pandemic and highlights that the benefits of vaccination outweigh the risks in controlling the post-COVID-19 GBS burden.

### The burden of GBS attributable to the SARS-CoV-2 infection

The association between SARS-CoV-2 infection and GBS has been a subject of debate. Our investigation provides novel evidence for the link between GBS and COVID-19 from a global perspective. The observed sharp rise in the burden of GBS during the first two years of the COVID-19 pandemic is consistent with several population-based studies.^27–29^ A cohort study conducted in the UK and Spain reported an increased risk of GBS associated with SARS-CoV-2 infection.^8^ Similarly, a self-controlled case series revealed an incidence rate ratio of 5.25 (95% confidence interval: 3.00 to 9.18) for GBS within 28 days following a positive SARS-CoV-2 test.^30^ Moreover, a recent case-control study from Israel reported an odds ratio of 6.30 (95% confidence interval: 2.55 to 15.56) for GBS associated with SARS-CoV-2 infection.^31^ However, some other observational studies did not observe an increased risk of GBS during the pandemic era, and some even suggested a reduction in GBS incidence compared to previous years.^32–35^ A recent meta-analysis revealed an increased risk of GBS in northern Italy during the early stages of the pandemic, while a slight reduction in GBS risk was observed in other countries during the pandemic.^36^ The case reports and cohort studies included in this meta-analysis primarily involved European countries and only analyzed hospitalized patients,^36^ which may lead to potential selection bias. This study highlights the importance of not only controlling infection transmission during a pandemic but also addressing the burden of diseases that arise as post-infection complications like GBS. By recognizing and addressing the broader spectrum of pandemic-related health challenges, we can better prepare for and mitigate the multifaceted impacts of future infectious disease outbreaks.

### Sex and age differences in the burden of GBS

The burden of GBS remained higher in males than in females across all age groups from 2019 to 2021, consistent with the pre-pandemic sex pattern.^1^ However, it is noteworthy that the GBS burden increased more rapidly in females than in males during this period, indicating a potential shift in the male-to-female ratio of GBS occurrence during the pandemic. Additionally, the global burden of GBS rose more rapidly among younger age groups of both sexes, particularly those aged 15–29 years. Studies have suggested that immune responses to SARS-CoV-2 infection differ between males and females and across various age groups.^37^ ^38^ These differences may contribute to the narrowed disparities between sexes and age groups in GBS following SARS-CoV-2 infection. Further research is needed to validate these changes in sex and age patterns and to explore the underlying causes.

### Regional difference in the burden of GBS

At the regional level, there is a clear variation in the burden of GBS, which is influenced by SDI. Specifically, regions with lower SDI levels experienced more rapid increases in the GBS burden during the first two years of the COVID-19 pandemic. Notably, compared to low SDI countries, high SDI countries experienced faster growth in the GBS burden from 1990 to 2019, as shown in this study and by others.^39^ This increase can be attributed to factors such as aging populations, better recognition and diagnostic techniques for GBS, and more healthcare resources.^39–41^ However, this trend contrasts with the pandemic period from 2019 to 2021, during which high SDI countries managed to decelerate the growth rate of the GBS burden. Indeed, countries with higher SDI, which typically have more robust healthcare systems and greater financial resources, were better equipped to promptly implement extensive measures to control the spread of SARS-CoV-2, thereby effectively managing the pandemic and, in turn, mitigating the associated rise in GBS cases.^42^ ^43^ In contrast, low SDI countries, with more limited healthcare and economic resources, faced substantial challenges in managing the pandemic. Resource constraints and the limited resilience of their healthcare system hindered their ability to control the spread of the virus, leading to a sharper rise in the GBS burden.^44–46^ Therefore, strengthening pandemic preparedness and response mechanisms in low SDI regions is essential to manage infectious disease outbreaks and associated health burdens more effectively. It also highlights the necessity for tailored response strategies that consider regional disparities in healthcare capacity and resource availability.

### Impact of vaccination on controlling the burden of GBS

Vaccination is an effective measure for controlling the transmission of SARS-CoV-2, with estimates suggesting that over 14 million deaths were prevented globally in just the first year of vaccination deployment.^11^ Our results revealed a negative correlation between the number of vaccinated individuals and the increase in the burden of GBS, suggesting that vaccination played a crucial role in controlling the GBS burden by mitigating the spread of SARS-CoV-2. However, significant disparities in vaccination coverage exist between countries. High-income countries have more immediate access to a variety of vaccines and higher vaccination rates, whereas low-income countries face substantial delays in vaccine rollout and consequently worse vaccination effects.^47^ ^48^ Research indicates that an additional 45% of COVID-19-related deaths could have been averted if low-income countries had met the 20% vaccination coverage target set by the COVID-19 Vaccines Global Access (COVAX).^11^ Moreover, a one-day delay in the first vaccination in low-income countries was associated with a 1.92% increase in cumulative cases compared to high-income countries.^49^ This inequality in vaccination coverage aligns with our findings that vaccination rates were positively correlated with SDI levels. The limited vaccination coverage in low SDI regions not only led to a surge in COVID-19 cases but also resulted in a faster growth burden of GBS as a post-infectious complication. Therefore, preventing and controlling SARS-CoV-2 infections through vaccination is an effective strategy to reduce the rise in GBS cases. Moreover, addressing the inequities in vaccine distribution and access is crucial to reducing the ongoing global burden of both COVID-19 and its associated post-infectious complications like GBS.

There were extensive concerns following the rollout of COVID-19 vaccines in the general public and the scientific community, because GBS can be triggered by certain vaccinations.^1^ ^50^ Previous research indicated that adenovirus-vectored vaccines might be associated with an increased risk of GBS,^30^ ^36^ ^51^ while mRNA vaccines were observed to have a lower risk of inducing GBS.^31^ ^36^ This difference may be attributed to the immune responses towards the viral vector through molecular mimicry. In contrast, mRNA vaccines lack the viral antigens that could induce such an autoimmune response.^52^ Our findings suggest that vaccination, regardless of the type, is effective in reducing the burden of GBS on a global scale. This protective effect persists even after accounting for SARS-CoV-2 infection, implying that vaccination might reduce the risk of GBS independently of its impact on COVID-19 infection. This raises the possibility that vaccination could inhibit autoimmune neurological diseases through immunological changes induced by the vaccine, as suggested by other vaccines.^53^ Future studies are needed to determine if COVID-19 vaccines have a protective effect against the development of GBS. Overall, these findings indicate that the benefits of vaccination significantly outweigh the risks in controlling the burden of GBS during the COVID-19 pandemic. Policymakers should advocate for transparent communication about the advantages and potential adverse effects of vaccination. This transparency ensures the public is well-informed and helps in countering vaccine hesitancy.

### Strengths and limitations of this study

This comprehensive analysis elaborates on the epidemiology of GBS before and during the COVID-19 pandemic at global, regional, and national levels, across all age groups and both sexes. Our findings highlight that the benefits of vaccination outweigh the risks in controlling the post-COVID-19 GBS burden. These insights are vital for health practice and policy-making, offering clear evidence for managing the GBS burden during future COVID-19 waves or other potential pandemics. However, this study has several potential limitations. First, variations in health information systems and reporting mechanisms across countries can lead to biases and incomplete data due to the use of predictive covariates. Using the same disability index for YLDs across all countries and age groups may not reflect the true burden of GBS, as recovery rates, disability levels, and survival vary with healthcare infrastructure, socioeconomic development, GBS variants, and patient demographics. Second, diagnosing GBS in resource-limited settings can be challenging, and diagnostic codes may not always be accurate. In severe COVID-19 cases, GBS might be misdiagnosed as critical illness polyneuropathy, affecting disease burden estimates. Third, the GBD database relies on data collected at the country level, which may lead to an ecological fallacy. This occurs when inferences about individuals are made based on aggregate data. Therefore, caution is needed when interpreting the results in real-world contexts. Nevertheless, our study has important implications for providing a comprehensive epidemiological profile of the GBS burden during the pandemic and highlighting the positive health effects of COVID-19 vaccination on the GBS burden.

## Conclusion

Predominantly driven by the surge of COVID-19 cases, the burden of GBS significantly increased during the first two years of the pandemic, especially in those countries with lower SDI levels. Females and younger populations (aged 15-29 years) experienced more rapid increases in the GBS burden. Despite significant disparities in vaccination coverage between countries, the benefits of vaccination outweighed the risks in managing the post-COVID-19 GBS burden. These insights can guide policymakers in planning targeted control measures to manage the burden of diseases that arise as post-infection complications, such as GBS, during future COVID-19 waves or other potential pandemics.

## Data Availability

The Global Burden of Disease (GBD) study is a comprehensive regional and global research program that assesses mortality and disability from major diseases, injuries, and risk factors.
Our World in Data is an online publication that presents empirical research and data on global development issues.

https://vizhub.healthdata.org/gbd-results/

https://ourworldindata.org/

## Contributors

XG and CZ contributed equally to this work and are joint first authors. SYZ, DSF, and ZKL contributed equally to this work and are joint senior authors. XG, CZ, DSF, and ZKL conceived and designed the study. SYZ provided supervision and mentorship. XG and CZ performed the methodology, statistical analysis and interpretation of data. JTY, ZMY, and JNF contributed to the acquisition and interpretation of data. XG, CZ, DSF, and ZKL drafted the manuscript. All authors revised the manuscript and approved the final version before submission. ZKL is the study guarantor and attests that all listed authors meet authorship criteria and that no others meeting the criteria have been omitted.

## Funding

This study was supported by the Postdoctoral Fellowship Program of China Postdoctoral Science Foundation (Grant No. GZC20240064), Young Scientists Fund of the National Natural Science Foundation of China (Grant No. 82204175), Key Program of the National Natural Science Foundation of China (Grant No. 82330107), National Natural Science Foundation of China (Grant No. 81873784 and 82071426), Funds for International Cooperation and Exchange of the National Natural Science Foundation of China (Grant No. 7231101396), Bill and Melinda Gates Foundation (Grant No. INV-035024), and Clinical Cohort Construction Program of Peking University Third Hospital (Grant No. BYSYDL2019002 and BYSYZD2021004). The funders of this study had no role in the study design, data collection, data analysis, data interpretation, writing of the report, or decision to submit the article for publication.

## Competing interests

All authors have completed the ICMJE uniform disclosure form at www.icmje.org/disclosure-of-interest/ and declare: support from the Key Program of the National Natural Science Foundation of China, National Natural Science Foundation of China, Young Scientists Fund of the National Natural Science Foundation of China, Funds for International Cooperation and Exchange of the National Natural Science Foundation of China, Postdoctoral Fellowship Program of China Postdoctoral Science Foundation, Bill and Melinda Gates Foundation, and Clinical Cohort Construction Program of Peking University Third Hospital for the submitted work; no financial relationships with any organizations that might have an interest in the submitted work in the previous three years; no other relationships or activities that could appear to have influenced the submitted work.

## Ethical approval

Not required as this study used secondary data aggregated at both country and global levels.

## Data sharing

The data used for analyses are publicly available at https://ghdx.healthdata.org/ gbd-results-tool, https://ourworldindata.org/.

## Transparency

The lead authors (ZKL and DSF) affirm that the manuscript is an honest, accurate, and transparent account of the study being reported; that no important aspects of the study have been omitted; and that any discrepancies from the study as planned (and, if relevant, registered) have been explained.

## Dissemination to participants and related patient and public communities

The research findings will be disseminated to the wider community through press releases, social media platforms, presentations at international forums, and reports to relevant government agencies and academic societies.

## Acknowledgment

The authors would like to thank the teams behind the Global Burden of Disease Study 2021 and Our World in Data for their invaluable contributions to making comprehensive and high-quality data available for our research. Their efforts in data collection, curation, and dissemination have been instrumental in enabling this study. We also extend our gratitude to all healthcare professionals, researchers, and volunteers who have worked tirelessly during the COVID-19 pandemic, providing the essential data and support that made this study possible.

## Notes

### Competing Interest Statement

The authors have declared no competing interest.

### Author Declarations

The Global Burden of Disease (GBD) study adhered to the Guidelines for Accurate and Transparent Health Estimates Reporting (GATHER) statement 22 and obtained ethical clearance from the Institutional Review Board of the University of Washington (study 9060). Our World in Data (OWID) used publicly available data from reliable sources, including the World Health Organization (WHO), the National Ministry of Health, and the Centers for Disease Control and Prevention (CDC). Secondary analysis of the de-identified data from GBD and OWID did not require additional ethical approval.

