## Supplementary table S1 for "Impact of the COVID-19 pandemic and vaccination on global, regional, and national burden of Guillain-Barré syndrome: observational study"

Supplementary information: Additional methods, results and discussions, tables, and figures

1. Supplementary methods

1.1. Overview of data sources

Global Burden of Diseases, Injuries, and Risk Factors Study (GBD) 2021

GBD 2021, led by the Institute for Health Metrics and Evaluation (IHME), is an extensive research initiative that provides comprehensive and comparable data on health loss worldwide. It utilized a vast array of data sources to ensure accurate and representative estimates, including: vital registration systems, household surveys, hospital and clinical records, disease registries, published studies, and population and housing censuses, to quantify the prevalence, incidence, mortality, and disability of 371 diseases and injuries, and 88 risk factors.^1-3^ Disability-adjusted life years (DALYs), years of life lost (YLLs), and years lived with disability (YLDs) were employed to offer insights into the burden of diseases and injuries and inform policy decisions.

Impairment in the GBD 2021 study refers to the functional limitations and restrictions caused by various health conditions, impacting an individual's ability to perform everyday activities. Impairment is a critical component of the study as it helps quantify the non-fatal health outcomes contributing to the overall disease burden.^1^ Guillain-Barré syndrome (GBS) is an acute, autoimmune disorder that affects the peripheral nervous system, often resulting in muscle weakness, paralysis, and sensory disturbances. It is considered an impairment in GBD 2021 because it leads to significant functional limitations and disabilities, contributing to YLDs.

Our World in Data (OWID)

OWID is an online publication that provides accessible and comprehensive data on a wide range of global issues, including health, education, environment, and economic development. It aims to present research and data in a clear, easily understandable format through interactive charts, maps, and extensive articles. The platform sources data from reputable organizations such as the World Health Organization (WHO), World Bank, and United Nations, making it a valuable resource for researchers, policymakers, and the public to understand and address global challenges. OWID provides extensive coverage and analysis of the global vaccination campaign and non-pharmacological interventions against COVID-19.^4 5^

Data from two sources were merged by standardizing country names.

1.2. Measures

Prevalence and YLD were used to measure the burden of GBS in this study. Point prevalence is provided by IHME and refers to the number of cases (or individuals) in a population at a specific point in time who have a particular health condition. YLD is calculated by multiplying the prevalence by the disability weight that are derived through surveys and expert consensus to reflect the impact of the condition on quality of life. In this study, numbers (per thousand) and age-standardized rates (ASRs, per 100,000 people) were expressed.

Socio-demographic Index (SDI) is a composite measure reflecting a region’s socio-economic development. It is calculated using three components: a country’s lag-distributed income per capita, average years of schooling, and the total fertility rate in females younger than age 25 years of all areas in GBD 2021. SDI ranges between 0 and 1, with higher values indicating greater development.^6^

In OWID, the metric vaccinated people per hundred is used to quantify the proportion of a population that has received COVID-19 vaccinations.^5^ This metric is often broken down into various categories, such as the percentage of people who have received at least one dose of a COVID-19 vaccine, those who have been fully vaccinated, and sometimes those who have received additional doses (booster shots).

The Government Stringency Index (GSI) is a composite measure developed by the Oxford COVID-19 Government Response Tracker (OxCGRT) to quantify the stringency of government responses to the COVID-19 pandemic.^4^ It provides a way to compare the severity and strictness of policies across different countries and regions over time. The index is calculated based on nine policy indicators, including school closures, workplace closures, cancellation of public events, restrictions on public gatherings, closures of public transport, stay-at-home requirements, public information campaigns, restrictions on internal movements, and international travel controls. Each indicator is scored on an ordinal scale, reflecting the level of strictness of the measure: no measures, recommend closing, require closing (some levels or categories, e.g., just high schools, or just public schools), and require closing all levels. The final stringency score is the unweighted average of the normalized scores for the nine indicators, adjusted as needed.

1.3. Uncertainty analysis

The IHME accounted for uncertainty in all calculations by sampling 1,000 draws at each step. Final estimates were derived using the mean of these 1,000 draws, with the 95% uncertainty intervals (UIs) defined as the values at the 25th and 75th positions of the ordered draws. Estimates with a 95% UI that did not include zero were considered statistically significant

1.4. Statistical analysis

Temporal trend analysis

Both estimated annual percentage change (EAPC) and joinpoint regression were used to describe trends of GBS burden over time in this study. EAPC presents the year-over-year change, providing a more immediate measure of change for each year.^7^ We employed a linear regression model, assuming a linear relationship: ln (ASR) = α + β*(calendar year) + ε. β denotes the positive or negative trend in the age-standardized rate, and ε accounts for the random deviation. EAPC was calculated using a log-linear regression model where the natural logarithm of the rate was regressed on time (year). The formula is EAPC = 100 × (exp(β) − 1). The 95% confidence interval (CI) was obtained from the linear model.

Joinpoint regression is a statistical technique used to identify points where a significant change in the trend of a data series occurs.^8^ These points are referred to as "joinpoints". To detect the "joinpoints", the analysis starts with a null model (no joinpoints, a single linear trend). Then, additional joinpoints are added iteratively, and their significance is tested using. The best model was selected using the Weighted Bayesian Information Criterion (WBIC) method, which is the most flexible method in adapting across a wide range of situations compared to the permutation test, BIC, and BIC3.^9^ Annual percentage change (APC) was calculated using the slope of the segment in the log-linear model. In this study, joinpoint regression was performed using the Joinpoint Regression Program released in June 2024 (version 5.2.0).

Generalized additive model

Generalized Additive Models (GAMs) are a class of statistical models that extend generalized linear models (GLMs) by allowing for non-linear relationships between the predictors and the response variable. The key idea behind GAMs is to model the expected value of the response variable as an additive function of smooth functions of the predictor variables. This flexibility allows GAMs to capture complex patterns in the data that linear models might miss. In a GAM, the linear predictor is an additive combination of smooth functions of the predictors: *g*(*E*(*Y*)) = *ꞵ*_0_+*s*_1_(*X*_1_)+*s*_2_(*X*_2_)+…+*s*_p_(*X*_p_).

- *g* is the link function.
- *E*(*Y*) is the expected value of the response variable.
- *β*_0_ is the intercept.
- *s*_i_(*X*_i_) are the smooth functions of the predictors *X*_i_ .

Missing data handling

We extracted vaccination rate data on December 31, 2021 for analyzing the potential impact of vaccination against COVID-19 on GBS burden. However, vaccination rate data might not be available for this day in some countries and territories. Therefore, we fill the data with the nearest values within a window of 5 days before and after the missing point date. Observations (i.e. countries and territories) were excluded from analyses if the missing point could not be filled or the countries and territories could not be found in the WOID database.

2. Supplementary Results and discussions

Prevalence of GBS

Supplementary table S1 presents data on the prevalent cases of GBS and the age-standardized rates (ASRs) per 100,000 people for the years 2020 and 2021, alongside the EAPC in ASRs from 1990 to 2019 and from 2019 to 2021. There was a substantial increase in the prevalence and ASRs of GBS globally during the COVID-19 pandemic period (2019-2021) compared to the long-term trends from 1990 to 2019. Regions with lower SDI levels showed higher relative increases in ASRs and prevalent cases compared to higher SDI regions. Significant regional variability in EAPC suggests differing impacts of the COVID-19 pandemic and other factors on GBS prevalence. Overall, the spatial distribution and trend of GBS prevalence were similar with GBS YLD.

Factors that potentially influence GBS burden during the COVID-19 pandemic

***COVID-19*:** For most countries and territories, the increase in GBS burden was mainly attributed to COVID-19 during the pandemic. We observed a strong association between age-standardized COVID-19 incidence and the ASYR of GBS **(supplementary figure S19-S20**).

***Socio-demographic index (SDI)*:** The relationship between the SDI and the burden of GBS was statistically significant, highlighting disparities in GBS incidence across different socio-economic contexts (**table 1**). In 2021, regions with lower SDI showed markedly higher ASYR for GBS, with low SDI regions having an ASYR of 2.37 per 100,000 people and low-middle SDI regions at 2.32 per 100,000 people. Conversely, high SDI regions exhibited lower ASYR values, with an ASYR of 1.48 per 100,000 people. This trend indicates that lower SDI regions bear a heavier burden of GBS, likely due to a higher COVID-19 incidence. Therefore, we investigated and observed a negative association between COVID-19 incidence and SDI levels (**supplementary figure S13**).

We further explored the relationship of SDI with vaccination coverage rate and GSI, which were utilized to control the pandemic. The total number of vaccinations (per hundred) was significantly associated with SDI levels (**supplementary figure S14**). **Supplementary figure S15** shows a moderate positive correlation (R = 0.38, *p* < 0.001), indicating that regions with higher SDI tended to implement stricter governmental measures in response to COVID-19.

These findings suggest that higher socio-demographic index countries might have more resources and organizational capabilities to promote public health measures during the pandemic. Therefore, we proposed a hypothesis that COVID-19 incidence, vaccination coverage, and non-pharmacological interventions are mediators in the link between GBS burden and SDI (**supplementary figure S1**).

Generalized linear models were applied to further assess the link between SDI and GBS burden adjusting for potential factors. **Supplementary table S5** shows that SDI levels were not statistically significantly associated with all-cause and COVID-19-specific GBS burden in 2021, after adjusting for vaccination coverage rate by the end of 2021 and GBS burden in 2020.

***Vaccination against COVI-19:*** Vaccination coverage rates by December 31, 2021 were statistically significantly associated with the age-standardized COVID-19 incidence in 2021, highlighting the effectiveness of vaccines in reducing the transmission of the disease within the population (**supplementary figure S22-S23**). The link between vaccination coverage and GBS burden has also been assessed and described with details in the main text. Detailed vaccination coverage rates of 204 countries and territories is shown in **supplementary table S6.**

***Non-pharmacological interventions (NPI):*** However, we did not observe significant association between GSI and GBS burden in 2020, the year in which COVID-19 vaccination campaign was not initiated (**supplementary figure S19-S21**). However, it has been shown in other studies that non-pharmacological measures, including precautions, air and droplet precautions, mask wearing, adequate room ventilation, and air treatment, were effective in preventing the occurrence of COVID-19.^10^ The attenuated association observed in our study could be explained by the “lag effect” of public health policy on the burden of COVID-19 and GBS. Usually, policy measures such as lockdowns or social distancing rules do not immediately affect infection rates. These measures require time to influence human behaviors and community transmission dynamics. Typically, the impact of policy measures may become apparent over several weeks, particularly as compliance with these measures increases.^11 12^

Other factors related to government policies such as the compliance with governmental restrictions, may influence the implementation of the policy.^13^ These factors which could affect the link between GSI and GBS burden, were not be obtained.

References

1. Global Burden of Disease Collaborative Network. Global Burden of Disease Study 2021 (GBD 2021) Results. Seattle, United States: Institute for Health Metrics and Evaluation (IHME), 2022.

Supplementary Tables

| **Supplementary Table S1** Prevalent cases for Guillain-Barré syndrome in 2020 and 2021 and the estimated annual percentage change in the age-standardised rates per 100,000, by GBD region and SDI, from 1990 to 2021 | | | | | | | |
| --- | --- | --- | --- | --- | --- | --- | --- |
|  |  | **2020** | | **2021** | | **2019–2021** | **1990–2019** |
| **GBD supper regions** | **GBD regions** | **Count  (thousands)** | **ASR of prevalence (per 100,000 people)** | **Count  (thousands)** | **ASR of prevalence (per 100,000 people)** | **EAPC** | **EAPC** |
| Global |  | 230.1 (194.3 to 269.7) | 2.89 (2.45 to 3.4) | 471.9 (389.2 to 554.1) | 5.91 (4.87 to 6.97) | 70.35 (38.73 to 109.17) | 0.16 (0.14 to 0.18) |
| Low SDI |  | 33.3 (27.3 to 39.9) | 3.39 (2.79 to 4.03) | 82.4 (65.1 to 99.3) | 8.03 (6.39 to 9.6) | 103.38 (71.09 to 141.77) | -0.01 (-0.02 to 0) |
| Low-middle SDI |  | 64.4 (53.4 to 77.4) | 3.54 (2.94 to 4.21) | 147.3 (118.9 to 177.6) | 7.84 (6.36 to 9.36) | 88.25 (56.64 to 126.25) | 0.02 (0.01 to 0.03) |
| Middle SDI |  | 60.7 (50.8 to 71.7) | 2.46 (2.07 to 2.93) | 124.5 (102.5 to 147) | 5.02 (4.11 to 5.98) | 68.69 (36.08 to 109.13) | 0.42 (0.4 to 0.44) |
| High-middle SDI |  | 25.5 (21.2 to 30.5) | 1.79 (1.49 to 2.15) | 53.5 (44.4 to 62.3) | 3.82 (3.16 to 4.56) | 67.04 (26.52 to 120.54) | 0.11 (0.09 to 0.14) |
| High SDI |  | 46 (39.2 to 53.5) | 3.45 (2.95 to 4.04) | 63.8 (55.4 to 73.6) | 4.99 (4.29 to 5.78) | 28.02 (11.45 to 47.05) | 0.11 (0.06 to 0.16) |
| Central Europe, eastern Europe, and central Asia |  | 12 (10 to 14.5) | 2.65 (2.19 to 3.17) | 35 (28.7 to 42) | 7.74 (6.31 to 9.33) | 113.4 (49.75 to 204.11) | -0.01 (-0.02 to 0) |
|  | Central Asia | 2.8 (2.2 to 3.4) | 3.04 (2.43 to 3.65) | 6.6 (5.1 to 8.3) | 6.98 (5.39 to 8.72) | 94.95 (61.83 to 134.84) | 0.01 (0 to 0.01) |
|  | Central Europe | 2.5 (2.1 to 3.1) | 1.92 (1.61 to 2.31) | 9.3 (7.6 to 11) | 7.24 (5.83 to 8.66) | 129.13 (30.68 to 301.74) | -0.14 (-0.18 to -0.1) |
|  | Eastern Europe | 6.7 (5.5 to 8.1) | 2.89 (2.39 to 3.46) | 19.1 (15.4 to 23.1) | 8.43 (6.75 to 10.21) | 114.03 (51.08 to 203.22) | 0.01 (0.01 to 0.01) |
| High-income |  | 50.1 (42.8 to 58.5) | 3.75 (3.21 to 4.39) | 67.8 (58.8 to 78) | 5.29 (4.56 to 6.13) | 25.98 (10.67 to 43.42) | 0.28 (0.22 to 0.33) |
|  | Australasia | 0.6 (0.4 to 0.7) | 1.59 (1.28 to 1.97) | 0.6 (0.5 to 0.7) | 1.62 (1.31 to 2.00) | 1.52 (1.01 to 2.04) | 1.24 (1.02 to 1.46) |
|  | High-income Asia Pacific | 11 (9.2 to 13.2) | 6.03 (4.97 to 7.40) | 11.6 (9.8 to 13.8) | 6.34 (5.28 to 7.74) | 2.84 (0.31 to 5.43) | 0.15 (0.1 to 0.2) |
|  | High-income North America | 22.6 (19.6 to 26.3) | 4.67 (4.05 to 5.40) | 31.5 (27.5 to 35.8) | 6.93 (5.96 to 7.90) | 30.43 (12.72 to 50.92) | 0.45 (0.33 to 0.57) |
|  | Southern Latin America | 3 (2.5 to 3.6) | 4.23 (3.58 to 5.09) | 4.3 (3.6 to 5.2) | 6.05 (5.02 to 7.31) | 25.64 (8.46 to 45.55) | 0.24 (0.16 to 0.32) |
|  | Western Europe | 12.9 (10.8 to 15.3) | 2.33 (1.97 to 2.74) | 19.8 (16.9 to 23.1) | 3.81 (3.21 to 4.46) | 42.25 (21.44 to 66.63) | 0.71 (0.66 to 0.76) |
| Latin America and Caribbean |  | 24.6 (21 to 28.7) | 4.15 (3.56 to 4.86) | 47.7 (39.5 to 56.5) | 7.93 (6.57 to 9.44) | 67.99 (45.29 to 94.25) | 0.11 (0.06 to 0.16) |
|  | Andean Latin America | 3 (2.5 to 3.6) | 4.76 (3.96 to 5.61) | 5.9 (4.6 to 7.3) | 8.94 (7.05 to 11.05) | 86.76 (85.6 to 87.93) | -0.17 (-0.21 to -0.13) |
|  | Caribbean | 1.2 (1 to 1.5) | 2.58 (2.06 to 3.15) | 1.9 (1.5 to 2.3) | 3.88 (3.03 to 4.82) | 36.61 (22.32 to 52.59) | 0.02 (-0.1 to 0.15) |
|  | Tropical Latin America | 6.4 (5.4 to 7.5) | 2.71 (2.27 to 3.18) | 16 (13 to 19.4) | 6.76 (5.48 to 8.21) | 124.12 (98.15 to 153.5) | 0.04 (-0.04 to 0.12) |
|  | Central Latin America | 13.9 (11.8 to 16.4) | 5.60 (4.75 to 6.63) | 24 (20.1 to 28.1) | 9.49 (7.97 to 11.13) | 48.25 (27.37 to 72.55) | 0.07 (0.03 to 0.1) |
| North Africa and Middle East | North Africa and Middle East | 18.1 (14.4 to 22.4) | 3.09 (2.47 to 3.77) | 38.9 (30.7 to 47.9) | 6.47 (5.13 to 7.94) | 98.72 (87.25 to 110.9) | 0.04 (0.02 to 0.06) |
| South Asia | South Asia | 66.1 (55.6 to 79.1) | 3.73 (3.14 to 4.44) | 155.1 (126.7 to 185.3) | 8.48 (6.93 to 10.11) | 85.92 (47.92 to 133.67) | 0 (0 to 0) |
| Southeast Asia, east Asia, and Oceania |  | 25.6 (20.1 to 31.7) | 1.15 (0.92 to 1.44) | 43.2 (35.7 to 51.5) | 2.01 (1.66 to 2.40) | 39.62 (8 to 80.49) | 0.44 (0.37 to 0.52) |
|  | Southeast Asia | 15.4 (12.4 to 18.9) | 2.22 (1.80 to 2.71) | 32.9 (27.2 to 39) | 4.69 (3.89 to 5.55) | 55.91 (10.42 to 120.12) | 0.09 (0.07 to 0.11) |
|  | East Asia | 10 (7.5 to 12.9) | 0.64 (0.50 to 0.83) | 9.9 (7.4 to 12.9) | 0.63 (0.48 to 0.82) | 0.39 (-2.78 to 3.66) | 0.32 (0.13 to 0.5) |
|  | Oceania | 0.2 (0.1 to 0.2) | 1.31 (1.03 to 1.65) | 0.4 (0.3 to 0.6) | 3.21 (2.22 to 4.54) | 59.32 (-1.75 to 158.37) | 0 (0 to 0) |
| Sub-Saharan Africa |  | 33.5 (27.4 to 40.5) | 3.36 (2.77 to 3.99) | 84.1 (66.9 to 101.4) | 8.13 (6.56 to 9.72) | 112.86 (84.03 to 146.21) | 0.02 (0.01 to 0.02) |
|  | Western Sub-Saharan Africa | 16.8 (13.6 to 20.3) | 3.96 (3.28 to 4.69) | 34.2 (27 to 41.3) | 7.79 (6.18 to 9.38) | 100.98 (96.14 to 105.94) | -0.01  (-0.02 to -0.01) |
|  | Eastern Sub-Saharan Africa | 9.8 (8 to 12.1) | 2.60 (2.14 to 3.12) | 32.6 (26.1 to 39.4) | 8.33 (6.78 to 10.04) | 129.58 (57.49 to 234.67) | 0.09 (0.07 to 0.11) |
|  | Central Sub-Saharan Africa | 4.7 (3.6 to 5.8) | 3.92 (3.00 to 4.80) | 11.1 (7.9 to 13.9) | 8.97 (6.38 to 11.12) | 121.29 (113.26 to 129.62) | -0.01 (-0.01 to -0.01) |
|  | Southern Sub-Saharan Africa | 2.2 (1.8 to 2.7) | 2.87 (2.37 to 3.46) | 6.2 (4.9 to 7.5) | 7.82 (6.24 to 9.42) | 99.29 (39.74 to 184.23) | 0 (0 to 0.01) |

Regions are grouped by GBD super-region and alphabetically ordered. ASR=age-standardized rate. CI=confidence interval. EAPC=estimated annual percentage change. GBD=Global Burden of Diseases, Injuries and Risk Factors Study. SDI=Socio-demographic index. UI=uncertainty interval.

| **Supplementary Table S2** Years lived with disability of Guillain–Barré syndrome in 2020 and 2021 and the estimated annual percentage change in the age-standardised rates per 100,000 in 204 countries and territories, from 1990 to 2021 | | | | |
| --- | --- | --- | --- | --- |
| **Country and territory** | **ASR in 2020 (per 100,000)** | **ASR in 2021 (per 100,000)** | **2019-2021  EAPC in ASR** | **1990-2019**  **EAPC in ASR** |
| Afghanistan | 1.1 (0.7 to 1.7) | 2.4 (1.2 to 3.8) | 125.19 (112.92 to 138.15) | 0 (-0.01 to 0) |
| Albania | 0.8 (0.5 to 1.2) | 3.2 (1.8 to 4.8) | 173.72 (78.89 to 318.8) | 0 (0 to 0) |
| Algeria | 0.6 (0.4 to 0.9) | 0.9 (0.5 to 1.4) | 36.64 (29.23 to 44.49) | 0 (0 to 0) |
| American Samoa | 0.4 (0.2 to 0.6) | 0.4 (0.3 to 0.7) | 9.34 (-0.11 to 19.67) | 0.1 (0 to 0.2) |
| Andorra | 0.9 (0.6 to 1.4) | 1.8 (1.1 to 2.6) | 83.86 (73.48 to 94.86) | -0.02 (-0.03 to -0.02) |
| Angola | 0.9 (0.5 to 1.3) | 2.8 (1.4 to 4.2) | 126.52 (52.64 to 236.15) | -0.03 (-0.03 to -0.03) |
| Antigua and Barbuda | 0.7 (0.4 to 1) | 1 (0.6 to 1.4) | 23.35 (2.55 to 48.36) | 0.22 (-0.03 to 0.48) |
| Argentina | 1.4 (0.9 to 2) | 2 (1.2 to 2.9) | 24.91 (7.34 to 45.35) | 0.2 (0.09 to 0.31) |
| Armenia | 1.1 (0.7 to 1.6) | 2.8 (1.6 to 4.2) | 127.58 (96.16 to 164.03) | 0.01 (0.01 to 0.01) |
| Australia | 0.4 (0.3 to 0.7) | 0.4 (0.3 to 0.7) | 1.88 (1.2 to 2.57) | 1.76 (1.45 to 2.06) |
| Austria | 1.1 (0.8 to 1.6) | 1.7 (1 to 2.4) | 24.49 (4.54 to 48.25) | 1.2 (1 to 1.41) |
| Azerbaijan | 0.8 (0.5 to 1.2) | 2.6 (1.4 to 4.2) | 120.43 (38.68 to 250.37) | 0.02 (0.02 to 0.03) |
| Bahamas | 0.7 (0.5 to 1.1) | 1.2 (0.7 to 1.9) | 39.63 (15.98 to 68.11) | 0.05 (0.01 to 0.08) |
| Bahrain | 0.8 (0.5 to 1.1) | 1.9 (1.1 to 2.9) | 101.44 (55.26 to 161.36) | 0.01 (0.01 to 0.01) |
| Bangladesh | 1.1 (0.7 to 1.7) | 2.5 (1.4 to 3.8) | 94 (64.24 to 129.16) | 0 (0 to 0) |
| Barbados | 0.6 (0.4 to 0.9) | 0.8 (0.5 to 1.2) | 14.55 (0.83 to 30.14) | 0.09 (-0.01 to 0.19) |
| Belarus | 0.8 (0.5 to 1.2) | 1.9 (0.9 to 3.4) | 95.75 (53.3 to 149.97) | 0 (0 to 0.01) |
| Belgium | 0.8 (0.5 to 1.1) | 1.3 (0.8 to 1.9) | 57.19 (44.18 to 71.38) | 0.01 (0.01 to 0.01) |
| Belize | 0.7 (0.5 to 1.1) | 1.5 (0.8 to 2.5) | 55.17 (11.69 to 115.57) | 0.22 (0.02 to 0.41) |
| Benin | 0.7 (0.4 to 1.1) | 1.2 (0.7 to 2.1) | 51.73 (27.75 to 80.21) | 0 (-0.01 to 0) |
| Bermuda | 0.7 (0.4 to 1) | 0.9 (0.6 to 1.3) | 20 (3.78 to 38.74) | 0.01 (0.01 to 0.01) |
| Bhutan | 0.7 (0.4 to 1) | 0.7 (0.5 to 1.1) | 4.64 (1.02 to 8.4) | -0.01 (-0.01 to -0.01) |
| Bolivia (Plurinational State of) | 1.6 (1 to 2.4) | 3.3 (2.1 to 4.9) | 104.91 (101.93 to 107.94) | 0.02 (0.02 to 0.03) |
| Bosnia and Herzegovina | 0.6 (0.4 to 0.9) | 2.6 (1.5 to 3.9) | 177.06 (71.06 to 348.74) | -0.25 (-0.39 to -0.1) |
| Botswana | 0.6 (0.4 to 0.9) | 1.9 (1 to 3.1) | 86.76 (-3.52 to 261.53) | 0 (0 to 0.01) |
| Brazil | 0.8 (0.5 to 1.2) | 2 (1.3 to 2.9) | 123.7 (100.57 to 149.51) | 0.04 (-0.04 to 0.12) |
| Brunei Darussalam | 1.7 (1.1 to 2.6) | 1.8 (1.1 to 2.7) | 2.15 (0.5 to 3.84) | -0.01 (-0.02 to 0) |
| Bulgaria | 0.6 (0.4 to 0.9) | 2.6 (1.3 to 4.1) | 147.58 (31.63 to 365.68) | 0 (-0.01 to 0) |
| Burkina Faso | 1 (0.6 to 1.6) | 2.6 (1.5 to 3.9) | 117.65 (81.37 to 161.19) | 0 (0 to 0) |
| Burundi | 0.5 (0.3 to 0.7) | 0.9 (0.6 to 1.3) | 39.16 (6.52 to 81.79) | 0.03 (0.02 to 0.03) |
| Cabo Verde | 0.9 (0.5 to 1.4) | 2.2 (1.2 to 3.6) | 102.9 (63.26 to 152.16) | 0.14 (-0.04 to 0.33) |
| Cambodia | 0.5 (0.3 to 0.8) | 0.9 (0.5 to 1.3) | 28.91 (-3.08 to 71.47) | 0.01 (0.01 to 0.01) |
| Cameroon | 0.9 (0.5 to 1.5) | 2 (0.6 to 3.5) | 91.57 (72.18 to 113.14) | 0 (0 to 0) |
| Canada | 0.9 (0.6 to 1.4) | 1.2 (0.8 to 1.7) | 32.57 (29.05 to 36.19) | 0.84 (0.68 to 0.99) |
| Central African Republic | 1.2 (0.7 to 2.1) | 1.8 (1.1 to 3) | 83.87 (42.06 to 137.99) | -0.01 (-0.01 to -0.01) |
| Chad | 1.2 (0.7 to 2.2) | 2 (1.1 to 3.4) | 89.07 (56.56 to 128.33) | 0.03 (0.03 to 0.03) |
| Chile | 1 (0.7 to 1.4) | 1.4 (0.9 to 2.1) | 27.78 (14.59 to 42.49) | 0.58 (0.49 to 0.67) |
| China | 0.2 (0.1 to 0.3) | 0.2 (0.1 to 0.3) | 0.33 (-3.03 to 3.79) | 0.31 (0.11 to 0.52) |
| Colombia | 1.4 (0.9 to 2) | 2.4 (1.5 to 3.5) | 43.8 (15.69 to 78.74) | 0.05 (-0.02 to 0.11) |
| Comoros | 0.6 (0.4 to 1) | 2.6 (1.6 to 3.8) | 141.35 (28.03 to 354.94) | 0 (0 to 0) |
| Congo | 1.1 (0.7 to 1.6) | 2.1 (1.3 to 3) | 94.46 (94.03 to 94.9) | 0.02 (0.01 to 0.02) |
| Cook Islands | 0.4 (0.3 to 0.6) | 0.7 (0.4 to 1) | 36.39 (12.7 to 65.06) | -0.01 (-0.01 to -0.01) |
| Costa Rica | 1.3 (0.8 to 1.9) | 2.3 (1.4 to 3.4) | 39.96 (9.69 to 78.6) | 0 (-0.02 to 0.03) |
| Côte d'Ivoire | 1.2 (0.7 to 1.8) | 2.1 (1.2 to 3.2) | 96.95 (77.23 to 118.86) | 0 (0 to 0) |
| Croatia | 0.5 (0.3 to 0.8) | 1.9 (1.2 to 2.9) | 115.08 (19.54 to 286.95) | 0.02 (0.02 to 0.02) |
| Cuba | 0.6 (0.4 to 0.9) | 0.9 (0.6 to 1.3) | 19.98 (-1.29 to 45.85) | 0.07 (0.01 to 0.13) |
| Cyprus | 0.5 (0.3 to 0.8) | 0.9 (0.6 to 1.2) | 28.55 (0.46 to 64.49) | 0 (-0.01 to 0) |
| Czechia | 0.6 (0.4 to 0.9) | 2.7 (1.6 to 4) | 152.23 (34.42 to 373.27) | 0.02 (0.02 to 0.02) |
| Democratic People's Republic of Korea | 0.2 (0.2 to 0.4) | 0.2 (0.2 to 0.4) | 1.1 (0.79 to 1.41) | 0.01 (0.01 to 0.01) |
| Democratic Republic of the Congo | 1.3 (0.7 to 1.9) | 2.7 (1.6 to 4) | 123.73 (115.05 to 132.76) | -0.01 (-0.01 to -0.01) |
| Denmark | 0.5 (0.3 to 0.7) | 0.7 (0.5 to 1.1) | 35.96 (14.86 to 60.94) | 1.43 (1.03 to 1.82) |
| Djibouti | 1 (0.6 to 1.6) | 1.8 (1 to 2.8) | 96.64 (73.3 to 123.11) | 0.02 (0.02 to 0.02) |
| Dominica | 0.6 (0.4 to 1) | 0.9 (0.5 to 1.2) | 16.16 (0.07 to 34.84) | 0.4 (-0.09 to 0.88) |
| Dominican Republic | 0.9 (0.6 to 1.4) | 1.4 (0.8 to 2.3) | 51.61 (45.93 to 57.51) | 0.11 (-0.03 to 0.24) |
| Ecuador | 1.3 (0.8 to 1.9) | 2.5 (1.5 to 3.8) | 97.74 (97.64 to 97.83) | -0.79 (-0.89 to -0.68) |
| Egypt | 1.2 (0.5 to 1.9) | 2.2 (0.7 to 3.6) | 113.19 (81.67 to 150.17) | 0 (0 to 0) |
| El Salvador | 1.2 (0.8 to 1.8) | 1.9 (1.1 to 2.8) | 33.07 (13.34 to 56.23) | 0.23 (0.12 to 0.34) |
| Equatorial Guinea | 1.6 (1 to 2.4) | 1.9 (1.1 to 2.9) | 87.68 (15.75 to 204.29) | 0.04 (0.04 to 0.05) |
| Eritrea | 0.5 (0.3 to 0.7) | 1 (0.6 to 1.5) | 50.3 (0.64 to 124.45) | 0.01 (0.01 to 0.01) |
| Estonia | 0.5 (0.3 to 0.8) | 1.2 (0.8 to 2) | 57.73 (1.07 to 146.13) | 0.01 (0.01 to 0.01) |
| Eswatini | 0.8 (0.5 to 1.1) | 2.3 (1.2 to 4) | 107.05 (38.15 to 210.3) | 0 (0 to 0) |
| Ethiopia | 0.9 (0.6 to 1.3) | 2.9 (1.8 to 4.3) | 141.33 (67.51 to 247.69) | 0.01 (0 to 0.01) |
| Fiji | 0.4 (0.2 to 0.6) | 0.9 (0.5 to 1.4) | 53.92 (-3.89 to 146.48) | 0 (-0.01 to 0) |
| Finland | 0.4 (0.3 to 0.6) | 0.5 (0.4 to 0.8) | 23 (11.03 to 36.26) | 0.46 (0.38 to 0.54) |
| France | 0.6 (0.4 to 0.8) | 1 (0.6 to 1.5) | 58.34 (39.52 to 79.71) | -0.39 (-0.6 to -0.17) |
| Gabon | 1.1 (0.6 to 1.8) | 2 (0.9 to 3.2) | 91.94 (76.17 to 109.11) | -0.01 (-0.01 to -0.01) |
| Gambia | 1.1 (0.7 to 1.6) | 2.5 (1.4 to 4) | 116.08 (91.17 to 144.24) | -0.02 (-0.02 to -0.01) |
| Georgia | 0.6 (0.4 to 0.9) | 2.7 (1.2 to 4.2) | 123.81 (13.07 to 343) | -0.14 (-0.18 to -0.1) |
| Germany | 0.6 (0.4 to 0.9) | 1 (0.6 to 1.4) | 28.18 (5.19 to 56.19) | 0.48 (0.32 to 0.64) |
| Ghana | 1 (0.6 to 1.4) | 2.1 (1.2 to 3.2) | 97.6 (74.32 to 123.99) | -0.02 (-0.02 to -0.02) |
| Greece | 0.5 (0.3 to 0.8) | 1 (0.6 to 1.4) | 37.68 (0.83 to 87.99) | 1.59 (1.24 to 1.94) |
| Greenland | 1 (0.6 to 1.5) | 1.1 (0.7 to 1.6) | 3.26 (0.46 to 6.14) | 0.04 (0.04 to 0.04) |
| Grenada | 0.6 (0.4 to 0.9) | 0.9 (0.6 to 1.3) | 19.48 (-1.24 to 44.54) | 0.13 (-0.03 to 0.29) |
| Guam | 0.6 (0.4 to 0.9) | 1.4 (0.8 to 2.3) | 96.5 (63.32 to 136.42) | -0.01 (-0.01 to -0.01) |
| Guatemala | 1.6 (1 to 2.3) | 2.8 (1.7 to 4.3) | 57.17 (36.93 to 80.39) | 0.08 (-0.04 to 0.2) |
| Guinea | 1.2 (0.8 to 1.9) | 2.5 (1.5 to 3.9) | 114.44 (102.3 to 127.32) | 0 (0 to 0) |
| Guinea-Bissau | 1.4 (0.8 to 2.3) | 1.8 (1 to 2.9) | 81.29 (18.94 to 176.33) | -0.01 (-0.01 to -0.01) |
| Guyana | 0.8 (0.5 to 1.1) | 1.6 (0.8 to 2.6) | 59.38 (16.64 to 117.79) | 0 (-0.01 to 0.01) |
| Haiti | 0.9 (0.5 to 1.3) | 1.2 (0.6 to 1.9) | 37.48 (36.12 to 38.85) | 0.02 (0 to 0.04) |
| Honduras | 1.5 (1 to 2.3) | 3.2 (2 to 4.8) | 75.71 (41.76 to 117.8) | 0.18 (0.03 to 0.33) |
| Hungary | 0.5 (0.3 to 0.8) | 2 (1.2 to 3.1) | 118.3 (16.44 to 309.24) | 0 (0 to 0.01) |
| Iceland | 0.6 (0.4 to 0.9) | 0.7 (0.4 to 1) | 12.32 (9.7 to 15) | 0.01 (0.01 to 0.01) |
| India | 1.1 (0.7 to 1.6) | 2.5 (1.6 to 3.7) | 85.26 (42.45 to 140.95) | 0.01 (0 to 0.01) |
| Indonesia | 0.7 (0.5 to 1.1) | 1.9 (1.2 to 2.7) | 79.94 (16.45 to 178.05) | 0 (0 to 0) |
| Iran (Islamic Republic of) | 0.8 (0.5 to 1.2) | 2 (1.2 to 2.8) | 106.62 (74.81 to 144.23) | 0.19 (0.08 to 0.3) |
| Iraq | 1.3 (0.8 to 2) | 3.3 (1.8 to 5) | 163.88 (145.54 to 183.61) | 0 (0 to 0) |
| Ireland | 0.6 (0.4 to 0.9) | 1 (0.6 to 1.5) | 39.08 (21.48 to 59.23) | 0 (0 to 0.01) |
| Israel | 0.6 (0.4 to 0.9) | 1.1 (0.7 to 1.7) | 48.42 (18.31 to 86.21) | 0.01 (0.01 to 0.01) |
| Italy | 0.8 (0.5 to 1.2) | 1.3 (0.8 to 1.8) | 34.33 (16.59 to 54.78) | 0.24 (0.22 to 0.27) |
| Jamaica | 0.7 (0.4 to 1) | 1.1 (0.6 to 1.6) | 29.55 (3.31 to 62.45) | 0.4 (-0.1 to 0.91) |
| Japan | 1.8 (1.2 to 2.7) | 1.9 (1.3 to 2.8) | 3.59 (0.21 to 7.07) | 0.21 (0.14 to 0.28) |
| Jordan | 0.6 (0.4 to 0.9) | 2.4 (1.3 to 3.7) | 123.26 (16.43 to 328.12) | 0 (0 to 0) |
| Kazakhstan | 0.8 (0.5 to 1.2) | 1.8 (0.9 to 3.1) | 80.2 (42.59 to 127.74) | 0.01 (0.01 to 0.01) |
| Kenya | 1 (0.6 to 1.4) | 2.9 (1.9 to 4.3) | 136.92 (79.31 to 213.04) | 0.01 (-0.01 to 0.02) |
| Kiribati | 0.4 (0.2 to 0.6) | 0.4 (0.2 to 0.6) | 1.46 (-0.19 to 3.14) | 0 (0 to 0) |
| Kuwait | 0.8 (0.5 to 1.2) | 1.6 (0.9 to 2.7) | 77.38 (53.59 to 104.85) | 0.03 (0.02 to 0.04) |
| Kyrgyzstan | 1.2 (0.7 to 1.8) | 2.4 (1.4 to 3.9) | 112.05 (101.11 to 123.59) | 0.01 (0.01 to 0.02) |
| Lao People's Democratic Republic | 0.5 (0.3 to 0.8) | 0.7 (0.4 to 1) | 12.55 (-1.28 to 28.32) | 0.01 (0.01 to 0.01) |
| Latvia | 0.5 (0.3 to 0.8) | 1.8 (0.9 to 3.5) | 89.59 (2.48 to 250.75) | 0.01 (0.01 to 0.01) |
| Lebanon | 0.7 (0.4 to 1.3) | 2.8 (1.6 to 4.6) | 142.42 (40.32 to 318.82) | 0 (0 to 0) |
| Lesotho | 0.7 (0.4 to 1) | 2.5 (1.3 to 4.3) | 114.31 (20.23 to 281.99) | 0.02 (0.02 to 0.03) |
| Liberia | 1.2 (0.7 to 1.8) | 1.8 (1.1 to 2.8) | 83.8 (53.01 to 120.77) | 0.01 (0 to 0.01) |
| Libya | 0.7 (0.4 to 1.1) | 2.8 (1.5 to 4.2) | 153.95 (59.07 to 305.41) | 0.74 (0.6 to 0.88) |
| Lithuania | 0.6 (0.4 to 0.8) | 2.3 (1.2 to 3.8) | 113.77 (7.17 to 326.38) | 0 (0 to 0) |
| Luxembourg | 0.7 (0.4 to 1) | 1.2 (0.8 to 1.7) | 49.94 (21.58 to 84.92) | 0.02 (0.02 to 0.02) |
| Madagascar | 1 (0.6 to 1.5) | 2.6 (1.6 to 4) | 140.63 (123.17 to 159.46) | -0.02 (-0.02 to -0.01) |
| Malawi | 0.6 (0.4 to 0.9) | 2.7 (1.6 to 4) | 145.58 (28.07 to 370.91) | -0.02 (-0.02 to -0.02) |
| Malaysia | 0.5 (0.3 to 0.8) | 1 (0.6 to 1.5) | 40.69 (-1.88 to 101.73) | 0.01 (0.01 to 0.01) |
| Maldives | 0.7 (0.4 to 1) | 1.3 (0.8 to 2.3) | 58.95 (18.91 to 112.46) | 0.01 (0.01 to 0.02) |
| Mali | 1.2 (0.7 to 1.8) | 2.5 (1.4 to 3.8) | 114.72 (111.14 to 118.35) | 0.01 (0.01 to 0.01) |
| Malta | 0.6 (0.4 to 0.9) | 1 (0.7 to 1.4) | 37.97 (5.31 to 80.77) | 0.02 (0.02 to 0.03) |
| Marshall Islands | 0.4 (0.2 to 0.6) | 0.4 (0.2 to 0.6) | 4.07 (0.14 to 8.16) | 0.01 (0.01 to 0.02) |
| Mauritania | 1 (0.6 to 1.5) | 2.2 (1.2 to 3.8) | 101.81 (71.15 to 137.96) | 0.01 (0.01 to 0.01) |
| Mauritius | 0.5 (0.3 to 0.8) | 0.6 (0.4 to 0.9) | 10.66 (1.42 to 20.75) | 0 (0 to 0) |
| Mexico | 1.9 (1.2 to 2.7) | 3.1 (2 to 4.5) | 48.86 (31.46 to 68.56) | 0.06 (0.05 to 0.08) |
| Micronesia (Federated States of) | 0.4 (0.2 to 0.6) | 0.4 (0.2 to 0.6) | 0.03 (0 to 0.07) | 0 (-0.01 to 0.01) |
| Monaco | 0.6 (0.4 to 0.9) | 1 (0.7 to 1.5) | 40.04 (4.98 to 86.82) | 0.01 (0.01 to 0.01) |
| Mongolia | 0.5 (0.4 to 0.8) | 1.4 (0.9 to 2.1) | 60.72 (-5.03 to 172) | 0 (0 to 0) |
| Montenegro | 0.6 (0.4 to 0.9) | 3.1 (2 to 4.6) | 188.69 (49.8 to 456.35) | -0.23 (-0.36 to -0.1) |
| Morocco | 0.8 (0.5 to 1.2) | 2.3 (1.1 to 3.9) | 121.79 (63.73 to 200.43) | 0 (0 to 0) |
| Mozambique | 0.6 (0.4 to 0.9) | 3 (1.8 to 4.6) | 159.91 (21.56 to 455.72) | -0.01 (-0.02 to -0.01) |
| Myanmar | 0.6 (0.4 to 0.9) | 1.2 (0.7 to 1.8) | 48.31 (5.17 to 109.14) | 0 (0 to 0) |
| Namibia | 0.6 (0.4 to 1) | 2.1 (1.2 to 3.4) | 99.03 (12.17 to 253.14) | -0.01 (-0.01 to -0.01) |
| Nauru | 0.4 (0.3 to 0.6) | 0.6 (0.4 to 1) | 31.57 (10.97 to 55.99) | -0.03 (-0.03 to -0.03) |
| Nepal | 0.8 (0.5 to 1.2) | 2.5 (1.5 to 4) | 108.97 (29.57 to 237.05) | -0.54 (-0.68 to -0.39) |
| Netherlands | 0.7 (0.4 to 1) | 1.2 (0.7 to 1.8) | 50.65 (27.96 to 77.36) | 0.81 (0.57 to 1.05) |
| New Zealand | 0.6 (0.4 to 0.9) | 0.6 (0.4 to 0.9) | 0.24 (0.23 to 0.26) | -0.19 (-0.26 to -0.12) |
| Nicaragua | 1.5 (1 to 2.1) | 2.5 (1.6 to 3.7) | 48 (24.62 to 75.76) | 0.07 (-0.01 to 0.14) |
| Niger | 1 (0.6 to 1.5) | 1.9 (1.1 to 3) | 85.5 (77.94 to 93.39) | -0.01 (-0.01 to -0.01) |
| Nigeria | 1.3 (0.9 to 2) | 2.5 (1.6 to 3.7) | 104.17 (86.49 to 123.52) | -0.02 (-0.03 to -0.01) |
| Niue | 0.4 (0.2 to 0.6) | 0.4 (0.2 to 0.6) | -0.01 (-0.01 to -0.01) | 0.01 (0 to 0.01) |
| North Macedonia | 0.8 (0.5 to 1.2) | 3.4 (2 to 5.1) | 182.69 (86.85 to 327.68) | 0 (0 to 0) |
| Northern Mariana Islands | 0.4 (0.3 to 0.7) | 0.6 (0.4 to 0.8) | 22.38 (17.34 to 27.64) | -0.01 (-0.01 to 0) |
| Norway | 1.1 (0.7 to 1.5) | 1.2 (0.8 to 1.8) | 10.33 (4.3 to 16.72) | 0.26 (0.13 to 0.4) |
| Oman | 0.8 (0.5 to 1.1) | 1.5 (0.9 to 2.4) | 80.41 (59.13 to 104.55) | 0 (0 to 0) |
| Pakistan | 1.3 (0.8 to 1.9) | 2.4 (1.5 to 3.7) | 82.22 (74.98 to 89.76) | 0.01 (0.01 to 0.01) |
| Palau | 0.4 (0.2 to 0.6) | 0.4 (0.2 to 0.6) | 0.2 (0 to 0.41) | 0.01 (0.01 to 0.02) |
| Palestine | 0.7 (0.5 to 1.1) | 3 (1.6 to 4.8) | 153.17 (40.94 to 354.77) | 0 (0 to 0) |
| Panama | 1.5 (0.9 to 2.2) | 2.3 (1.5 to 3.3) | 40 (26.82 to 54.54) | 0.16 (-0.01 to 0.34) |
| Papua New Guinea | 0.4 (0.2 to 0.6) | 1 (0.6 to 1.7) | 66.79 (-2.9 to 186.49) | 0 (0 to 0) |
| Paraguay | 0.5 (0.3 to 0.8) | 2.1 (1.2 to 3.3) | 134.86 (24.47 to 343.17) | 0.02 (-0.01 to 0.05) |
| Peru | 1.4 (0.9 to 2.1) | 2.5 (1.5 to 3.8) | 75.7 (75.54 to 75.85) | 0.06 (0.01 to 0.1) |
| Philippines | 0.8 (0.5 to 1.2) | 1.7 (1.1 to 2.5) | 58.79 (20.96 to 108.47) | 0.48 (0.35 to 0.61) |
| Poland | 0.6 (0.4 to 0.8) | 1.9 (1.2 to 2.9) | 112.45 (21.24 to 272.28) | -0.35 (-0.42 to -0.27) |
| Portugal | 0.6 (0.4 to 0.9) | 1.2 (0.8 to 1.7) | 52.3 (12.39 to 106.39) | 0 (0 to 0.01) |
| Puerto Rico | 0.5 (0.3 to 0.8) | 0.8 (0.5 to 1.2) | 32.2 (8.55 to 60.99) | -0.74 (-1.21 to -0.27) |
| Qatar | 1.2 (0.7 to 1.9) | 2 (1.1 to 3) | 102.7 (54.37 to 166.16) | 0.01 (0.01 to 0.01) |
| Republic of Korea | 1.7 (1.1 to 2.6) | 1.8 (1.1 to 2.7) | 1.06 (0.18 to 1.94) | 0.01 (0 to 0.01) |
| Republic of Moldova | 0.9 (0.5 to 1.3) | 2.6 (1.4 to 4.3) | 128.37 (75.14 to 197.76) | 0 (0 to 0.01) |
| Romania | 0.6 (0.4 to 0.9) | 2.1 (1.1 to 3.2) | 121.03 (43.69 to 239.98) | 0 (0 to 0) |
| Russian Federation | 0.9 (0.6 to 1.3) | 2.6 (1.6 to 3.8) | 118.77 (60.47 to 198.25) | 0.01 (0.01 to 0.01) |
| Rwanda | 0.5 (0.3 to 0.7) | 1.4 (0.9 to 2.3) | 79.9 (1.34 to 219.38) | -0.01 (-0.02 to 0) |
| Saint Kitts and Nevis | 0.6 (0.4 to 1) | 0.8 (0.5 to 1.1) | 11.25 (1.1 to 22.43) | 0.35 (-0.06 to 0.76) |
| Saint Lucia | 0.6 (0.4 to 0.9) | 1.1 (0.6 to 1.7) | 33.64 (-2.25 to 82.72) | 0.14 (-0.02 to 0.29) |
| Saint Vincent and the Grenadines | 0.6 (0.4 to 1) | 0.9 (0.6 to 1.4) | 20.78 (0.13 to 45.68) | 0.16 (-0.02 to 0.34) |
| Samoa | 0.4 (0.2 to 0.6) | 0.4 (0.2 to 0.6) | 0.87 (0.01 to 1.74) | 0 (0 to 0) |
| San Marino | 0.9 (0.6 to 1.3) | 1.7 (1.1 to 2.4) | 79.58 (70.85 to 88.74) | -0.01 (-0.01 to -0.01) |
| Sao Tome and Principe | 1.2 (0.7 to 1.9) | 1.5 (0.9 to 2.3) | 67.53 (25.21 to 124.16) | 0.01 (0.01 to 0.01) |
| Saudi Arabia | 0.8 (0.5 to 1.2) | 1 (0.5 to 1.5) | 41.9 (19.36 to 68.7) | 0 (0 to 0.01) |
| Senegal | 0.9 (0.6 to 1.4) | 2.8 (1.7 to 4.2) | 125.4 (64 to 209.78) | 0 (0 to 0) |
| Serbia | 0.4 (0.3 to 0.6) | 1.9 (1.2 to 3) | 171.86 (43.63 to 414.56) | -0.49 (-0.77 to -0.21) |
| Seychelles | 0.5 (0.3 to 0.8) | 1.7 (1.1 to 2.5) | 78.18 (-4.26 to 231.61) | 0.01 (0.01 to 0.01) |
| Sierra Leone | 1.2 (0.7 to 1.9) | 1.3 (0.8 to 2) | 55.21 (4.89 to 129.68) | 0 (0 to 0.01) |
| Singapore | 1.8 (1.1 to 2.7) | 1.8 (1.2 to 2.7) | 2.16 (1.23 to 3.09) | 0.02 (0.01 to 0.03) |
| Slovakia | 0.5 (0.3 to 0.7) | 1.8 (1.1 to 2.8) | 108.68 (4.86 to 315.31) | 0.01 (0 to 0.01) |
| Slovenia | 0.5 (0.3 to 0.8) | 1.6 (0.9 to 2.9) | 94.97 (11.9 to 239.72) | 0.03 (0.02 to 0.03) |
| Solomon Islands | 0.4 (0.2 to 0.6) | 0.4 (0.2 to 0.6) | 1.59 (0.29 to 2.91) | -0.01 (-0.01 to -0.01) |
| Somalia | 0.8 (0.5 to 1.4) | 2 (1.2 to 3.1) | 113.36 (82.8 to 149.04) | -0.02 (-0.02 to -0.02) |
| South Africa | 0.9 (0.6 to 1.3) | 2.3 (1.4 to 3.3) | 93.8 (47.19 to 155.16) | 0.01 (0 to 0.01) |
| South Sudan | 0.9 (0.5 to 1.6) | 1.7 (0.9 to 2.9) | 91.75 (78.54 to 105.94) | -0.02 (-0.03 to -0.02) |
| Spain | 0.7 (0.5 to 1) | 1.3 (0.9 to 1.9) | 58.77 (36.74 to 84.34) | 2.11 (1.74 to 2.48) |
| Sri Lanka | 0.5 (0.3 to 0.8) | 0.8 (0.5 to 1.2) | 23.31 (-1.15 to 53.83) | -0.01 (-0.01 to -0.01) |
| Sudan | 1.1 (0.6 to 1.8) | 1.7 (0.8 to 2.8) | 89.26 (48.01 to 141.99) | 0 (0 to 0) |
| Suriname | 0.7 (0.5 to 1.1) | 1.6 (0.9 to 2.5) | 59.13 (15.08 to 120.04) | 0.15 (-0.03 to 0.33) |
| Sweden | 1 (0.6 to 1.4) | 1.4 (0.9 to 2) | 33.31 (21.45 to 46.32) | 2.2 (1.88 to 2.53) |
| Switzerland | 0.6 (0.4 to 0.9) | 1.1 (0.7 to 1.5) | 41.74 (14.35 to 75.69) | 0.02 (0.01 to 0.02) |
| Syrian Arab Republic | 0.5 (0.3 to 0.8) | 0.8 (0.4 to 1.3) | 26.37 (3.18 to 54.78) | -0.01 (-0.01 to 0) |
| Taiwan (Province of China) | 0.3 (0.2 to 0.4) | 0.3 (0.2 to 0.4) | 2.36 (0.88 to 3.86) | 0.68 (0.58 to 0.79) |
| Tajikistan | 0.9 (0.6 to 1.3) | 2.2 (1.3 to 3.4) | 99.9 (53.63 to 160.12) | 0.03 (0.03 to 0.03) |
| Thailand | 0.5 (0.3 to 0.8) | 0.7 (0.4 to 1.1) | 14.6 (-1.22 to 32.95) | 0 (0 to 0) |
| Timor-Leste | 0.5 (0.3 to 0.8) | 1 (0.6 to 1.6) | 37.94 (-3.72 to 97.62) | 0 (0 to 0) |
| Togo | 0.7 (0.5 to 1.1) | 1.9 (1.1 to 2.9) | 85.57 (29.39 to 166.16) | -0.02 (-0.02 to -0.02) |
| Tokelau | 0.4 (0.2 to 0.6) | 0.4 (0.2 to 0.6) | -0.01 (-0.01 to 0) | 0.01 (0 to 0.01) |
| Tonga | 0.4 (0.2 to 0.6) | 0.4 (0.2 to 0.6) | 0.14 (-0.02 to 0.3) | 0 (0 to 0) |
| Trinidad and Tobago | 0.7 (0.4 to 1) | 1 (0.7 to 1.5) | 28.45 (3.17 to 59.92) | 0.03 (0 to 0.06) |
| Tunisia | 0.6 (0.4 to 0.9) | 2.8 (1.5 to 4.5) | 141.13 (18.82 to 389.34) | 0 (0 to 0) |
| Turkey | 0.9 (0.5 to 1.4) | 1.7 (1 to 2.8) | 76.21 (55.02 to 100.31) | 0.06 (0.05 to 0.08) |
| Turkmenistan | 0.9 (0.5 to 1.3) | 2.2 (1.3 to 3.6) | 101.69 (54.34 to 163.57) | 0.02 (0.01 to 0.02) |
| Tuvalu | 0.4 (0.3 to 0.6) | 0.7 (0.4 to 1) | 32.2 (11.32 to 57) | 0.02 (0.02 to 0.02) |
| Uganda | 0.5 (0.3 to 0.8) | 1.7 (1.1 to 2.6) | 97.54 (11.95 to 248.56) | -0.02 (-0.02 to -0.01) |
| Ukraine | 0.7 (0.5 to 1.1) | 2.2 (1.2 to 3.8) | 100.73 (23.31 to 226.76) | 0.01 (0.01 to 0.01) |
| United Arab Emirates | 0.7 (0.5 to 1.1) | 1.7 (1 to 2.8) | 90.51 (49.12 to 143.39) | 0.02 (0.02 to 0.02) |
| United Kingdom | 0.6 (0.4 to 0.9) | 1.1 (0.7 to 1.6) | 53.7 (37.96 to 71.23) | 2.65 (2.23 to 3.07) |
| United Republic of Tanzania | 0.7 (0.5 to 1) | 2.2 (1.4 to 3.2) | 127.34 (57.44 to 228.29) | 0.78 (0.63 to 0.94) |
| United States of America | 1.4 (0.9 to 2) | 2.1 (1.4 to 3.1) | 30.22 (11.24 to 52.44) | 0.44 (0.32 to 0.56) |
| United States Virgin Islands | 0.7 (0.5 to 1.1) | 1.1 (0.7 to 1.7) | 34.31 (16.34 to 55.06) | 0.14 (-0.04 to 0.31) |
| Uruguay | 1 (0.6 to 1.5) | 1.6 (1 to 2.3) | 27.12 (-2.16 to 65.17) | 0 (0 to 0) |
| Uzbekistan | 1 (0.6 to 1.5) | 1.8 (0.9 to 2.9) | 83.24 (81.83 to 84.66) | 0.02 (0.02 to 0.02) |
| Vanuatu | 0.4 (0.2 to 0.6) | 0.4 (0.3 to 0.6) | 3.17 (-0.42 to 6.87) | -0.01 (-0.01 to -0.01) |
| Venezuela (Bolivarian Republic of) | 1.4 (0.9 to 2.1) | 2.3 (1.5 to 3.5) | 42.7 (21.39 to 67.74) | 0.06 (-0.02 to 0.14) |
| Viet Nam | 0.5 (0.3 to 0.8) | 0.6 (0.4 to 0.9) | 9.26 (-0.99 to 20.57) | 0.01 (0 to 0.01) |
| Yemen | 0.9 (0.5 to 1.4) | 1.3 (0.6 to 2.3) | 63.47 (39.22 to 91.94) | 0 (0 to 0) |
| Zambia | 0.7 (0.5 to 1.1) | 2.7 (1.6 to 4) | 144.76 (58.15 to 278.8) | -0.03 (-0.03 to -0.02) |
| Zimbabwe | 0.7 (0.4 to 1) | 2.6 (1.4 to 4.1) | 121.2 (20.54 to 305.93) | -0.01 (-0.02 to -0.01) |

Notes: Countries and territories were ordered alphabetically. ASR=age-standardized rate. CI=confidence interval. EAPC=estimated annual percentage change. GBD=Global Burden of Diseases, Injuries and Risk Factors Study. SDI=Socio-demographic index. UI=uncertainty interval. YLD=years lived with disability.

| **Supplementary Table S3** Male and female age-standardized years lived with disability rates (ASYRs), and ASYR sex ratio, in 2020 and 2021 | | | | | | | | |
| --- | --- | --- | --- | --- | --- | --- | --- | --- |
|  |  | **2020** | | |  | **2021** | | |
| **GBD super regions** | **GBD regions** | **ASYR in males (per 100,000)** | **ASYR in females (per 100,000)** | **ASYR male to female ratio** |  | **ASYR in males (per 100,000)** | **ASYR in females (per 100,000)** | **ASYR male to female ratio** |
| Global |  | 0.25 (0.16 to 0.39) | 0.25 (0.16 to 0.38) | 1.00 |  | 1.15 (0.72 to 1.73) | 1.14 (0.71 to 1.71) | 1.01 |
| Low SDI |  | 0.44 (0.27 to 0.66) | 0.42 (0.26 to 0.63) | 1.05 |  | 1.83 (1.12 to 2.77) | 1.77 (1.07 to 2.66) | 1.03 |
| Low-middle SDI |  | 0.4 (0.25 to 0.6) | 0.39 (0.24 to 0.58) | 1.03 |  | 1.7 (1.02 to 2.49) | 1.64 (0.98 to 2.41) | 1.04 |
| Middle SDI |  | 0.21 (0.13 to 0.31) | 0.2 (0.13 to 0.31) | 1.05 |  | 0.97 (0.6 to 1.43) | 0.95 (0.58 to 1.41) | 1.02 |
| High-middle SDI |  | 0.12 (0.07 to 0.19) | 0.13 (0.07 to 0.19) | 0.92 |  | 0.71 (0.44 to 1.08) | 0.73 (0.46 to 1.11) | 0.97 |
| High SDI |  | 0.12 (0.07 to 0.17) | 0.11 (0.07 to 0.16) | 1.09 |  | 0.58 (0.36 to 0.85) | 0.56 (0.35 to 0.83) | 1.04 |
| Central Europe, eastern Europe, and central Asia |  | 0.29 (0.18 to 0.43) | 0.28 (0.18 to 0.42) | 1.04 |  | 1.82 (1.13 to 2.72) | 1.75 (1.1 to 2.65) | 1.04 |
|  | Central Asia | 0.36 (0.2 to 0.57) | 0.35 (0.19 to 0.56) | 1.03 |  | 1.55 (0.91 to 2.36) | 1.49 (0.86 to 2.29) | 1.04 |
|  | Central Europe | 0.16 (0.1 to 0.25) | 0.16 (0.09 to 0.24) | 1.00 |  | 1.77 (1.07 to 2.62) | 1.7 (1.03 to 2.51) | 1.04 |
|  | Eastern Europe | 0.32 (0.2 to 0.48) | 0.31 (0.19 to 0.46) | 1.03 |  | 1.99 (1.23 to 3.05) | 1.91 (1.19 to 2.95) | 1.04 |
| Hign-income |  | 0.12 (0.07 to 0.17) | 0.11 (0.07 to 0.17) | 1.09 |  | 0.57 (0.36 to 0.84) | 0.56 (0.35 to 0.82) | 1.02 |
|  | Australasia | 0 (0 to 0.01) | 0 (0 to 0.01) | NA |  | 0.01 (0.01 to 0.02) | 0.01 (0.01 to 0.02) | 1.00 |
|  | High-income Asia Pacific | 0.01 (0.01 to 0.02) | 0.01 (0.01 to 0.02) | 1.00 |  | 0.1 (0.06 to 0.15) | 0.1 (0.06 to 0.15) | 1.00 |
|  | High-income North America | 0.15 (0.09 to 0.23) | 0.15 (0.09 to 0.22) | 1.00 |  | 0.83 (0.52 to 1.23) | 0.8 (0.49 to 1.18) | 1.04 |
|  | Southern Latin America | 0.12 (0.07 to 0.18) | 0.11 (0.07 to 0.18) | 1.09 |  | 0.66 (0.38 to 1.04) | 0.64 (0.37 to 1.01) | 1.03 |
|  | Western Europe | 0.13 (0.08 to 0.2) | 0.13 (0.08 to 0.19) | 1.00 |  | 0.56 (0.35 to 0.84) | 0.54 (0.34 to 0.81) | 1.04 |
| Latin America and Caribbean |  | 0.41 (0.25 to 0.6) | 0.39 (0.24 to 0.59) | 1.05 |  | 1.55 (0.94 to 2.31) | 1.5 (0.91 to 2.25) | 1.03 |
|  | Andean Latin America | 0.66 (0.39 to 1.02) | 0.64 (0.38 to 1) | 1.03 |  | 1.91 (1.14 to 2.95) | 1.86 (1.13 to 2.87) | 1.03 |
|  | Caribbean | 0.15 (0.07 to 0.25) | 0.15 (0.07 to 0.24) | 1.00 |  | 0.54 (0.3 to 0.85) | 0.53 (0.29 to 0.83) | 1.02 |
|  | Tropical Latin America | 0.41 (0.26 to 0.61) | 0.4 (0.25 to 0.59) | 1.02 |  | 1.64 (1.01 to 2.45) | 1.57 (0.98 to 2.38) | 1.04 |
|  | Central Latin America | 0.39 (0.23 to 0.57) | 0.37 (0.23 to 0.56) | 1.05 |  | 1.56 (0.95 to 2.32) | 1.51 (0.92 to 2.26) | 1.03 |
| North Africa and Middle East | North Africa and Middle East | 0.44 (0.25 to 0.68) | 0.42 (0.24 to 0.67) | 1.05 |  | 1.44 (0.87 to 2.19) | 1.41 (0.84 to 2.12) | 1.02 |
| South Asia | South Asia | 0.38 (0.23 to 0.58) | 0.37 (0.22 to 0.57) | 1.03 |  | 1.82 (1.11 to 2.74) | 1.75 (1.07 to 2.63) | 1.04 |
| Southeast Asia, east Asia, and Oceania |  | 0.03 (0.02 to 0.05) | 0.03 (0.02 to 0.05) | 1.00 |  | 0.29 (0.18 to 0.43) | 0.29 (0.18 to 0.43) | 1.00 |
|  | Southeast Asia | 0.09 (0.05 to 0.13) | 0.08 (0.05 to 0.12) | 1.12 |  | 0.84 (0.52 to 1.24) | 0.8 (0.5 to 1.19) | 1.05 |
|  | East Asia | 0.01 (0 to 0.01) | 0.01 (0 to 0.01) | 1.00 |  | 0 (0 to 0) | 0 (0 to 0) | NA |
|  | Oceania | 0.02 (0.01 to 0.03) | 0.01 (0.01 to 0.03) | 1.00 |  | 0.59 (0.28 to 1.05) | 0.56 (0.27 to 1) | 1.05 |
| Sub-Saharan Africa |  | 0.47 (0.3 to 0.71) | 0.46 (0.29 to 0.69) | 1.02 |  | 1.91 (1.19 to 2.85) | 1.84 (1.15 to 2.75) | 1.04 |
|  | Western Sub-Saharan Africa | 0.61 (0.38 to 0.93) | 0.59 (0.37 to 0.9) | 1.03 |  | 1.77 (1.11 to 2.67) | 1.7 (1.06 to 2.58) | 1.04 |
|  | Eastern Sub-Saharan Africa | 0.31 (0.19 to 0.46) | 0.3 (0.19 to 0.44) | 1.03 |  | 2.03 (1.26 to 3.01) | 1.96 (1.21 to 2.92) | 1.04 |
|  | Central Sub-Saharan Africa | 0.63 (0.36 to 1) | 0.61 (0.34 to 0.95) | 1.03 |  | 2.14 (1.26 to 3.2) | 2.07 (1.22 to 3.09) | 1.03 |
|  | Southern Sub-Saharan Africa | 0.27 (0.17 to 0.4) | 0.26 (0.16 to 0.38) | 1.04 |  | 1.76 (1.1 to 2.58) | 1.71 (1.07 to 2.53) | 1.03 |

NA=not applicable.

| **Supplementary Table S4** Cause-specific age-standardized years lived with disability of Guillain-Barré syndrome at global and supper-regional levels, before and during the years of COVID-19 pandemic | | | | |
| --- | --- | --- | --- | --- |
| **Locations** | **Causes/triggers** | **2019** | **2020** | **2021** |
| Global | COVID-19 | 0 (0 to 0) | 0.25 (0.16 to 0.38) | 1.14 (0.71 to 1.72) |
|  | Diarrheal diseases | 0.07 (0.04 to 0.11) | 0.07 (0.04 to 0.11) | 0.07 (0.04 to 0.11) |
|  | Lower respiratory infections | 0.07 (0.04 to 0.12) | 0.07 (0.04 to 0.12) | 0.07 (0.04 to 0.12) |
|  | Other neurological disorders | 0.23 (0.13 to 0.35) | 0.23 (0.13 to 0.36) | 0.23 (0.13 to 0.36) |
|  | Other unspecified infectious diseases | 0.04 (0.02 to 0.07) | 0.04 (0.02 to 0.07) | 0.04 (0.02 to 0.07) |
|  | Upper respiratory infections | 0.19 (0.12 to 0.3) | 0.19 (0.12 to 0.3) | 0.19 (0.12 to 0.3) |
|  | Zika virus | 0 (0 to 0) | 0 (0 to 0) | 0 (0 to 0) |
| Central Europe, Eastern Europe, and Central Asia | COVID-19 | 0 (0 to 0) | 0.28 (0.18 to 0.42) | 1.78 (1.11 to 2.68) |
|  | Diarrheal diseases | 0.06 (0.03 to 0.09) | 0.06 (0.03 to 0.09) | 0.06 (0.03 to 0.09) |
|  | Lower respiratory infections | 0.06 (0.03 to 0.1) | 0.06 (0.03 to 0.1) | 0.06 (0.03 to 0.1) |
|  | Other neurological disorders | 0.19 (0.11 to 0.3) | 0.19 (0.11 to 0.3) | 0.19 (0.11 to 0.3) |
|  | Other unspecified infectious diseases | 0.04 (0.02 to 0.06) | 0.04 (0.02 to 0.06) | 0.04 (0.02 to 0.06) |
|  | Upper respiratory infections | 0.16 (0.1 to 0.26) | 0.16 (0.1 to 0.26) | 0.16 (0.1 to 0.26) |
|  | Zika virus | 0 (0 to 0) | 0 (0 to 0) | 0 (0 to 0) |
| High-income | COVID-19 | 0 (0 to 0) | 0.11 (0.07 to 0.17) | 0.57 (0.35 to 0.83) |
|  | Diarrheal diseases | 0.11 (0.06 to 0.18) | 0.11 (0.06 to 0.18) | 0.11 (0.07 to 0.18) |
|  | Lower respiratory infections | 0.11 (0.06 to 0.19) | 0.12 (0.06 to 0.19) | 0.12 (0.06 to 0.19) |
|  | Other neurological disorders | 0.38 (0.21 to 0.58) | 0.38 (0.21 to 0.58) | 0.38 (0.21 to 0.59) |
|  | Other unspecified infectious diseases | 0.07 (0.04 to 0.11) | 0.07 (0.04 to 0.11) | 0.07 (0.04 to 0.11) |
|  | Upper respiratory infections | 0.32 (0.2 to 0.48) | 0.32 (0.2 to 0.49) | 0.32 (0.2 to 0.49) |
|  | Zika virus | 0 (0 to 0) | 0 (0 to 0) | 0 (0 to 0) |
| Latin America  and Caribbean | COVID-19 | 0 (0 to 0) | 0.4 (0.24 to 0.59) | 1.52 (0.92 to 2.28) |
|  | Diarrheal diseases | 0.09 (0.06 to 0.15) | 0.09 (0.05 to 0.15) | 0.09 (0.05 to 0.15) |
|  | Lower respiratory infections | 0.1 (0.05 to 0.16) | 0.1 (0.05 to 0.16) | 0.1 (0.05 to 0.16) |
|  | Other neurological disorders | 0.32 (0.18 to 0.48) | 0.32 (0.18 to 0.48) | 0.32 (0.18 to 0.48) |
|  | Other unspecified infectious diseases | 0.06 (0.03 to 0.09) | 0.06 (0.03 to 0.09) | 0.06 (0.03 to 0.09) |
|  | Upper respiratory infections | 0.27 (0.17 to 0.41) | 0.27 (0.17 to 0.41) | 0.27 (0.17 to 0.41) |
|  | Zika virus | 0 (0 to 0.01) | 0 (0 to 0) | 0 (0 to 0) |
| North Africa and Middle East | COVID-19 | 0 (0 to 0) | 0.43 (0.25 to 0.67) | 1.43 (0.86 to 2.16) |
|  | Diarrheal diseases | 0.05 (0.03 to 0.09) | 0.05 (0.03 to 0.09) | 0.05 (0.03 to 0.09) |
|  | Lower respiratory infections | 0.06 (0.03 to 0.09) | 0.06 (0.03 to 0.09) | 0.06 (0.03 to 0.09) |
|  | Other neurological disorders | 0.18 (0.1 to 0.29) | 0.19 (0.1 to 0.29) | 0.19 (0.1 to 0.29) |
|  | Other unspecified infectious diseases | 0.03 (0.02 to 0.06) | 0.03 (0.02 to 0.06) | 0.03 (0.02 to 0.06) |
|  | Upper respiratory infections | 0.16 (0.09 to 0.25) | 0.16 (0.09 to 0.25) | 0.16 (0.09 to 0.25) |
|  | Zika virus | 0 (0 to 0) | 0 (0 to 0) | 0 (0 to 0) |
| South Asia | COVID-19 | 0 (0 to 0) | 0.38 (0.23 to 0.57) | 1.78 (1.09 to 2.68) |
|  | Diarrheal diseases | 0.08 (0.05 to 0.13) | 0.08 (0.05 to 0.13) | 0.08 (0.05 to 0.13) |
|  | Lower respiratory infections | 0.08 (0.04 to 0.14) | 0.08 (0.04 to 0.14) | 0.08 (0.04 to 0.14) |
|  | Other neurological disorders | 0.28 (0.15 to 0.44) | 0.28 (0.15 to 0.44) | 0.28 (0.15 to 0.44) |
|  | Other unspecified infectious diseases | 0.05 (0.03 to 0.08) | 0.05 (0.03 to 0.08) | 0.05 (0.03 to 0.08) |
|  | Upper respiratory infections | 0.23 (0.14 to 0.37) | 0.23 (0.14 to 0.37) | 0.23 (0.14 to 0.37) |
|  | Zika virus | 0 (0 to 0) | 0 (0 to 0) | 0 (0 to 0) |
| Southeast Asia, East Asia, and Oceania | COVID-19 | 0 (0 to 0) | 0.03 (0.02 to 0.05) | 0.29 (0.18 to 0.43) |
|  | Diarrheal diseases | 0.03 (0.02 to 0.06) | 0.03 (0.02 to 0.06) | 0.03 (0.02 to 0.06) |
|  | Lower respiratory infections | 0.04 (0.02 to 0.06) | 0.04 (0.02 to 0.06) | 0.04 (0.02 to 0.06) |
|  | Other neurological disorders | 0.12 (0.06 to 0.18) | 0.12 (0.06 to 0.18) | 0.12 (0.06 to 0.18) |
|  | Other unspecified infectious diseases | 0.02 (0.01 to 0.04) | 0.02 (0.01 to 0.04) | 0.02 (0.01 to 0.04) |
|  | Upper respiratory infections | 0.1 (0.06 to 0.16) | 0.1 (0.06 to 0.16) | 0.1 (0.06 to 0.16) |
|  | Zika virus | 0 (0 to 0) | 0 (0 to 0) | 0 (0 to 0) |
| Sub-Saharan Africa | COVID-19 | 0 (0 to 0) | 0.46 (0.29 to 0.7) | 1.87 (1.16 to 2.8) |
|  | Diarrheal diseases | 0.06 (0.03 to 0.1) | 0.06 (0.03 to 0.1) | 0.06 (0.03 to 0.1) |
|  | Lower respiratory infections | 0.06 (0.03 to 0.1) | 0.06 (0.03 to 0.1) | 0.06 (0.03 to 0.1) |
|  | Other neurological disorders | 0.2 (0.11 to 0.32) | 0.2 (0.11 to 0.32) | 0.2 (0.11 to 0.32) |
|  | Other unspecified infectious diseases | 0.04 (0.02 to 0.06) | 0.04 (0.02 to 0.06) | 0.04 (0.02 to 0.06) |
|  | Upper respiratory infections | 0.17 (0.1 to 0.28) | 0.17 (0.1 to 0.28) | 0.17 (0.1 to 0.28) |
|  | Zika virus | 0 (0 to 0) | 0 (0 to 0) | 0 (0 to 0) |

| **Supplementary Table S5** Association between potential influence factors and age-standardized years lived with disability rate in 2021, using generalized linear model with Gaussian distribution and log-link function. | | | | | | | | | | |
| --- | --- | --- | --- | --- | --- | --- | --- | --- | --- | --- |
| **Factors** |  | **All-cause age-standardized YLD rate** | | | |  | **Age-standardized YLD rate due to COVID-19** | | | |
|  |  | ***β*** | **SE** | **t-value** | ***p*** |  | ***β*** | **SE** | **t-value** | ***p*** |
| Number of total vaccinations (per hundred)* | | -0.0033 | 0.0006 | -5.81 | <.001 |  | -0.004 | 0.0009 | -4.56 | <.001 |
| SDI, 2021 | | 0.1196 | 0.2061 | 0.58 | 0.563 |  | 0.3026 | 0.2938 | 1.03 | 0.305 |
| Median of GSI, 2021 | | 0.0036 | 0.0022 | 1.62 | 0.107 |  | 0.0049 | 0.0033 | 1.51 | 0.134 |
| ASYR, 2020* | | 0.5797 | 0.0669 | 8.66 | <0.001 |  | 1.1146 | 0.1606 | 6.94 | <0.001 |
| Number of people vaccinated (per hundred)* | | -0.0066 | 0.0014 | -4.73 | <0.001 |  | -0.0070 | 0.0022 | -3.13 | 0.002 |
| SDI, 2021 | | 0.0211 | 0.2118 | 0.099 | 0.921 |  | 0.1553 | 0.3094 | 0.502 | 0.617 |
| Median of GSI, 2021 | | 0.0021 | 0.0024 | 0.909 | 0.365 |  | 0.0029 | 0.0035 | 0.829 | 0.409 |
| ASYR, 2020 | | 0.5847 | 0.0692 | 8.44 | <0.001 |  | 1.1779 | 0.1784 | 6.604 | <0.001 |
| Number people fully vaccinated (per hundred)* | | -0.0081 | 0.0015 | -5.60 | <0.001 |  | -0.0090 | 0.0023 | -3.91 | <0.001 |
| SDI, 2021 | | 0.1765 | 0.2153 | 0.82 | 0.414 |  | 0.3263 | 0.3043 | 1.07 | 0.285 |
| Median of GSI, 2021 | | 0.0033 | 0.0023 | 1.46 | 0.148 |  | 0.0043 | 0.0034 | 1.27 | 0.205 |
| ASYR, 2020 | | 0.5800 | 0.0691 | 8.39 | <0.001 |  | 1.136 | 0.174 | 6.52 | <0.001 |

ASYR=age-standardized years lived with disability rate; β=regression coefficient; COVID-19=coronavirus disease 2019; GSI=government stringency index; SDI= socio-demographic index; SE=standard error; YLD=years lived with disability.

* Data on December 31, 2021 were obtained. Missing values were filled by data within a 11-day window (5 days before and after December 31, 2021).

| **Supplementary Table S6** COVID-19 vaccine coverage rates of 204 countries and territories, data collected between Dec. 24, 2021 and Jan. 07, 2022 | | | |
| --- | --- | --- | --- |
| **Countries and territories** | **Number of total vaccinations (per 100)** | **Number of people vaccinated (per 100)** | **Number of people fully vaccinated (per 100)** |
| Afghanistan | 11.37 | 10.07 | 9.13 |
| Albania | 83.3 | 40.83 | 37.06 |
| Algeria | NA | NA | NA |
| American Samoa | 146.85 | 71.5 | 64.37 |
| Andorra | 32.64 | 21.59 | 11.04 |
| Angola | NA | NA | NA |
| Antigua and Barbuda | 129.19 | 66.42 | 62.77 |
| Argentina | 175.8 | 85.89 | 68.42 |
| Armenia | 59.71 | 33.98 | 25.7 |
| Australia | 165.36 | 78.27 | 75.76 |
| Austria | 186.56 | 75.03 | 70.64 |
| Azerbaijan | 109.81 | 49.93 | 45.18 |
| Bahamas | 75.18 | 38.57 | 36.44 |
| Bahrain | 221.07 | 81.87 | 80.22 |
| Bangladesh | NA | NA | NA |
| Barbados | 106.51 | 55.42 | 51.08 |
| Belarus | 90.74 | 51.45 | 37.45 |
| Belgium | 189.25 | 76.76 | 75.83 |
| Belize | NA | NA | NA |
| Benin | 14.21 | 13.6 | 10.65 |
| Bermuda | 177.93 | 72.27 | 70.84 |
| Bhutan | 148.45 | 75.73 | 72.72 |
| Bolivia (Plurinational State of) | 83.88 | 48.58 | 38.54 |
| Bosnia and Herzegovina | NA | NA | NA |
| Botswana | NA | 44.45 | 39.31 |
| Brazil | 154.95 | 77.32 | 66.82 |
| Brunei Darussalam | 200.09 | 90.4 | 89.24 |
| Bulgaria | 55.57 | NA | 28.42 |
| Burkina Faso | 5.26 | 4.89 | 3.2 |
| Burundi | NA | NA | NA |
| Cabo Verde | NA | NA | NA |
| Cambodia | 182.91 | 85.14 | 81.56 |
| Cameroon | 3.65 | 2.99 | 2.36 |
| Canada | 182.12 | 82.74 | 76.69 |
| Central African Republic | NA | NA | NA |
| Chad | NA | NA | NA |
| Chile | 228.59 | 88.59 | 84.65 |
| China | 201.42 | 88.36 | 84.68 |
| Colombia | 126.39 | 74.34 | 55.41 |
| Comoros | 69.5 | 40.08 | 29.41 |
| Congo | 12.71 | 10.43 | NA |
| Cook Islands | 150 | 75.96 | 74.04 |
| Costa Rica | 152.1 | 76.31 | 68.45 |
| Croatia | 118.89 | 56.21 | 53.49 |
| Cuba | 276.67 | 93.33 | 86.48 |
| Cyprus | NA | NA | NA |
| Czechia | 150.28 | 65.24 | 63.73 |
| Côte d'Ivoire | 25.89 | 17.58 | 7.68 |
| Democratic People's Republic of Korea | NA | NA | NA |
| Democratic Republic of the Congo | 0.34 | 0.27 | 0.13 |
| Denmark | 202.01 | 79.79 | 76.63 |
| Djibouti | NA | NA | NA |
| Dominica | 81.32 | 42.24 | 39.09 |
| Dominican Republic | 126.1 | 61.88 | 50.88 |
| Ecuador | 154.93 | 79.36 | 71.01 |
| Egypt | 51.8 | 31.85 | 20.57 |
| El Salvador | 152.31 | 70.81 | 65.73 |
| Equatorial Guinea | NA | NA | NA |
| Eritrea | 137.64 | 63.67 | 62.61 |
| Estonia | 34.2 | 28.18 | 25.62 |
| Eswatini | 8.88 | 7.59 | NA |
| Ethiopia | 182.88 | 78.04 | 76.35 |
| Fiji | 136.89 | 71.29 | 65.6 |
| Finland | 175.44 | NA | NA |
| France | 186.54 | 78.72 | 73.69 |
| Gabon | NA | NA | NA |
| Gambia | 10.89 | 9.72 | 9.22 |
| Georgia | 67.75 | 36.74 | 31.01 |
| Germany | 187.3 | 75.49 | 71.31 |
| Ghana | NA | NA | NA |
| Greece | 171.52 | 72.32 | 68.31 |
| Greenland | 139.06 | 71.63 | 67.42 |
| Grenada | NA | NA | NA |
| Guam | NA | NA | NA |
| Guatemala | 64.1 | 36.86 | 26.67 |
| Guinea | 24.6 | 17.75 | 9.29 |
| Guinea-Bissau | 19.67 | 18.57 | 1.1 |
| Guyana | 88.81 | 50.83 | 36.52 |
| Haiti | 1.73 | 1.08 | 0.65 |
| Honduras | 91.91 | 47.11 | 41.18 |
| Hungary | 152.69 | 62.93 | 60.16 |
| Iceland | 192.69 | 77.34 | 76.19 |
| India | 104.59 | 61 | 43.59 |
| Indonesia | 99.73 | 60.7 | 41.68 |
| Iran (Islamic Republic of) | 134.75 | 67.56 | 58.65 |
| Iraq | 32.43 | 19.33 | 13.04 |
| Ireland | 198.74 | 78.52 | 77.4 |
| Israel | 180.28 | 70.06 | 63.16 |
| Italy | 192.75 | 82.58 | 75.96 |
| Jamaica | 43.09 | 25.31 | 20.24 |
| Japan | 163 | 81.67 | 80.46 |
| Jordan | 73.24 | 38.38 | 34.86 |
| Kazakhstan | 90.56 | 46.47 | 44.08 |
| Kenya | 19.17 | NA | 8.05 |
| Kiribati | 79.67 | 50.65 | 29.02 |
| Kuwait | 165.44 | 78.27 | 75.55 |
| Kyrgyzstan | 34.37 | 18.72 | 14.95 |
| Lao People's Democratic Republic | NA | 60.97 | 48.38 |
| Latvia | 139.97 | 71.22 | 68.51 |
| Lebanon | 81.2 | 41.75 | 33.66 |
| Lesotho | 37.21 | 33.92 | 31.55 |
| Liberia | NA | NA | NA |
| Libya | 39.34 | 27.22 | 12.12 |
| Lithuania | 152.42 | 69.93 | 66.8 |
| Luxembourg | 169.47 | 72.01 | NA |
| Madagascar | 3.28 | 2.76 | 2.59 |
| Malawi | 8.9 | 7.32 | 3.53 |
| Malaysia | 172.97 | 76.75 | 75.67 |
| Maldives | 151.19 | 75.78 | 70.25 |
| Mali | 5.3 | 3.99 | 2.29 |
| Malta | 207.51 | 83.95 | 81.86 |
| Marshall Islands | NA | NA | NA |
| Mauritania | 40.9 | 29.24 | 19.48 |
| Mauritius | 156.75 | NA | 70.02 |
| Mexico | 117.31 | 64.4 | 57.24 |
| Micronesia (Federated States of) | NA | NA | NA |
| Monaco | NA | NA | NA |
| Mongolia | 157.53 | 66.69 | 63.69 |
| Montenegro | 101.69 | 45.35 | 43.64 |
| Morocco | 135.63 | 65.86 | 61.53 |
| Mozambique | 44.64 | 26.19 | 18.46 |
| Myanmar | 67.16 | 38.42 | 28.74 |
| Namibia | 25.08 | 15.79 | 13.4 |
| Nauru | NA | NA | NA |
| Nepal | 82.74 | 47.68 | 35.06 |
| Netherlands | 159.45 | 70.68 | 65.97 |
| New Zealand | 159.12 | 76.63 | 74.53 |
| Nicaragua | 112.04 | 70.22 | 41.82 |
| Niger | NA | NA | NA |
| Nigeria | 6.79 | 4.72 | 2.05 |
| Niue | NA | NA | NA |
| North Macedonia | 83.83 | 40.26 | 39.01 |
| Northern Mariana Islands | NA | NA | NA |
| Norway | 183.91 | 79.06 | 72.48 |
| Oman | 133.69 | 68.22 | 63.42 |
| Pakistan | 68.12 | 41.92 | 30.95 |
| Palau | NA | NA | NA |
| Palestine | 64.44 | NA | 28.87 |
| Panama | 141.67 | 70.85 | 63.72 |
| Papua New Guinea | 5.04 | 2.83 | 2.21 |
| Paraguay | 100.78 | 51.17 | 43.48 |
| Peru | 153.02 | 73.3 | 65.66 |
| Philippines | 95.95 | NA | 44.21 |
| Poland | 119.78 | 54.66 | 53.06 |
| Portugal | 195.15 | 91.5 | 83.15 |
| Puerto Rico | NA | NA | NA |
| Qatar | 194.64 | NA | NA |
| Republic of Korea | 203.63 | 85.02 | 82.09 |
| Republic of Moldova | 54.72 | NA | 30.13 |
| Romania | 80.98 | NA | 39.92 |
| Russian Federation | 101.4 | 51.01 | 46.3 |
| Rwanda | 91.38 | 55.38 | 39.45 |
| Saint Kitts and Nevis | 115.94 | 58.05 | 53.98 |
| Saint Lucia | 58.6 | 31.05 | 27.55 |
| Saint Vincent and the Grenadines | 58.87 | 32.22 | 25.24 |
| Samoa | NA | NA | NA |
| San Marino | 168.59 | 72.33 | 64.37 |
| Sao Tome and Principe | NA | NA | NA |
| Saudi Arabia | 142.73 | 68.88 | 63.95 |
| Senegal | NA | NA | NA |
| Serbia | 120.65 | 48.26 | 46.87 |
| Seychelles | 171.25 | 76.86 | 73.05 |
| Sierra Leone | NA | NA | NA |
| Singapore | 212.41 | 86.62 | 83.18 |
| Slovakia | 91.92 | 45.04 | 43.86 |
| Slovenia | 131.4 | 59.07 | 56.22 |
| Solomon Islands | 33.98 | 25.61 | 8.37 |
| Somalia | 8.55 | 6.84 | 4.6 |
| South Africa | 47 | 31.83 | 26.68 |
| South Sudan | NA | NA | NA |
| Spain | 178.29 | 83.96 | 79.99 |
| Sri Lanka | 155.71 | 73.27 | 63.44 |
| Sudan | NA | NA | NA |
| Suriname | 79.44 | 42.39 | 37.05 |
| Sweden | 168.7 | 72.09 | 68.95 |
| Switzerland | 161.31 | 68.39 | NA |
| Syrian Arab Republic | 13.22 | NA | 3.99 |
| Taiwan (Province of China) | 146.72 | 78.44 | 68.28 |
| Tajikistan | 69.26 | 38.63 | 30.63 |
| Thailand | 147.75 | 71.81 | 65.01 |
| Timor-Leste | 90.22 | 49.62 | 40.59 |
| Togo | 27.28 | 15.73 | 11.54 |
| Tokelau | NA | NA | NA |
| Tonga | 127.98 | 69.14 | 58.85 |
| Trinidad and Tobago | 92.71 | 46.77 | 43.85 |
| Tunisia | 99.8 | 56.41 | 48.69 |
| Turkey | 157.25 | 66.8 | 60.65 |
| Turkmenistan | NA | NA | NA |
| Tuvalu | NA | NA | NA |
| Uganda | NA | NA | NA |
| Ukraine | 72.29 | 37.22 | 35.08 |
| United Arab Emirates | 240.71 | NA | 96.69 |
| United Kingdom | 198.9 | 76.84 | 70.46 |
| United Republic of Tanzania | 3.71 | 3.18 | 2.51 |
| United States of America | 158.6 | 73.9 | 63.58 |
| United States Virgin Islands | NA | NA | NA |
| Uruguay | 205 | 80.83 | 78.49 |
| Uzbekistan | 112.73 | NA | NA |
| Vanuatu | NA | NA | NA |
| Venezuela (Bolivarian Republic of) | NA | NA | NA |
| Viet Nam | 159.8 | 79.55 | 69.7 |
| Yemen | NA | NA | NA |
| Zambia | 9.07 | NA | 6.37 |
| Zimbabwe | 44.88 | 25.42 | 19.37 |

NA=not available; COVID-19=coronavirus disease 2019.

Supplementary Figures

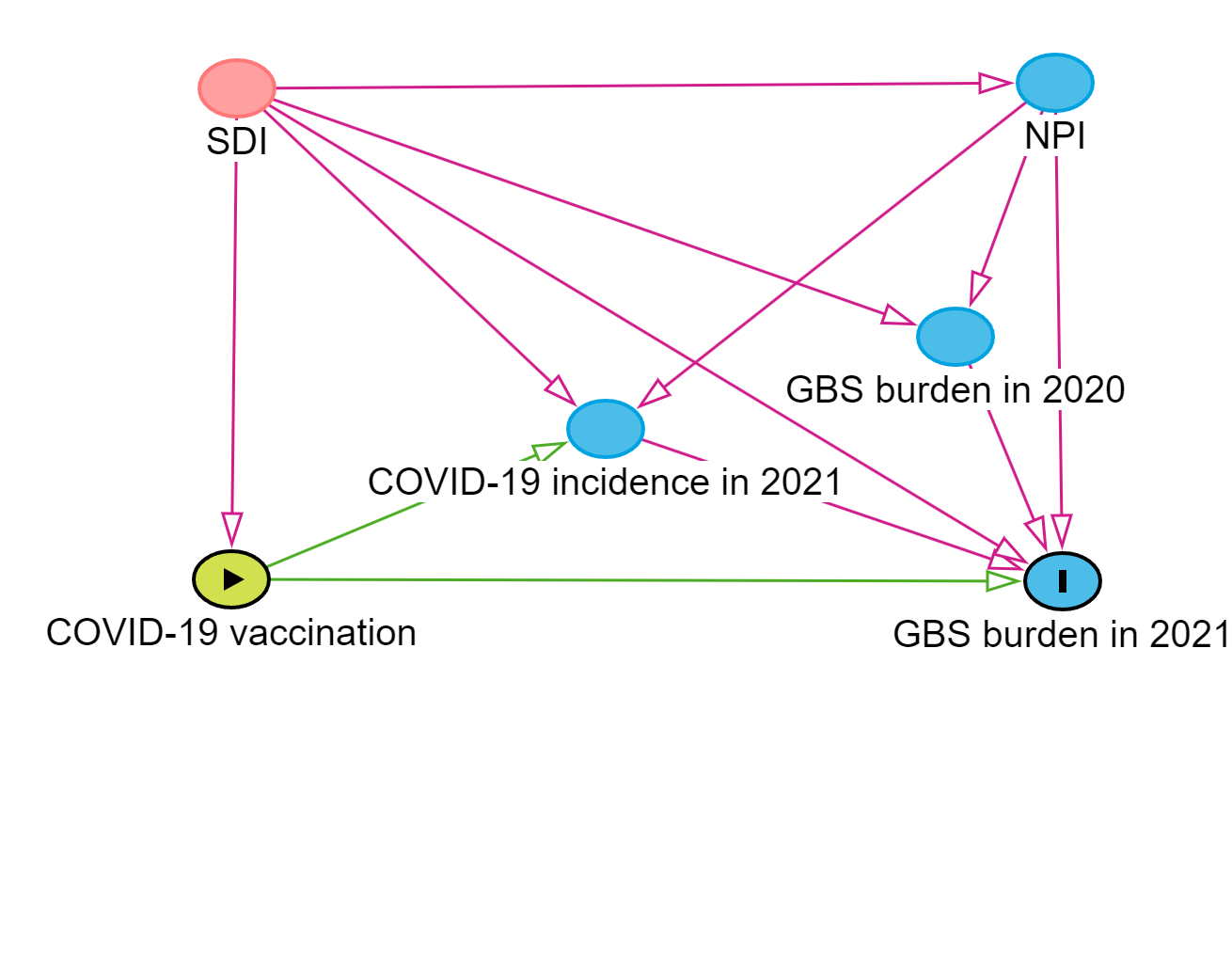

**Supplementary Figure S1** Directed acyclic graph for the association between

GBS= Guillain-Barré syndrome; NPI=non-pharmacological interventions (government stringency index was used to represent the degree of NPI); SDI=socio-demographic index.

Notes: Pink line: biasing path. Green line: causal path. Pink pie: confounder (ancestor of exposure and outcome). Green pie: exposure/ancestor of exposure. Blue pie: ancestor of outcome.

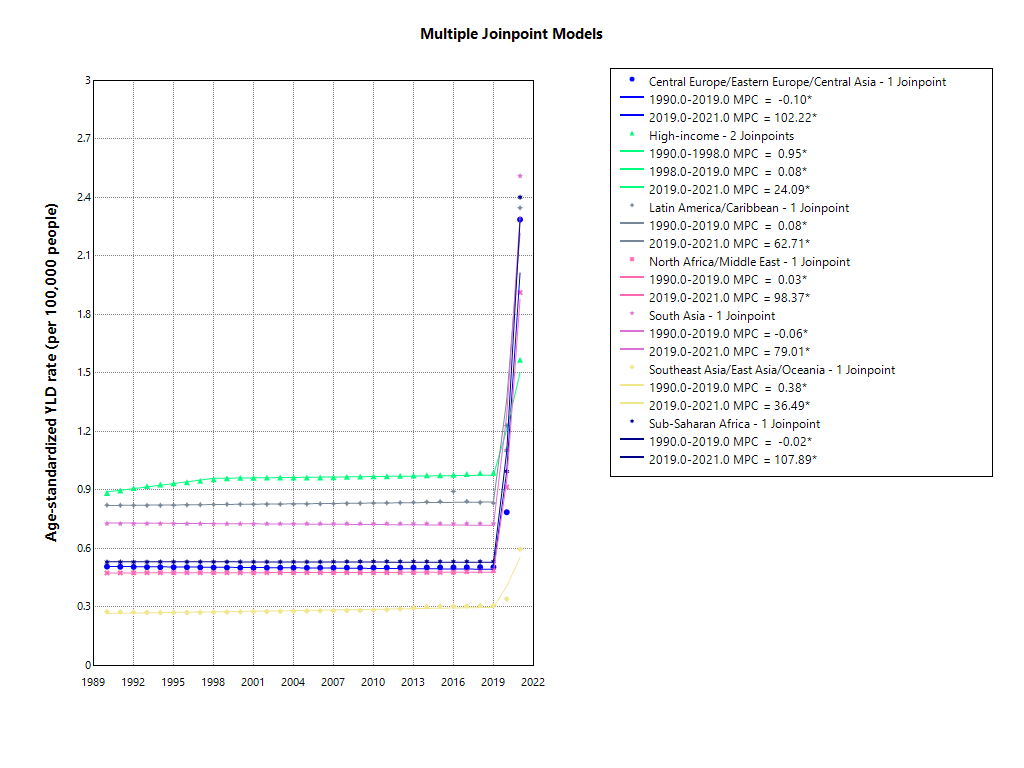

**Supplementary Figure S2** Joinpoint regression analysis of the age-standardized years lived with disability rate of Guillain-Barré syndrome by seven GBD super regions, 1990–2021.

Notes: APC=annual percentage change; GBD=global burden of disease study; Asterisks represent those values that are statistically significantly different from 0.

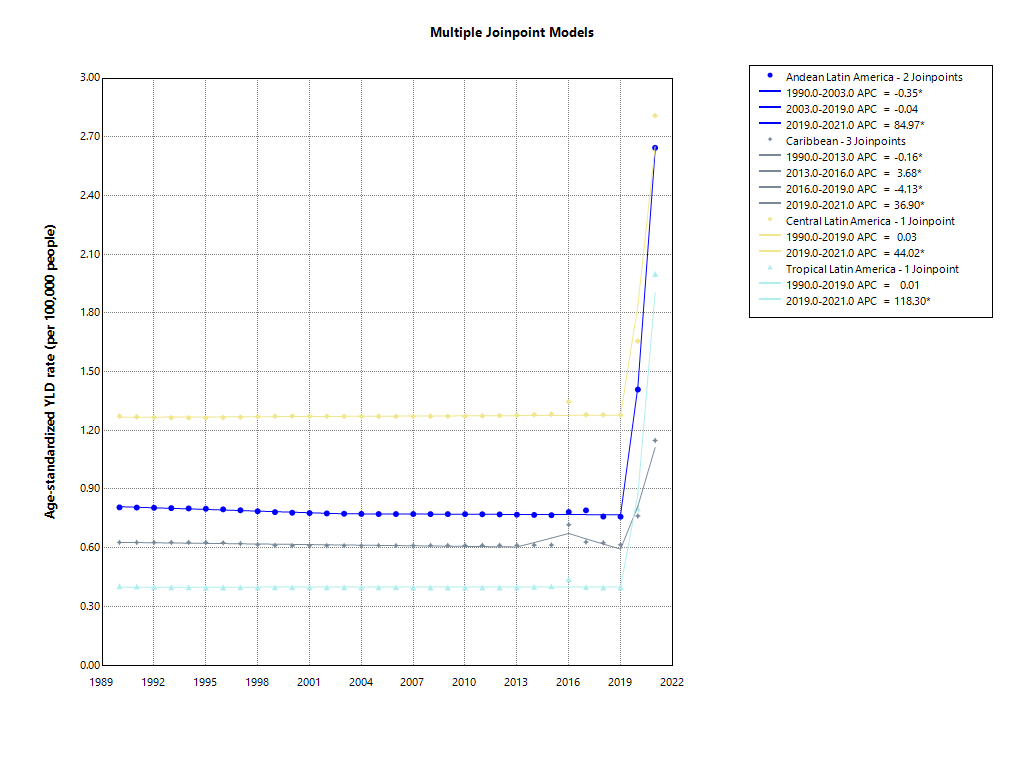

**Supplementary Figure S3** Joinpoint regression analysis of the age-standardized years lived with disability rate of Guillain-Barré syndrome by Latin America regions, 1990–2021.

Notes: APC=annual percentage change; Asterisks represent those values that are statistically significantly different from 0.

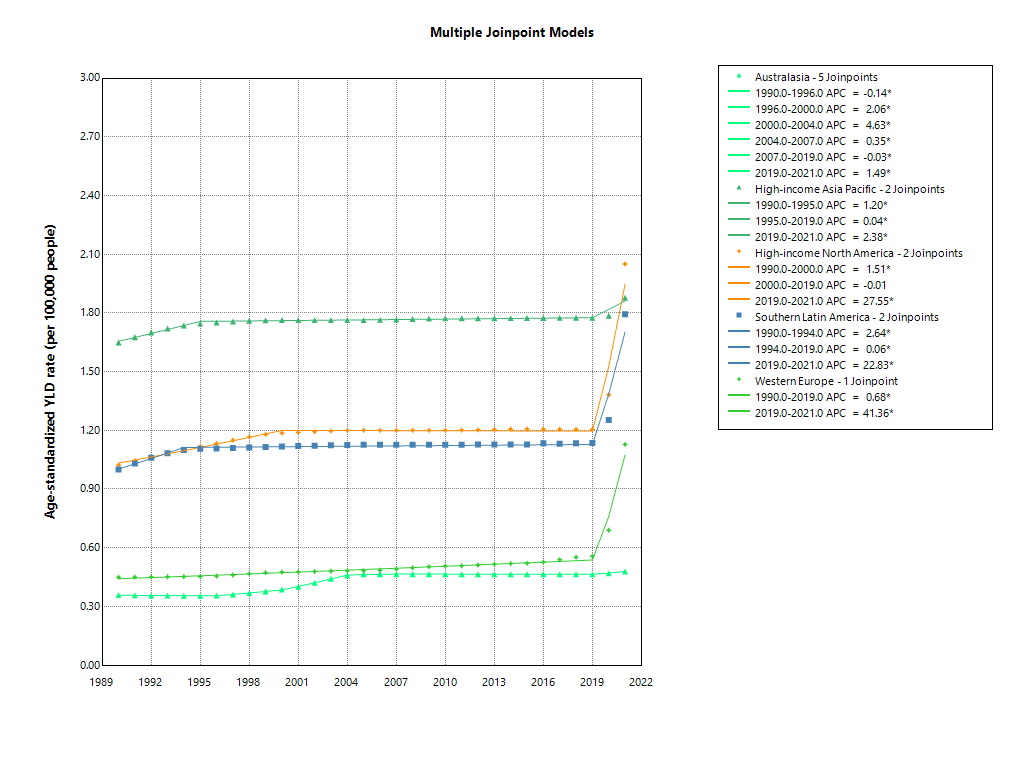

**Supplementary Figure S4** Joinpoint regression analysis of the age-standardized years lived with disability rate of Guillain-Barré syndrome by High-income regions, 1990–2021.

Notes: APC=annual percentage change; Asterisks represent those values that are statistically significantly different from 0.

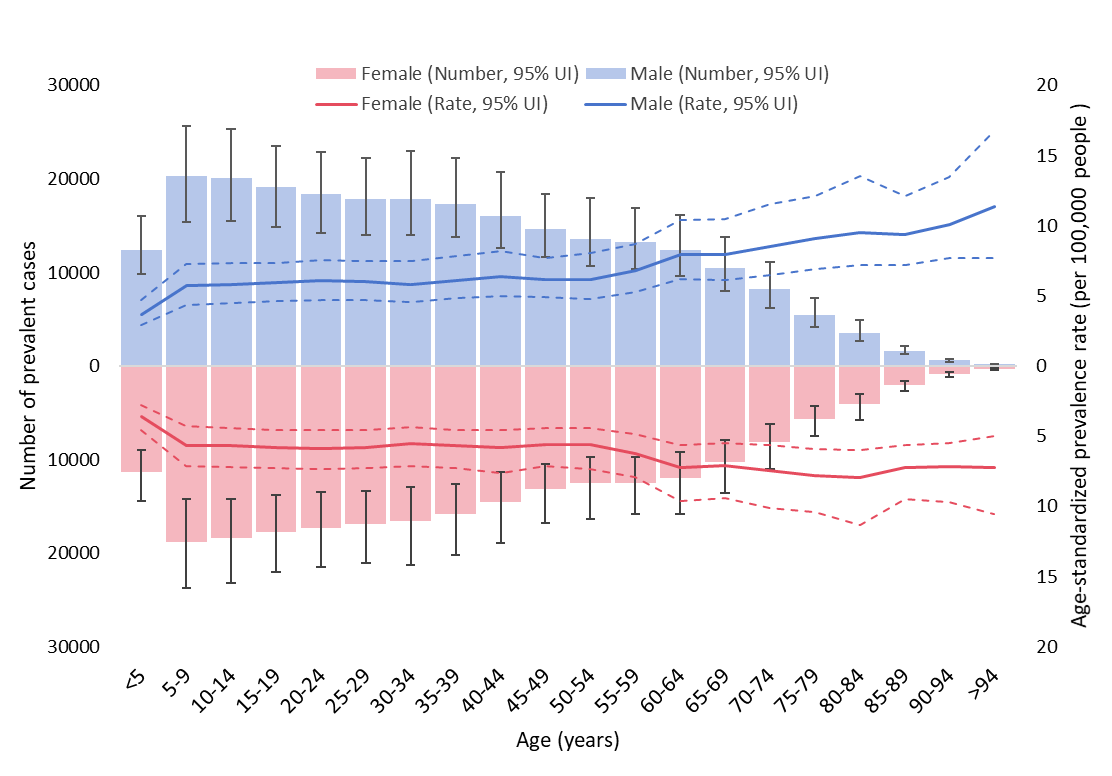

**Supplementary Figure S5** Age-sex distribution of prevalence of Guillain-Barré syndrome in 2021

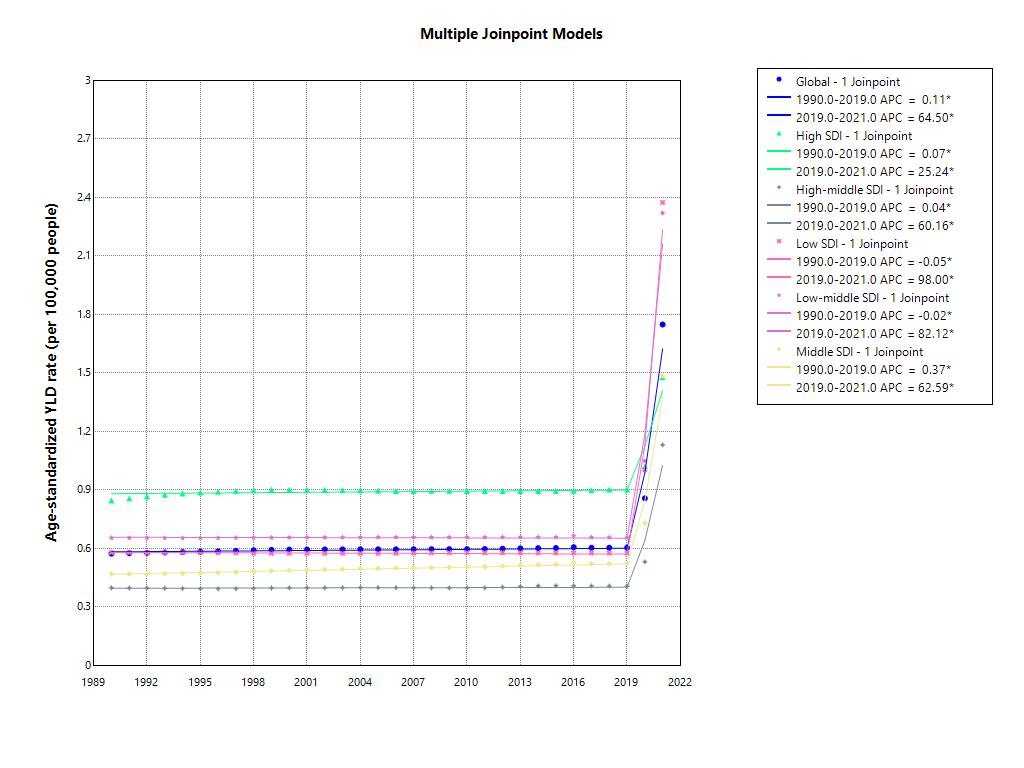
**Supplementary Figure S6** Joinpoint regression analysis of the age-standardized years lived with disability rate of Guillain-Barré syndrome globally and by quintile of the socio-demographic index, 1990–2021.

Notes: APC=annual percentage change; Asterisks represent those values that are statistically significantly different from 0.

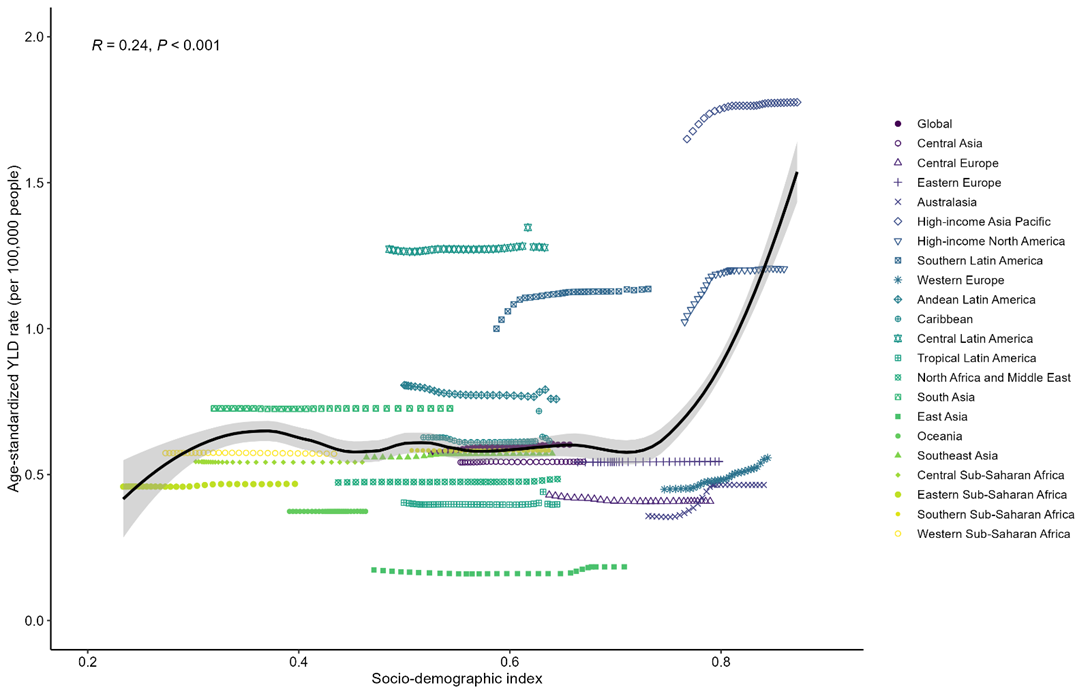

**Supplementary Figure S7** Age-standardised years lived with disability rates (YLDs) of Guillain-Barré syndrome versus Socio-demographic Index, globally and across 21 regions. Each point shows the observed age-standardised YLDs rate for each region from 1990 to 2019.

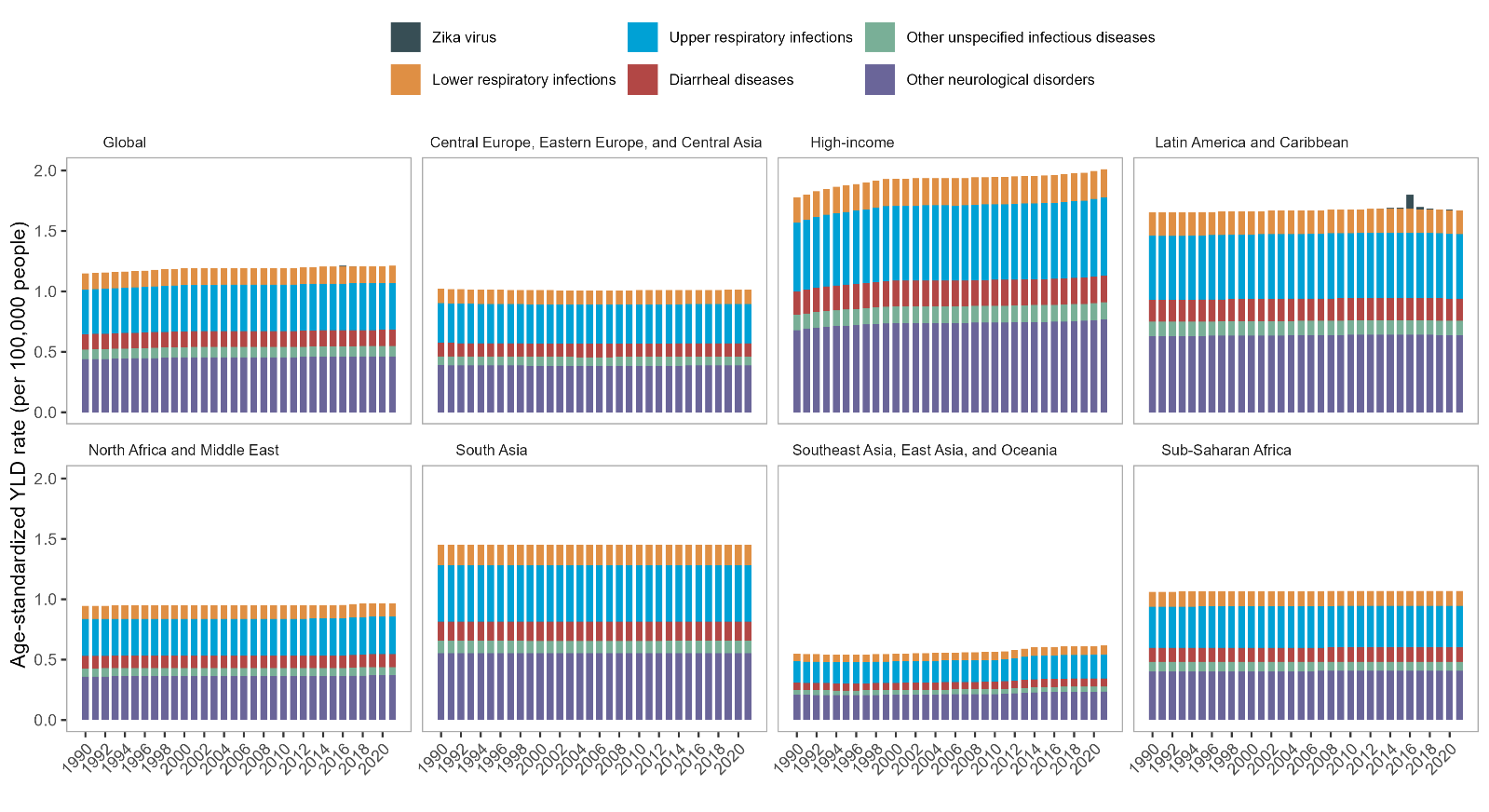

S**upplementary Figure S8** Age-standardized years lived with disability rates of Guillain-Barré syndrome attributed to causes excluding COVID-19, by super region and years from 1990-2021.

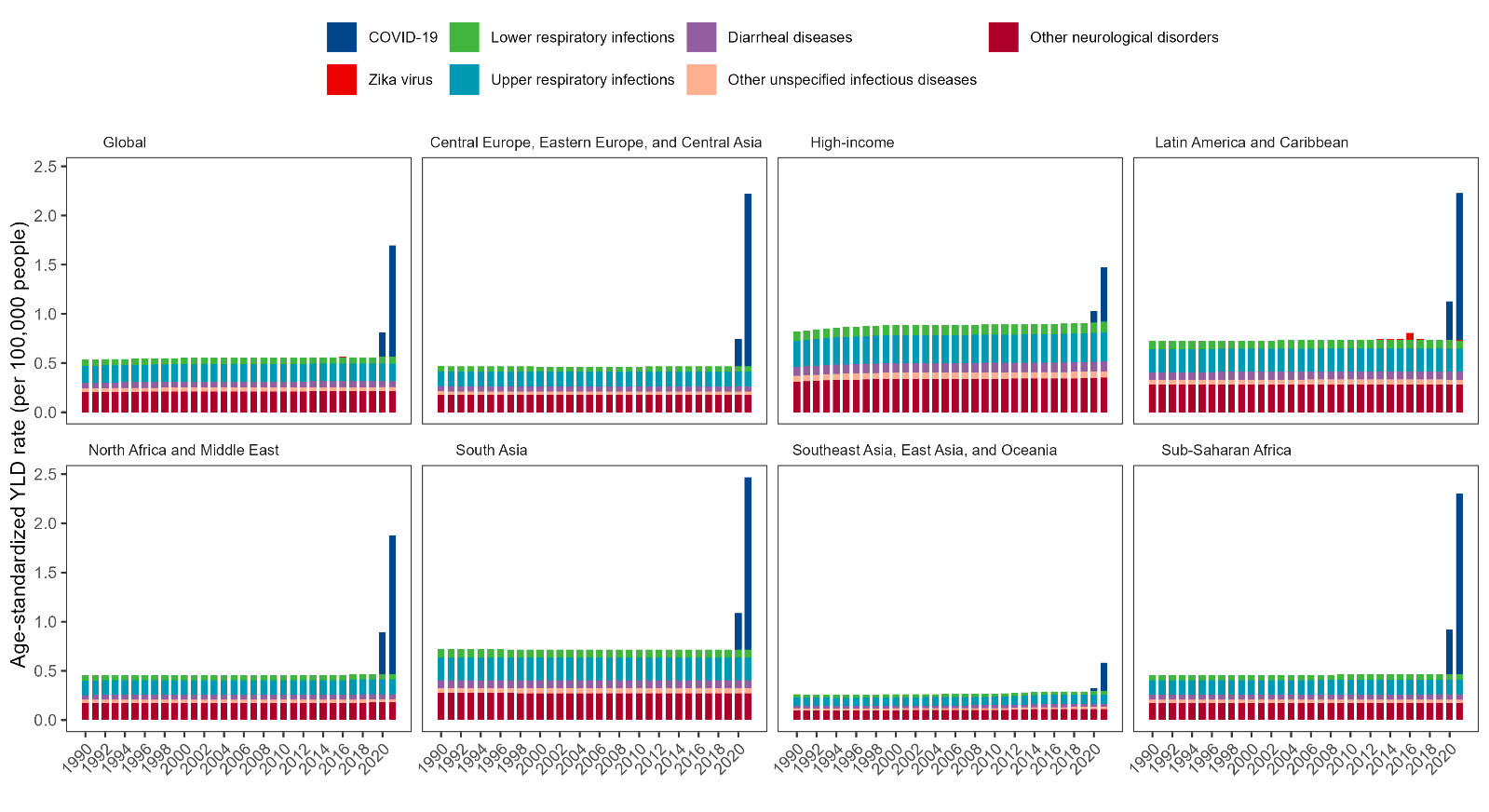

**Supplementary Figure S9** Age-standardized years lived with disability rates of Guillain-Barré syndrome attributed to underlying causes in females, by super region and years from 1990-2021.

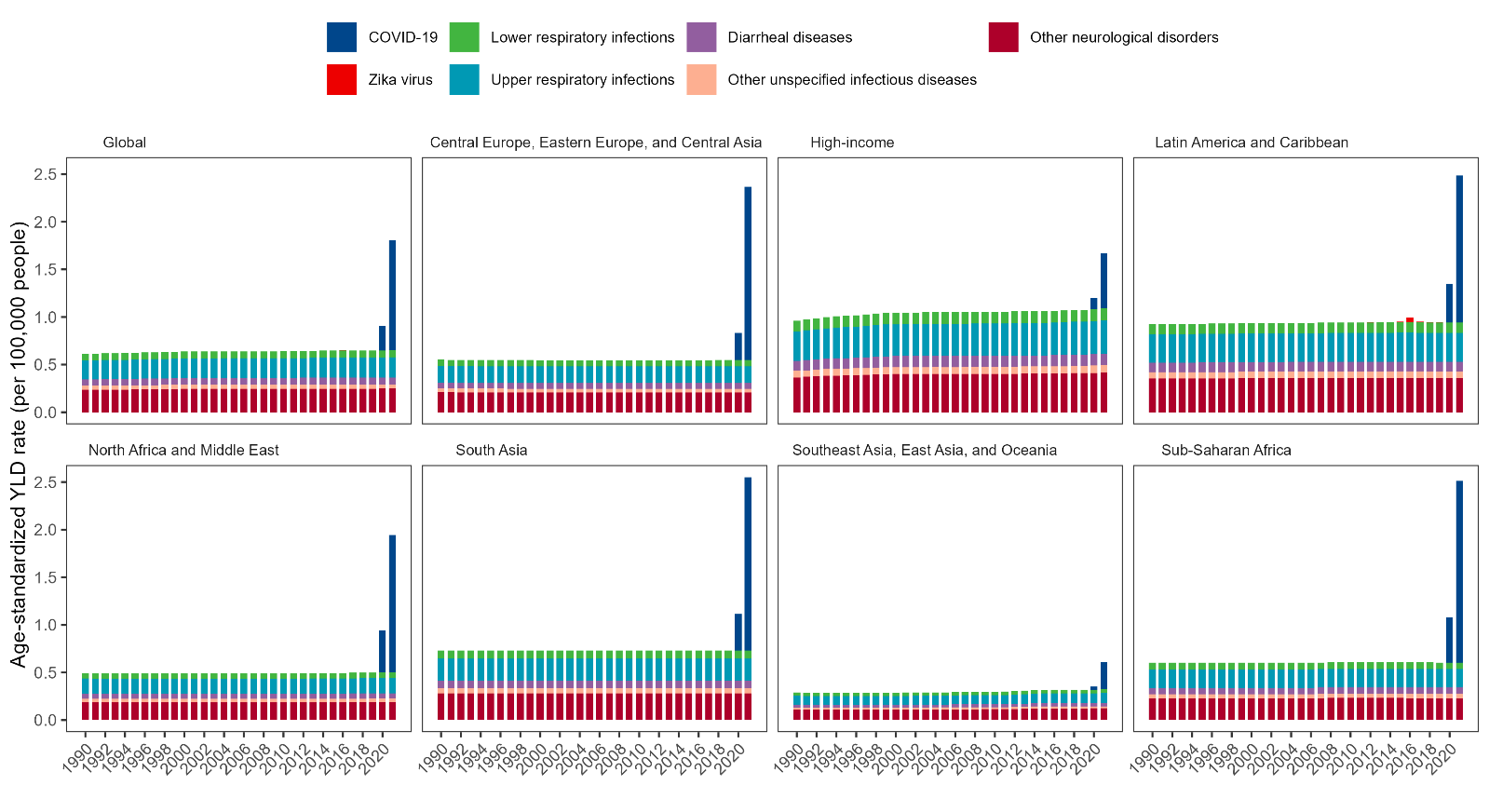

**Supplementary Figure S10** Age-standardized years lived with disability rates of Guillain-Barré syndrome attributed to underlying causes in males, by super region and years from 1990-2021.

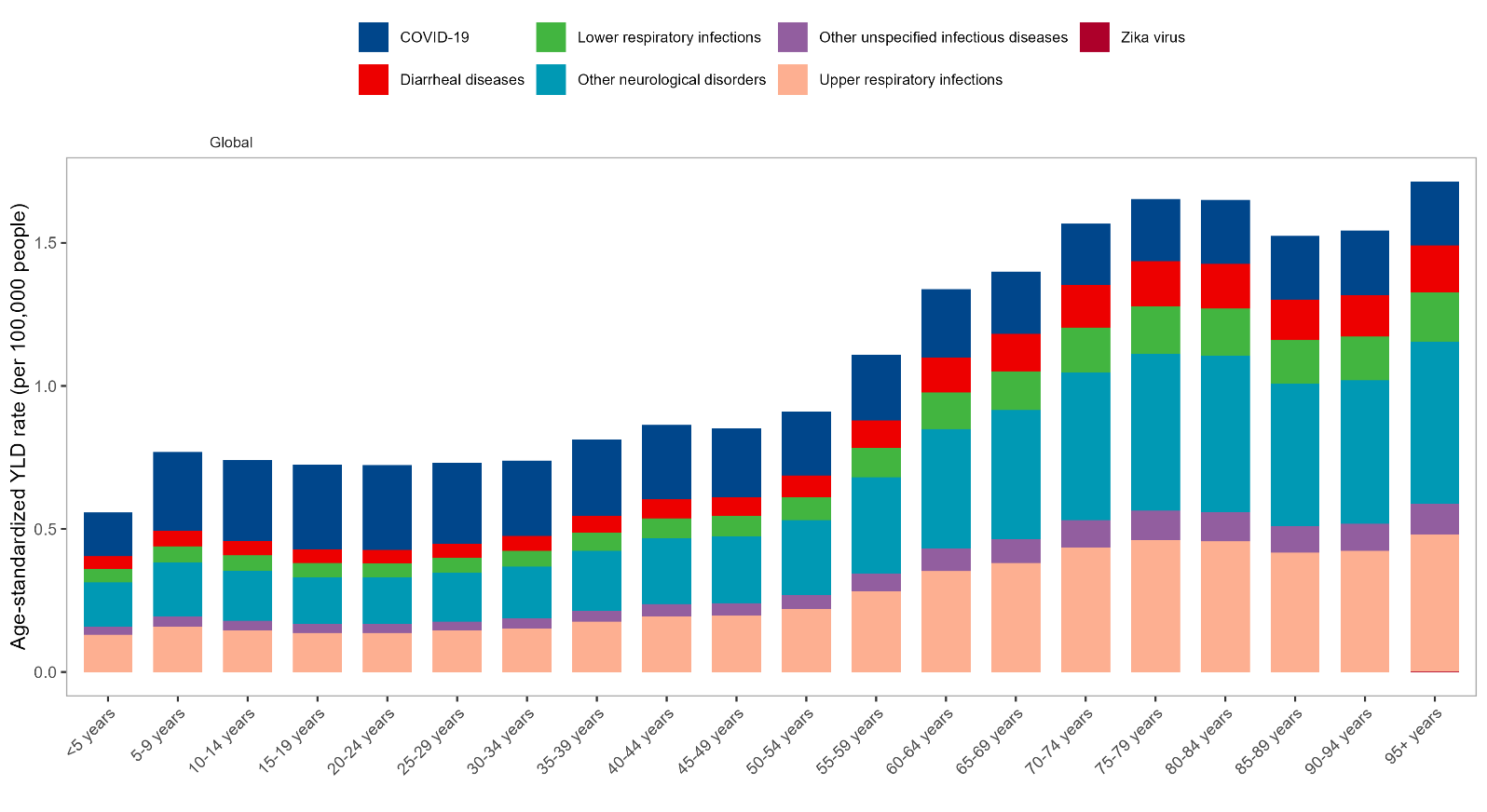

**Supplementary Figure S11** Years lived with disability rates of Guillain-Barré syndrome attributed to specific causes by age group in 2020.

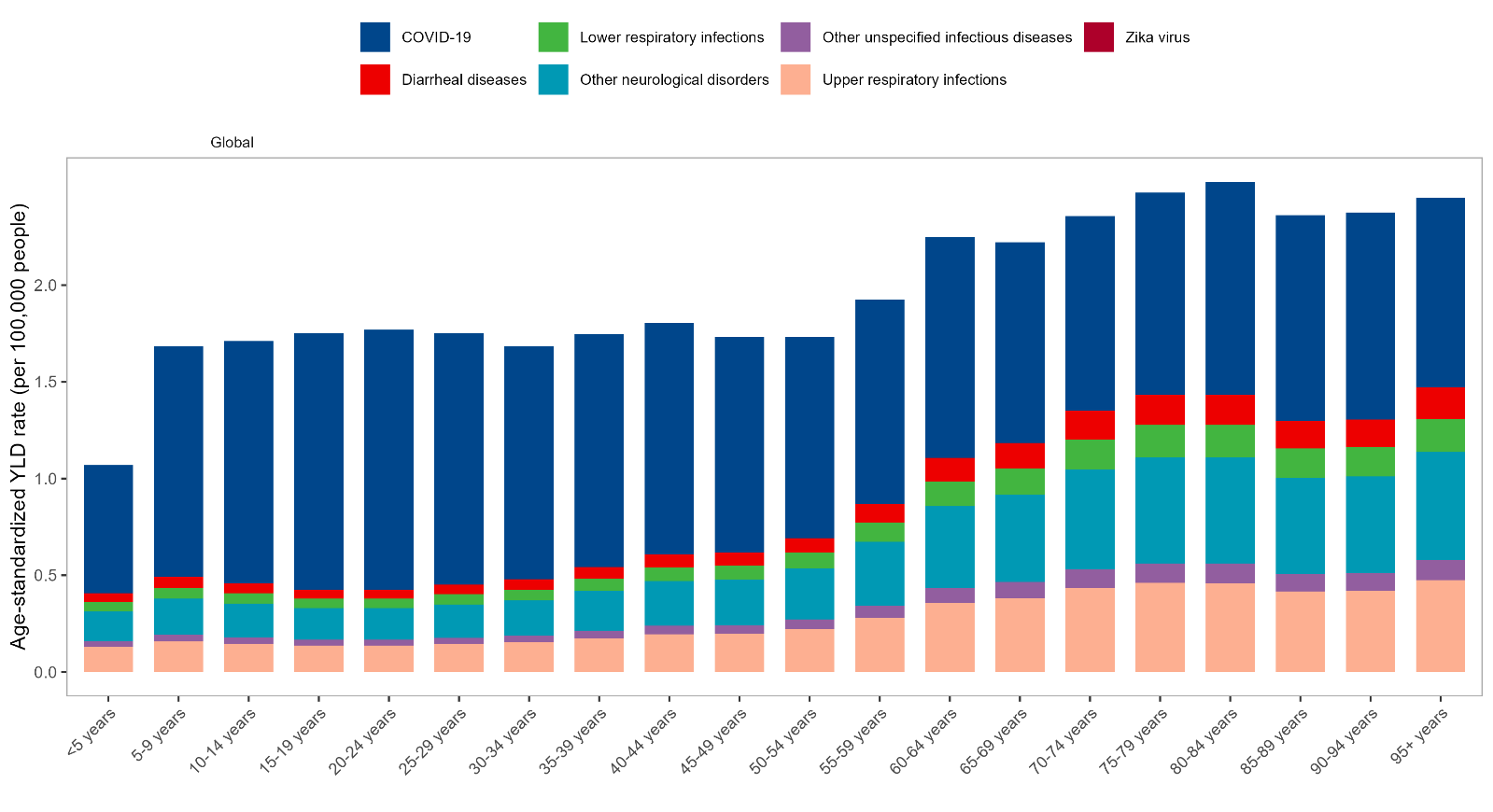

**Supplementary Figure S12** Years lived with disability rates of Guillain-Barré syndrome attributed to specific causes by age group in 2021.

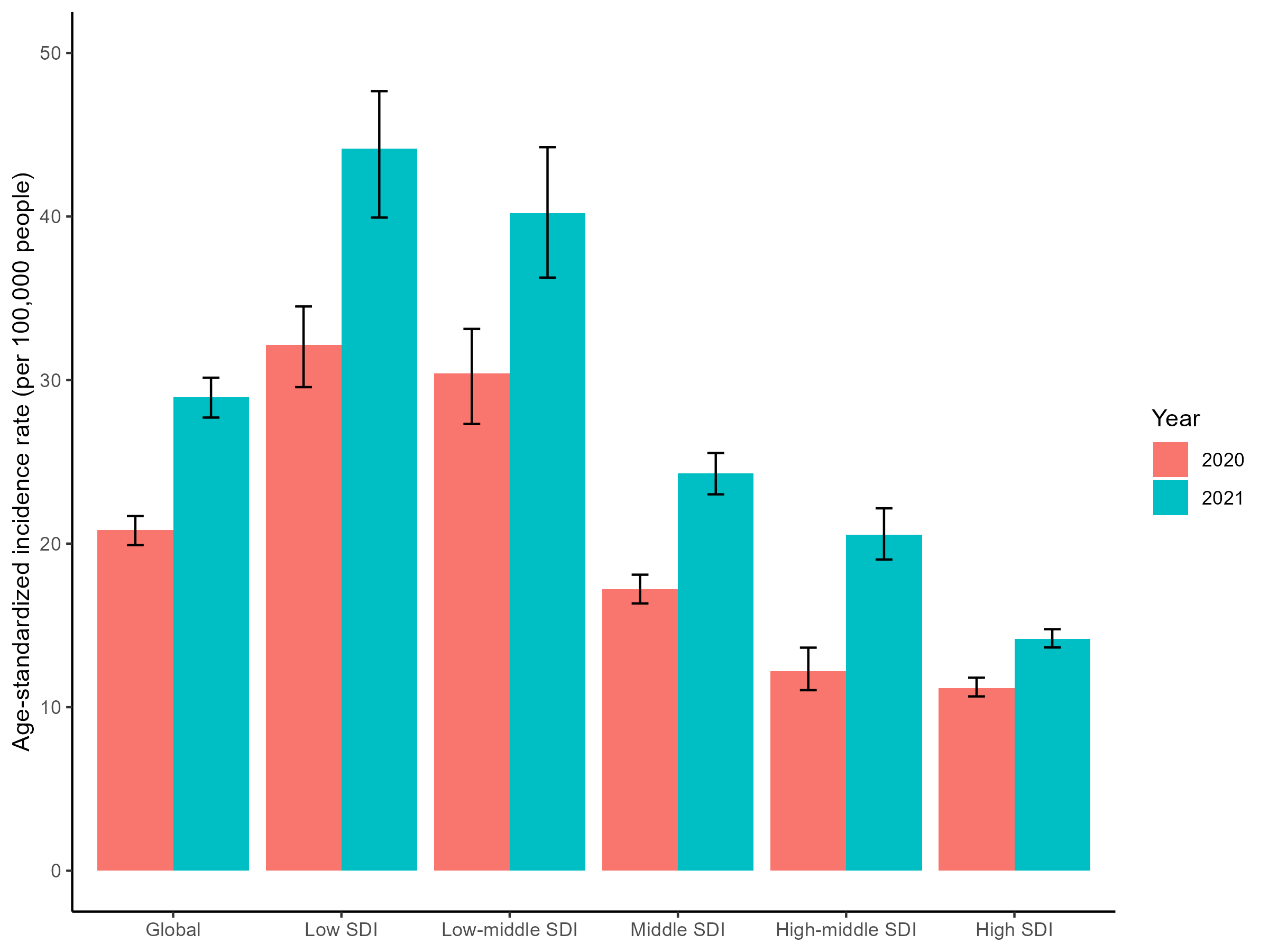

**Supplementary Figure S13** Age-standardized COVID-19 incidence by years and socio-demographic index

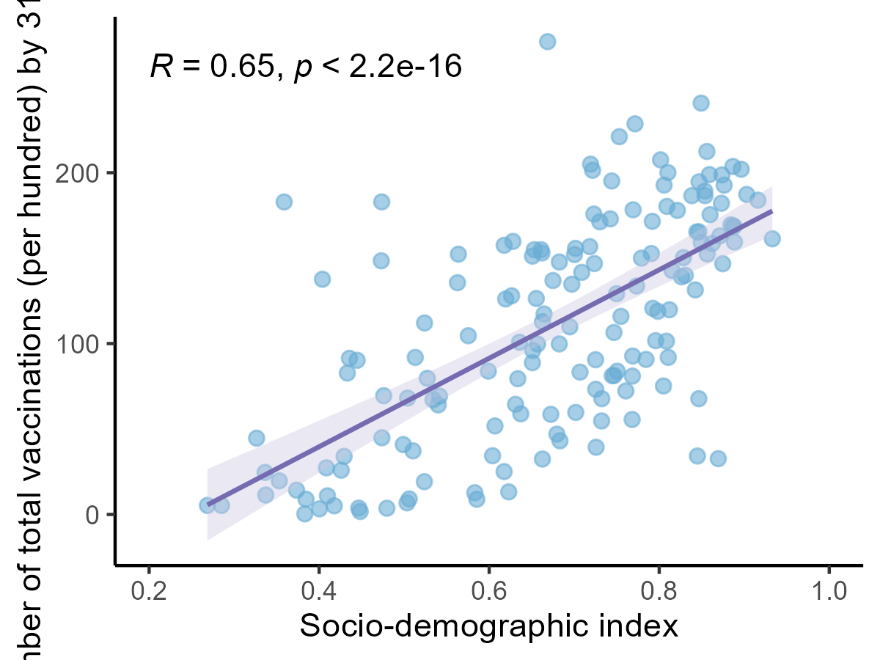

**Supplementary Figure S14** Relationship between socio-demographic index and vaccination rate (number of people vaccinated per hundred) by 31.12.2021.

Notes: R=Pearson’s correlation coefficient.

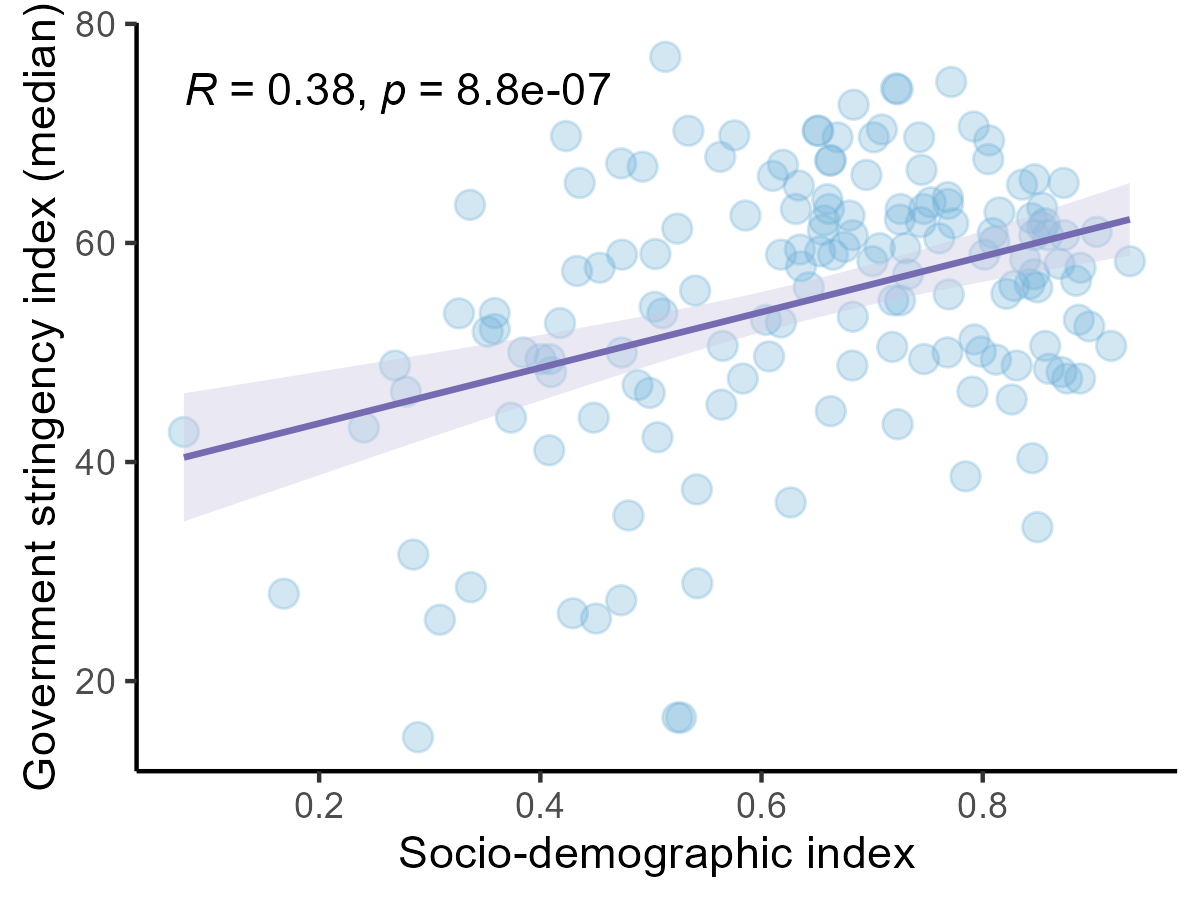

**Supplementary Figure S15** Relationship between Socio-demographic Index and median Government Stringency Index during the first two years of COVID-19 pandemic, 2020-2021.

Notes: R=Pearson’s correlation coefficient.

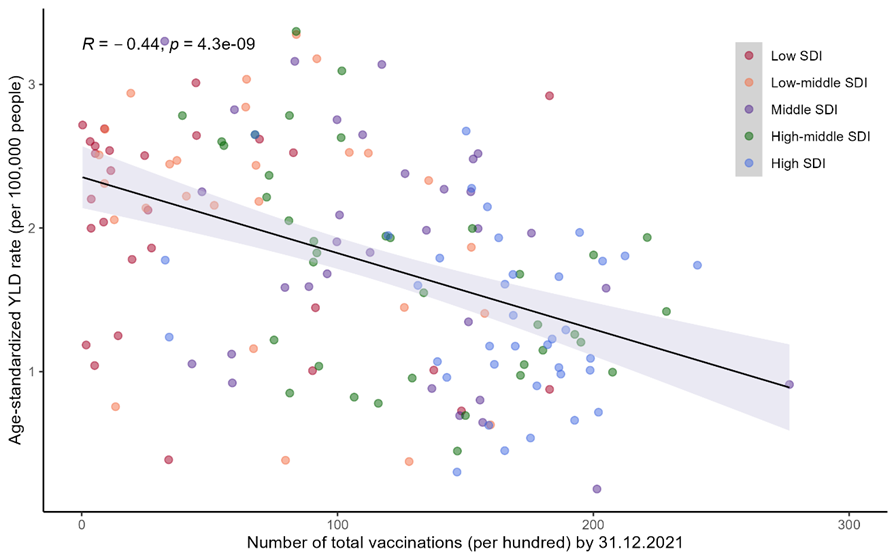
**Supplementary Figure 16** Number total vaccinations per hundred (data collected by 31.12.2021) versus all-cause age-standardized year lived with disability (YLD) rate of Guillain-Barré syndrome in 2021.

Notes: R=Pearson’s correlation coefficient.

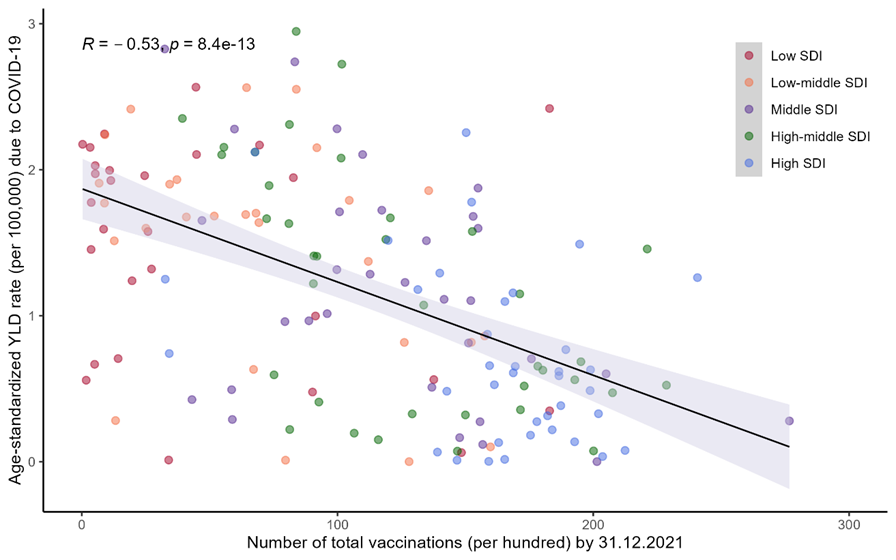

**Supplementary Figure 17** Number total vaccinations per hundred (data collected by 31.12.2021) versus age-standardized year lived with disability rate of Guillain-Barré syndrome due to COVID-19 in 2021.

Notes: R=Pearson’s correlation coefficient.

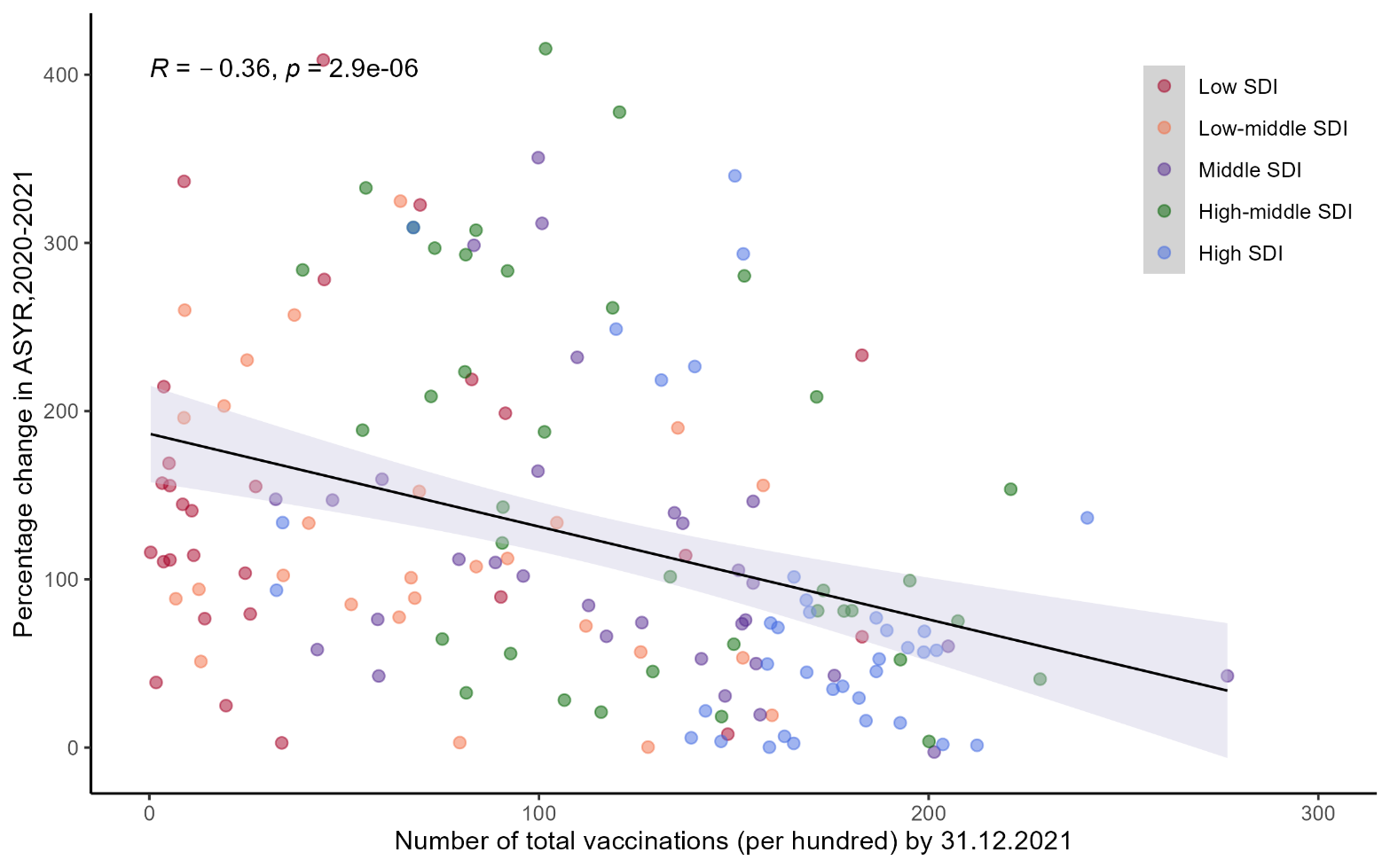

**Supplementary Figure S18** Relationship between percentage change in age-standardized year lived with disability rate of Guillain-Barré syndrome from 2019 to 2021 and number of total vaccinations (per hundred) by December 12, 2021.

Notes: R=Pearson’s correlation coefficient.

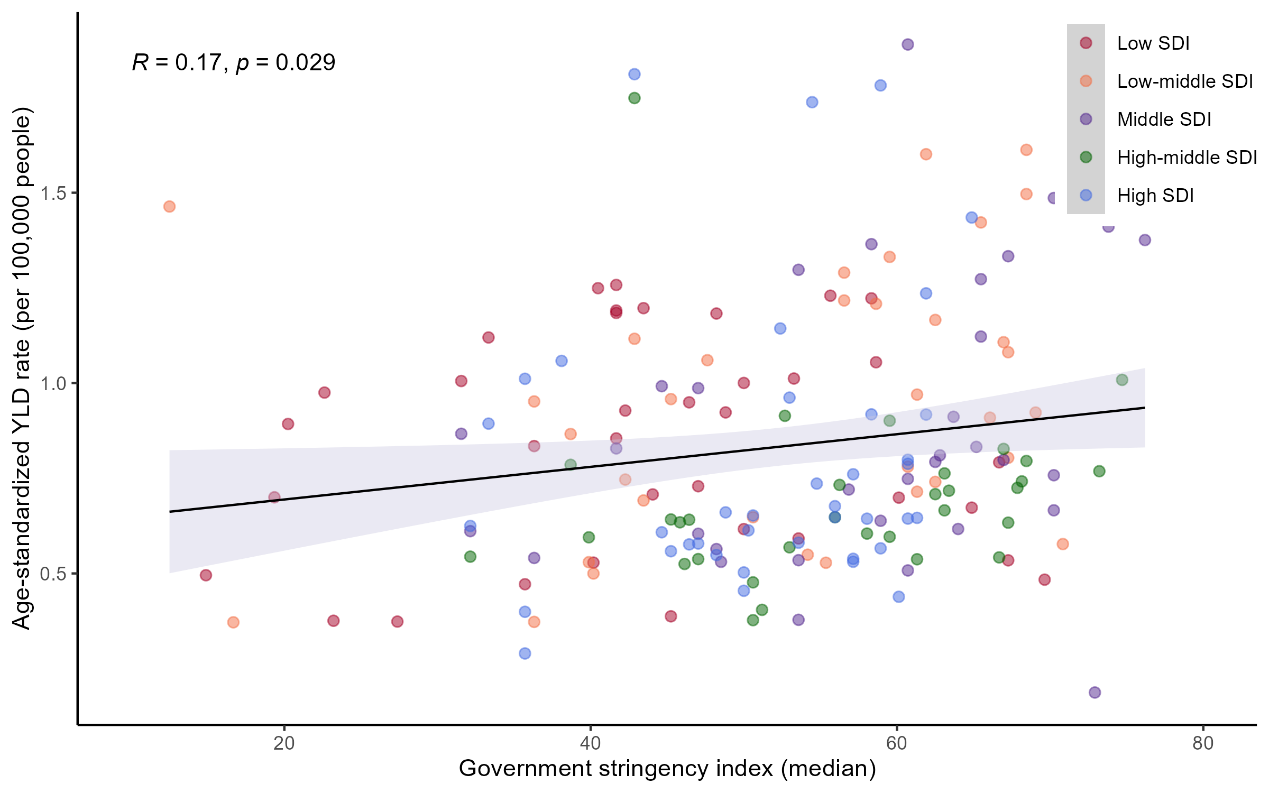

**Supplementary Figure S19** Relationship between all-cause age-standardized years lived with disability rate of Guillain-Barré syndrome and median of Government Stringency in 2020.

Notes: R=Pearson’s correlation coefficient.

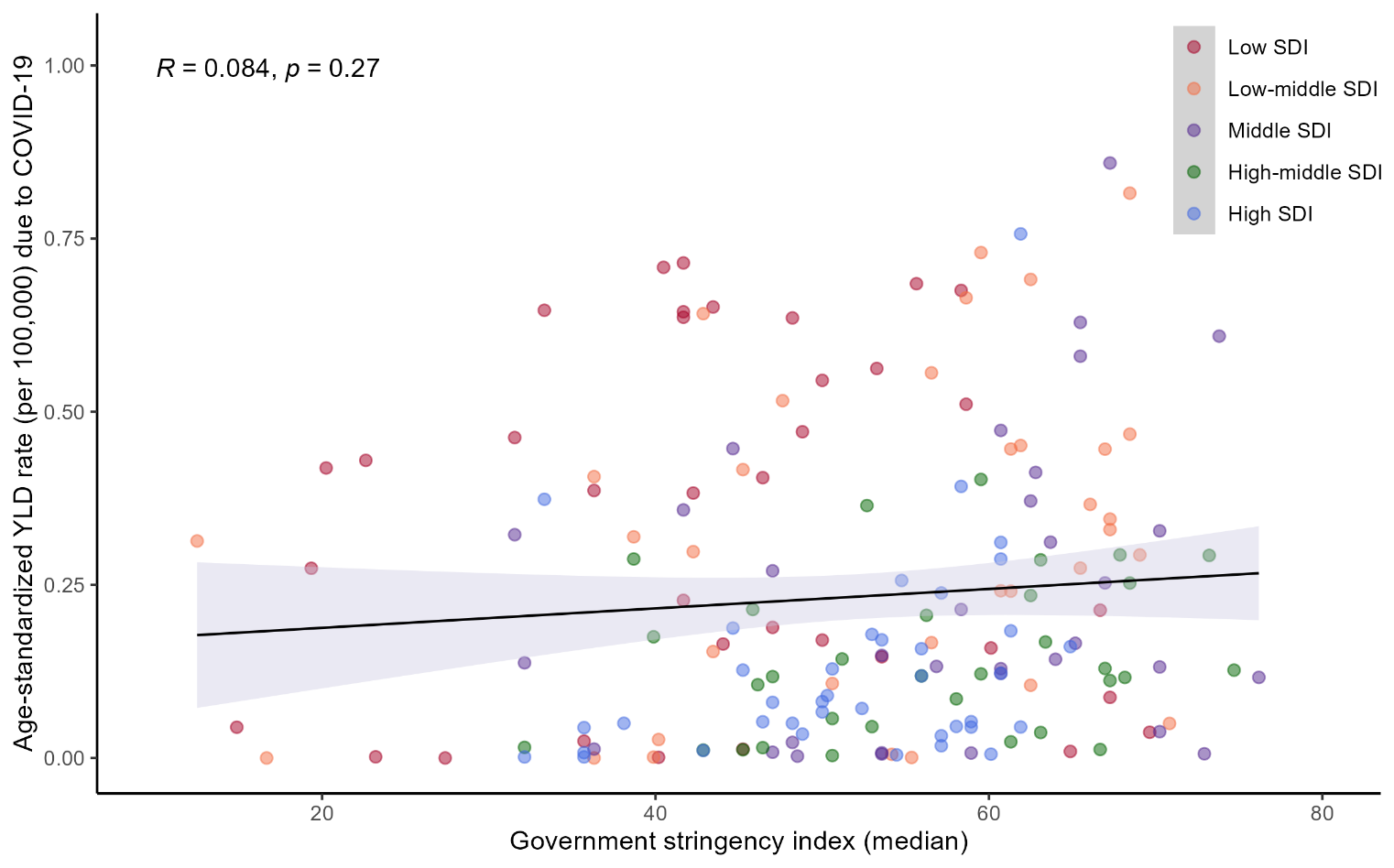

**Supplementary Figure S20** Relationship between all-cause age-standardized years lived with disability rate of Guillain-Barré syndrome due to COVID-19 and median Government Stringency in 2020.

Notes: R=Pearson’s correlation coefficient.

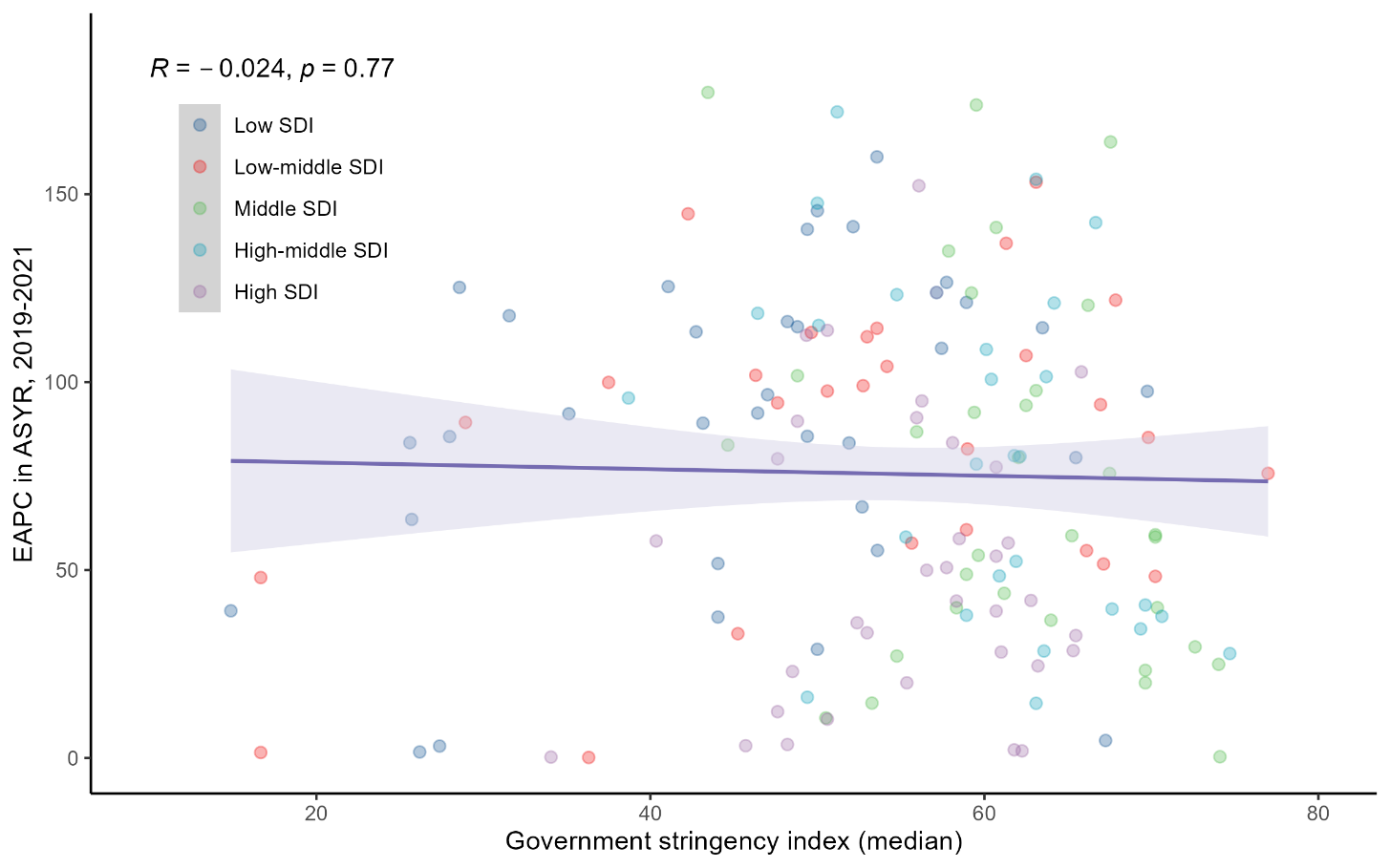

**Supplementary Figure S21** Relationship between percentage change in age-standardized year lived with disability rate of Guillain-Barré syndrome from 2019 to 2021 and median government stringency index in 2020 and 2021.

Notes: R=Pearson’s correlation coefficient.

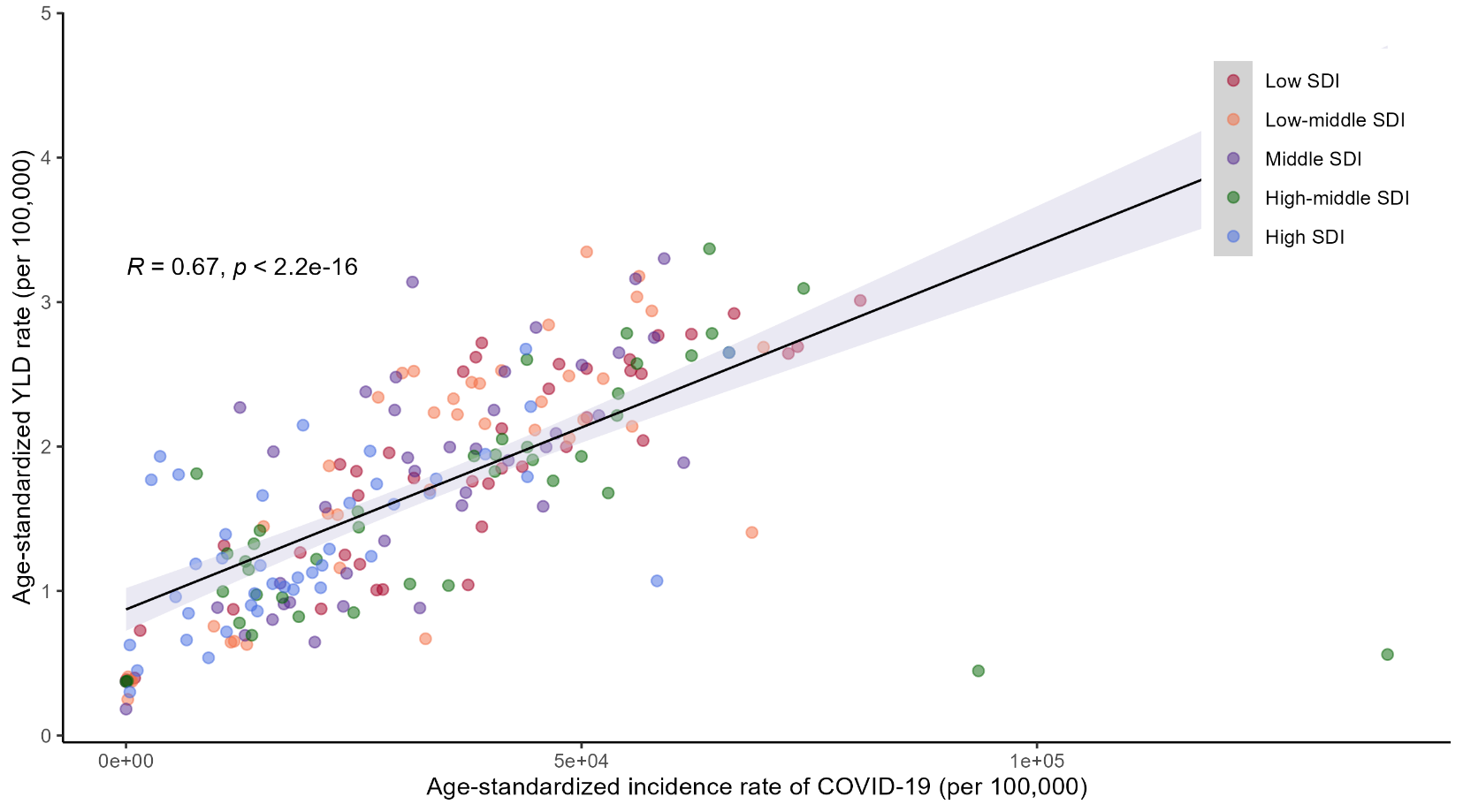

**Supplementary Figure S22** Relationship between age-standardized COVID-19 incidence and age-standardized years lived with disability of Guillain-Barré syndrome in 2021.

Notes: R=Pearson’s correlation coefficient.

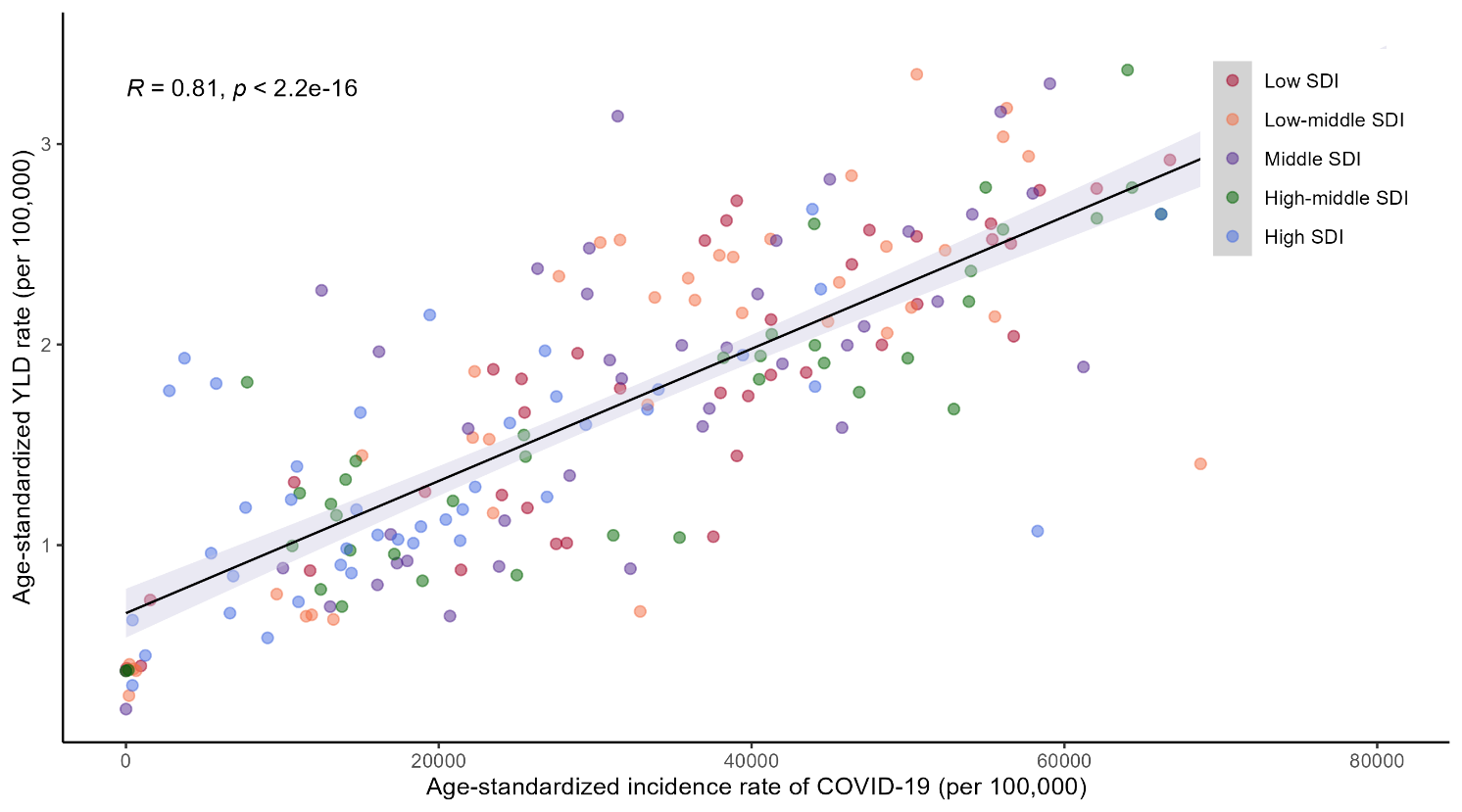

**Supplementary Figure S23** Relationship between age-standardized COVID-19 incidence and age-standardized years lived with disability of Guillain-Barré syndrome in 2021, a sensitivity analysis excluding two outliers (Northern Mariana Islands and American Samoa).

Notes: R=Pearson’s correlation coefficient.
